# Genome-wide association studies of thyroid-related hormones, dysfunction, and autoimmunity among 85,421 Chinese pregnancies

**DOI:** 10.1101/2024.07.01.24309813

**Authors:** Yuandan Wei, Jianxin Zhen, Liang Hu, Yuqin Gu, Yanhong Liu, Xinxin Guo, Zijing Yang, Hao Zheng, Shiyao Cheng, Fengxiang Wei, Likuan Xiong, Siyang Liu

**Affiliations:** School of Public Health (Shenzhen), Shenzhen Campus of Sun Yat-sen University, Shenzhen, Guangdong 518107, China; Central Laboratory, Shenzhen Baoan Women’s and Children’s Hospital, Shenzhen, Guangdong 518102, China; Longgang District Maternity & Child Healthcare Hospital of Shenzhen City (Longgang Maternity and Child Institute of Shantou University Medical College), Shenzhen, Guangdong 518172, China; Shenzhen Key Laboratory of Birth Defects Research, Shenzhen, Guangdong 518102, China

## Abstract

Maintaining normal thyroid function is crucial in pregnancy, yet thyroid dysfunction and the presence of thyroid peroxidase antibodies (TPOAb) affect 0.5% to 18% of pregnant women. Here, we conducted a genome-wide association study (GWAS) of eight thyroid traits, including two thyroid-related hormones, four thyroid dysfunctions, and two thyroid autoimmunity measurements among 85,421 Chinese pregnant women to investigate the genetic basis of thyroid function during pregnancy. Our study identified 176 genetic loci, including 125 previously unknown genome-wide associations. Joint epidemiological and Mendelian randomization analyses revealed significant associations between the gestational thyroid phenotypes and gestational complications, birth outcomes, and later-age health outcomes. Specifically, genetically elevated thyroid-stimulating hormone (TSH) levels during pregnancy correlated with lower glycemic levels, reduced blood pressure, and longer gestational duration. Additionally, TPOAb and thyroid functions during pregnancy share genetic correlations with later-age thyroid and cardiac disorders. These findings provide novel insights into the genetic determinants of thyroid traits during pregnancy, which may lead to new therapeutics, early pre-diagnosis and preventive strategies starting from early adulthood.

## Introduction

Pregnancy represents a crucial phase in human reproduction, impacting both maternal and offspring health. Particularly, normal maternal thyroid function and lack of autoimmunity during pregnancy, commonly assessed by measuring circulating thyroid-stimulating hormone (TSH), free thyroxine (FT4) levels, and presence and quantification of thyroid peroxidase antibodies (TPOAb), holds significant relevance for pregnancy outcomes and long-term health[1,2]. Thyroid dysfunctions during pregnancy are determined by these measurements and demonstrate varying prevalence, ranging from 0.5% for overt hypothyroidism (defined as elevated TSH levels and low free T4 concentrations) to 3.47% for subclinical hypothyroidism (defined as elevated TSH levels with normal FT4 concentrations)[3], and 18% for thyroid autoimmunity[2]. Observationally epidemiologic studies have consistently linked abnormal thyroid function tests during pregnancy to adverse maternal and infant outcomes, including preterm birth[4], low birth weight[5], miscarriage, and preeclampsia[6]. However, the underlying biological mechanisms controlling the population variability of TSH, FT4, TPOAb, and thyroid dysfunctions during pregnancy, and their potential causal relationships with pregnancy and long-term health outcomes remain unclear.

Previous research indicates that a significant portion of the inter-individual variability in thyroid hormone levels among general populations can be attributed to genetics. Twin and family studies estimate heritable contributions to approximately 64%-70.8% for serum TSH, 39%-80% for FT4[7–9], and 48.8% for anti-thyroid peroxidase antibody (TPOAb)[9]. Large-scale meta-genome-wide association studies of thyroid hormones among general European populations have identified lead SNPs explaining 22.8% of TSH variation and 4% of FT4 variation, respectively[10,11]. These studies suggest that genetics may also influence thyroid function measurements during gestation, an aspect that has been under-studied. Notably, unlike thyroid functions outside the pregnancy period, changes in the hormonal environment during pregnancy lead to more complex alterations in a woman’s thyroid function. Maternal demand for thyroid rises during pregnancy due to fetal use of thyroid hormones and an increase in thyroxine-binding globulin (TBG), among other reasons[12]. In particular, human chorionic gonadotrophin (hCG), which is structurally similar to TSH, significantly increases in early pregnancy, leading to higher thyroid hormone production, eventually resulting in decreased TSH levels under negative feedback regulation eventually[13]. Therefore, GWAS studies based on pregnant women may not be extrapolated from studies on general adults, and would greatly enhance our understanding of the genetic basis of thyroid-related traits throughout life. However, very few GWAS on thyroid-related traits have included pregnant women. To date, only one candidate gene study that involved 974 healthy pregnant women, reported an association between the number of TSH-raising alleles and subclinical hypothyroidism in pregnancy[14]. In addition, the association between subclinical dysfunction and pregnancy outcomes, as well as the efficacy of treating mild abnormalities, remains uncertain and controversial owing to the lack of robust causal evidence[15,16].

To elucidate the genetic factors influencing thyroid function and disorders during pregnancy, and their potential causal impact on pregnancy complications, birth outcomes, and later-age physical conditions, we conducted a genome-wide association and Mendelian randomization study among 85,421 Chinese pregnant participants with non-invasive prenatal test (NIPT) sequencing data, comprehensive pregnancy screening and medical records from two hospitals in China. We focused on the most commonly used thyroid biomarkers in pregnancy assessments, including thyroid-stimulating hormone (TSH) and free thyroxine (FT4), and thyroid autoimmunity measurements such as anti-thyroid peroxidase antibody (TPOAb) levels and positivity. Additionally, we examined four clinical or subclinical thyroid statuses, including subclinical hypothyroidism (SHO), isolated hypothyroxinemia (ISH), overt hyperthyroidism (OHP), subclinical hyperthyroidism (SHP).

## Results

### Study design and demographic characteristics

Our study design and analysis scheme are depicted in **Figure 1**. Briefly, we enrolled pregnant women during routine pregnancy screening at two maternal and child healthcare hospitals – abbreviated as Longgang and Baoan hospitals, located in Shenzhen, a metropolitan city in southern China. We imputed genotypes and examined population structure from non-invasive prenatal test sequencing data following a standardized protocol developed by our laboratory[17,18]. We conducted GWAS analyses for 2 thyroid-related hormones (TSH and FT4) within reference ranges, 4 thyroid dysfunctions (subclinical hypothyroidism, isolated hypothyroxinemia, subclinical hyperthyroidism, and overt hyperthyroidism), and 2 thyroid autoimmunity traits (TPOAb levels and TPOAb positivity) for each of the two hospitals, followed by a fixed-effect meta-analysis across 12.7 million SNPs (**Methods**). Subsequently, phenotypic association and Mendelian randomization analyses were used to investigate the relationships between these eight traits, gestational complications such as gestational diabetes mellitus (GDM), birth outcomes, and later-life physical conditions using summary statistics from GWAS. Since TSH and FT4 have been investigated in prior GWAS studies in the general population, we further conducted conditional analyses to identify potential novel association signals within the established loci for these two traits. We evaluated the GWAS results by examining the consistency of effect size estimates between the two independent hospitals, the direction of effect sizes with an external GWAS with a smaller sample size (NIPT PLUS cohort, n = 4,688), and by comparing effect size estimates with publicly available GWAS data for the same phenotypes among general populations, if available.

**Figure 1.**
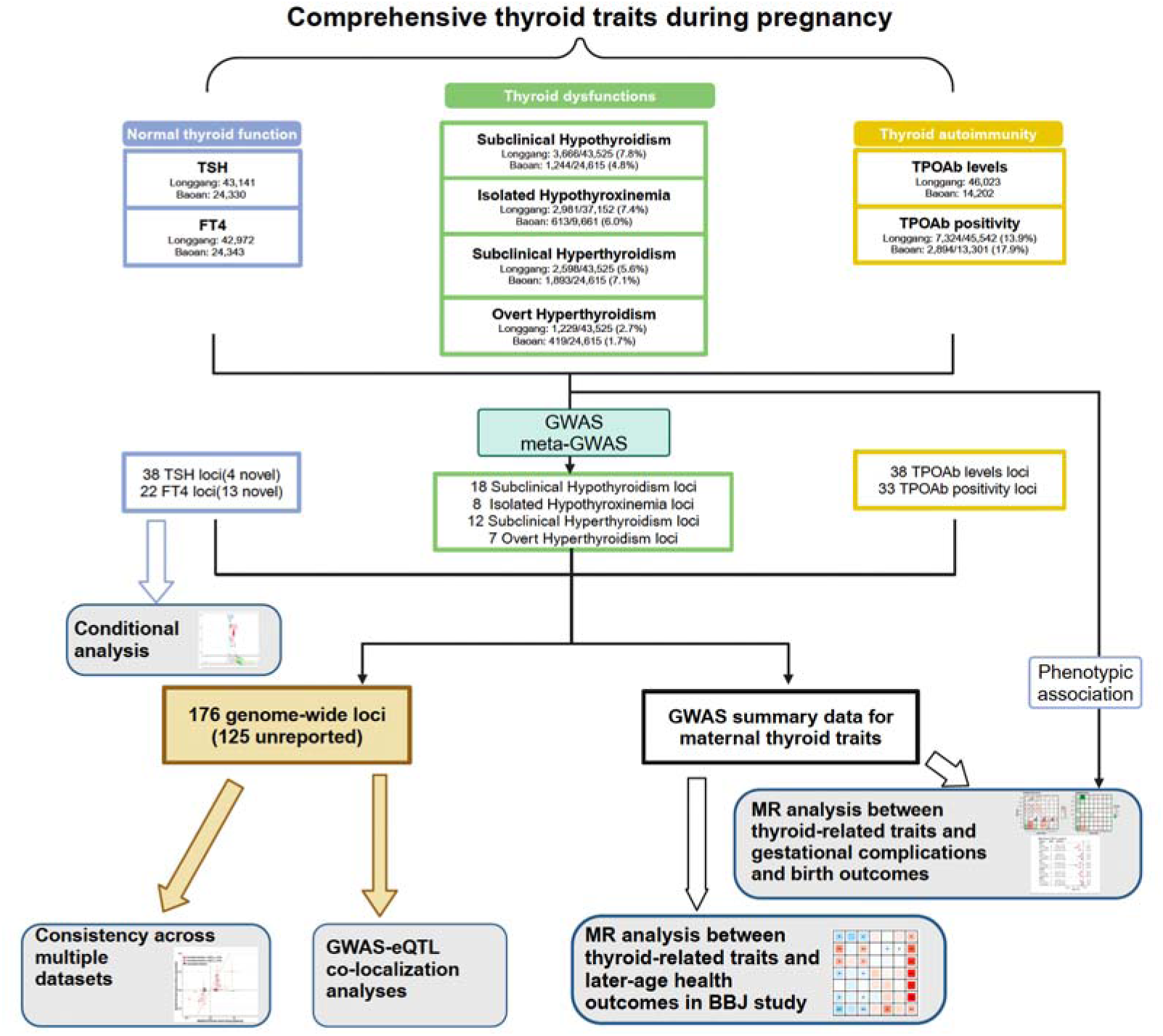
Schematic design of the study. **TSH**: thyroid stimulating hormone; **FT4**: free thyroxine; **TPOAb** thyroid peroxidase antibody. **BBJ**: BioBank Japan Study. Created with BioRender.com.

Following the screening process, our analysis included a total of over 85,000 subjects, amalgamating data from the two hospitals (Methods for inclusion and exclusion criteria). Among them, 67,323 participants had available TSH level data within the reference range, and 67,047 participants had available FT4 level data within the reference range. The sample sizes and demographic characteristics of the research subjects included in the GWAS analysis for each trait, such as TSH levels, FT4 levels, TPOAb levels, age, BMI, and gestational weeks at the time of thyroid function measurement, are detailed in **Table S1**. The majority of subjects were in the age range of 29 to 30 years. As expected, maternal serum TSH concentrations decreased from 8 to 12 weeks and then increased in both cohorts, while the opposite changing pattern was observed for FT4 **(Figure S1)**, corresponding to the increase of hCG during pregnancy[19].

### Genetic associations with normal thyroid function during pregnancy

We identified 38 genome-wide significant loci associated with serum TSH levels and 22 loci associated with serum FT4 levels (*P*-values□≤□5□×□10^−8^) (**Figure 2 and Table S2**). The genomic inflation factor for the 12,699,587 SNPs was 1.068 for TSH and 1.069 for FT4, suggesting negligible inflation of test statistics in the genome-wide association analysis **(Figure S2)**. Among the 60 loci, 4 TSH loci and 13 FT4 loci were identified as novel, with the lead variant situated more than 500kb away and demonstrated LD R^2^ less than 0.1 from any previously established loci **(Table S3; Figure S3).**

**Figure 2.**
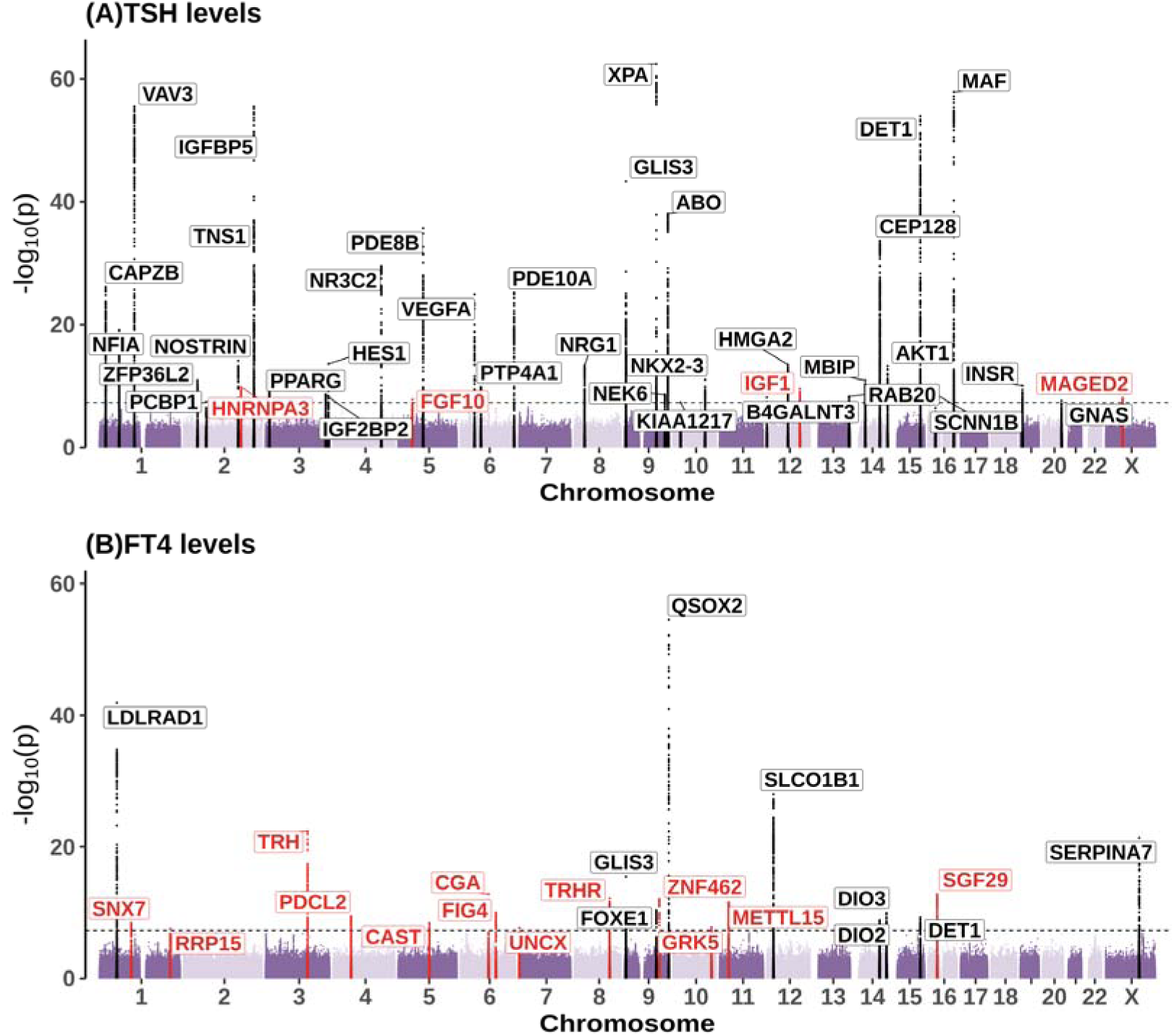
The manhattan plots for GWAS meta-analyses of TSH(A), and FT4(B). The x-coordinate of the SNPs in the plot represents their position on each chromosome when the y-coordinate implies their P-value (-log10 scale) of the association test. The black dotted horizontal line indicates the significant threshold for the genome-wide association test(i.e.,5×10^-8^). Genomic loci containing phenotype-associated variants previously reported in the GWAS catalog are colored in black, while novel loci are colored in red.

Among all loci, 59/60(98.3%) loci demonstrated consistent directionality and had a nominally significant *P*-value below 0.05 between the two hospitals, suggesting little heterogeneity and high fidelity of our genetic discoveries **(Table S2, Figure S4A, Figure S4C).** The remaining *SERPINA7* locus was a known locus associated with FT4 in a previous study and the difference in effect size estimates may be attributed to variations in fetal sex distribution between the two hospitals. In comparison with the GWAS statistics with the NIPT PLUS cohort, 86.7% (52/60) of lead SNPs showed consistent effect directions **(Figure S5, Table S2)**.

Given that available genetic studies on thyroid function among East Asians are limited to small sample sizes in the general population (e.g., 3,618 Koreans[20], 4,581 Chinese [21], and 437 Chinese [22]), with only a few suggestive significant sites identified for TSH, and the lack of publicly available GWAS summary statistics, we compared our effect size estimates with the two largest-scale GWAS studies of TSH and FT4 in European populations (N_TSH_=249,715; N_FT4_=49,269) [10,23]. Among the loci, 49/60 (81.6%) loci exhibited *P*-values below 0.05 with consistent directional effects compared with the European dataset **(Figure S4B, Figure S4D, Table S2)**. While the consistent effect size estimates suggest shared genetic determinants between the two populations (European and East Asian) and between pregnancy and non-pregnancy status, differences in the comparison could be attributed to different LD structures between Europeans and East Asians, as indicated by differing allele frequencies for the lead SNP (**Figure S6**) or potential effect modifications driven by pregnancy status. Further investigation is required to clarify these hypotheses.

### Novel loci and association signals for TSH and FT4

The 4 newly identified loci associated with TSH levels comprise *HNRNPA3, FGF10, IGF1,* and *MAGED2* (**Figure 2A, Table S2, Figure S3**). The 13 novel loci associated with FT4 levels include *SNX7, RRP15, TRH, PDCL2, CAST, CGA, FIG4, UNCX, TRHR, ZNF462, GRK5, METTL15,* and *SGF29* **( Figure 2B, Table S2, Figure S3**).

Potential eGenes which expression in 49 issues in GTEx demonstrates colocalization signals with the 17 loci are presented in **Table S4**. Among the 17 loci, *HMGA2, IGF1, CGA* and *TRHR* contain SNPs that were previously reported to have genome-wide associations with several thyroid-related traits in prior GWAS analyses **(Table S3B**). And some loci include genes directly involved in thyroid functions, such as *TRH* and *TRHR*, as well as genes involved in the regulation of protein signal transduction and cell proliferation (**Table S5**).

Furthermore, through stepwise conditional analyses, we identified that 13 out of the 60 loci contained more than two independent signals, contributing to a total of 33 independent signals (**Table S6, Figure S7**). Particularly, we identified 10 novel significant signals within 10 loci known for their involvement in thyroid function traits [10 TSH-associated loci: *CAPZB, IGFBP5, PDE8B, VEGFA, GLIS3, XPA, MBIP, CEP128, DET1*, and *MAF*; 3 FT4-associated loci: *GLIS3, FOXE1*, and *DET1*] **(Table S7, Methods)**.

Among the 33 independent signals, the genetic effects of all 33 (100%) SNPs were consistent across the two hospitals **(Table S6)**, and 26 out of 33 (78.8%) signals demonstrated consistent direction and significance at *P*=0.05 compared to the two European datasets mentioned earlier[10,23] **(Table S6)**.

Collectively, we identified a total of 53 independent association signals across 38 loci for TSH levels and 27 signals across 22 loci for the FT4 levels, collectively explaining 6.6% and 2.8% of the phenotypic variance, respectively.

### Genetic associations with normal thyroid function within 8-12 gestational weeks

Considering the strong stimulation of thyroid function by hCG, leading to a significant reduction in TSH and an increase in FT4 during early pregnancy (weeks 8 to 12, Figure S1), we conducted a more focused GWAS analysis within this specific timeframe (weeks 8 to 12) to investigate potential association loci related to hCG stimulation. Within the 8 to 12 weeks period, we identified 20 genome-wide significant loci associated with TSH levels and 11 loci associated with FT4 levels **(Figure S8)**. However, after applying the Bonferroni correction, we did not find any loci associated with TSH/FT4 within 8 to 12 weeks that were related to previously published hCG GWAS after Bonferroni correction (**Table S8)**[24]. These findings suggest that the gestational TSH and FT4 level do not share a significant genetic correlation with hCG.

Interestingly, when comparing the genetic effects of the 31 loci within the 8 to12 weeks period with the effects estimated from the entire pregnancy period, we observed that the effect size for 4 out of the 20 TSH loci and 6 out of the 11 FT4 loci did not have overlapping 95% confidence intervals between the two periods **(Figure S9 and Table S9).** Notably, the TSH receptor locus (*TSHR*, marked by the nearest gene *CEP128*) showed stronger associations with TSH and FT4 specifically during the 8 to 12 weeks timeframe of pregnancy. According to GTEx, we found that the lead SNP mutation rs17111346-G, associated with TSH levels during the 8-12 gestational weeks, was associated with a higher TSHR expression level in several tissues and organs **(Figure S10**). In our study, this variant was associated with increased TSH levels and reduced FT4 levels (**Table S9**). These observations may provide a genetic explanation for the occurrence of hypothyroxinemia during this stage, despite hCG stimulation. Individuals carrying specific genetic variants, unrelated to hCG stimulation but associated with lower FT4 and higher TSH during early pregnancy, such as the rs17111346-G variant, may have a higher risk of developing hypothyroxinemia, despite hCG stimulation.

### Identification of genetic associations with thyroid dysfunction

During early pregnancy, the fetus heavily relies on maternal thyroid hormones. Evidence underscores the correlation between untreated clinical thyroid dysfunction and adverse birth outcomes[25,26]. Subclinical hypothyroidism, characterized by low TSH levels but normal FT4 level, and isolated hypothyroxinaemia, characterized by low free T4 levels and normal TSH levels—represent common mild thyroid dysfunctions during pregnancy associated with adverse pediatric outcomes, including preterm birth[4]. Nevertheless, the optimal management of subclinical thyroid dysfunction remains contentious, as some treatments for subclinical hypothyroidism or hypothyroxinemia have not significantly improved cognitive or birth outcomes[26,27]. Moreover, untreated overt hyperthyroidism during pregnancy is linked to various maternal and fetal complications, such as pre-eclampsia and pregnancy loss, while subclinical hyperthyroidism is not thought to be associated with adverse birth outcomes[28].

To elucidate the genetic underpinnings of these thyroid dysfunctions, we conducted a GWAS analysis on four categorical traits: subclinical hypothyroidism, isolated hypothyroxinemia, overt hyperthyroidism, and subclinical hyperthyroidism (cases). We utilized pregnant women with euthyroid function, defined as those maintaining TSH and FT4 levels within the pregnancy-specific reference range throughout pregnancy, as the control group. Notably, we also examined clinical hypothyroidism; however, the statistical power was limited due to the small number of cases (N_Longgang_=464, N_Baoan_=148) and robust genetic associations were not identified. Specifically, we excluded individuals who tested positive for TPOAb from both the case and control populations in the analysis for isolated hypothyroxinemia. After exclusions, the sample sizes of the included research subjects were as follows: (1) subclinical hypothyroidism [N=73,050 (Longgang: 3,666 cases versus 43,525 controls; Longang: 1,244 cases versus 24,615 controls)]; (2) isolated hypothyroxinemia: [N=50,407 (Longgang: 2,981 cases versus 37,152 controls; Longang: 613 cases versus 9,961 controls)]; (3) subclinical hyperthyroidism [N=72,631(Longgang: 2,598 cases versus 43,525 controls; Longang: 1,893 cases versus 24,615 controls)]; (4) overt hyperthyroidism [N=69,788 (Longgang: 1,229 cases versus 43,525 controls; Longang: 419 cases versus 24,615 controls)].

Our analysis led to the identification of 18, 8, 12, and 7 genome-wide association loci associated with maternal subclinical hypothyroidism, isolated hypothyroxinemia, subclinical hyperthyroidism, and overt hyperthyroidism, respectively (**Figure 3, Table S10, Figure S11**). eGenes with colocalization signals with these 45 loci are presented in **Table S4**. All lead SNPs in these 45 loci exhibited consistent directionality within two hospitals, with 42 out of 45 (93.3%) compared at a *P-value* below 0.05 with consistent directionality **(Figure S12A)**. When we replicated these 45 lead SNPs with the NIPT PLUS cohort, 77.8% (35/45) SNPs showed consistent directions of genetic effects **(Figure S12B)**.

**Figure 3.**
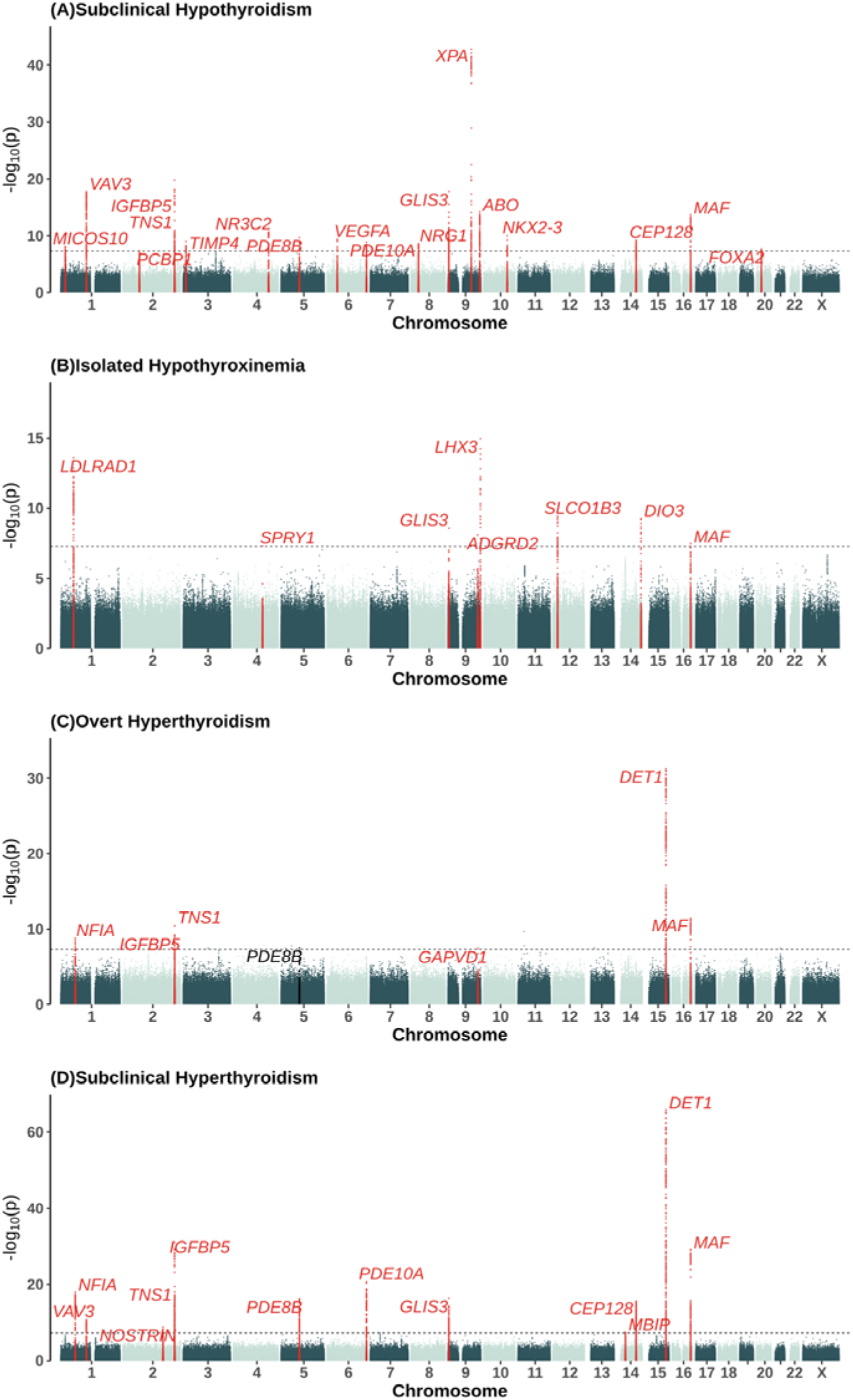
The manhattan plots for GWAS meta-analyses of thyroid dysfunction: subclinical hypothyroidism**(A)**, isolated hypothyroxinemia **(B)**, overt hyperthyroidism**(C)**, and subclinical hyperthyroidism**(D)**. The x-coordinate of the SNPs in the plot represents their position on each chromosome when the y-coordinate implies their P-value (-log10 scale) of the association test. The black dotted horizontal line indicates the significant threshold for the genome-wide association test(i.e.,5×10-8). Genomic loci containing phenotype-associated variants previously reported in the GWAS catalog are colored in black, while novel loci are colored in red.

As no previous GWAS studies have been conducted on the four thyroid dysfunctions investigated in this study, we were unable to directly compare the outcome with external datasets. However, we noted that most of these loci have been reported to be associated with thyroid-related traits such as hypothyroidism, TSH, and FT4 **(Table S3B)**. After excluding loci previously associated with thyroid traits, we identified 2 novel low-frequency genome-wide thyroid-traits loci including isolated hypothyroxinemia-related *SPRY1* (rs190595112-G, MAF=0.012, OR[95%]:4.22[2.52-7.07], P= 4.77E-08) and overt hyperthyroidism-related *GAPVD1* (rs78254323-T, MAF=0.04, OR[95%]: 1.72[1.42-2.09], P= 3.94E-08). These two variants demonstrated consistent directionality and significance between the two hospital cohorts (**Table S10**). The *GAPVD1* locus demonstrated certain levels of evidence of associations with GAPVD1 expression in the thyroid, while we did not find eGene for *SRY1* locus in all tissues (**Table S4**).

### Genetic associations with thyroid autoimmunity

Thyroid autoimmunity, characterized by the presence of antibodies targeting thyroperoxidase, thyroglobulin, and thyroid-stimulating hormone receptor antibodies (TRAbs), is prevalent among pregnant women and is often diagnosed through the detection of thyroperoxidase antibodies (TPOAb) in clinical settings. This condition is associated with thyroid dysfunction and adverse obstetric outcomes[29]. In this study, we conducted genome-wide association studies (GWAS) focusing on TPOAb quantitative levels and positivity during pregnancy (**Methods**). The sample sizes of the included research subjects were as follows (1) quantitative TPOAb levels [N= 60,225 (Longgang: 46,023; Longang: 14,202)]; (2) TPOAb-positivity [N= 69,061(Longgang: 7,324 cases versus 45,542 controls; Longang: 2,894 cases versus 13,301 controls)].

We identified 38 and 33 loci associated with TPOAb levels and TPOAb positivity, respectively (**Figure 4, Figure S13 and Table S11**), revealing 35 and 30 newly identified association loci, respectively compared to previously reported signals in the GWAS catalog. Most of the loci for both traits were shared. The four most significant novel loci including *VANGL2, CTLA4, GPX6*, and *GPR174*, were identified as associated with Graves’ disease (GD)/Hashimoto’s thyroiditis (HT) in the Japanese population[30] (**Table S3B**). We identified 4 missense variants in novel loci: rs7522061-C in *FCRL3*, rs3775291-T in *TLR3*, rs12793348-G in *PANX1* and rs229527-C in *C1QTNF6.* GWAS-eQTL co-localization analyses identified several TPOAb levels and positivity-related loci that regulate the expression of *RPS26, RNASET2, C1QTNF6, FCRL1, FCRL3* genes in multiple tissues (**Table S4**). Previous study has reported that *RNASET2* is expressed in CD4 + T-helper and CD8 + T cells[31], while *FCRL3* is expressed at high levels during the maturation of B-cells and is thought to regulate B-cell signaling in both positive and negative ways[32]. *FCRL1* also plays a role in the assembly of the BCR signalosome, influences B cell signaling, and enhances humoral responses[33].

**Figure 4.**
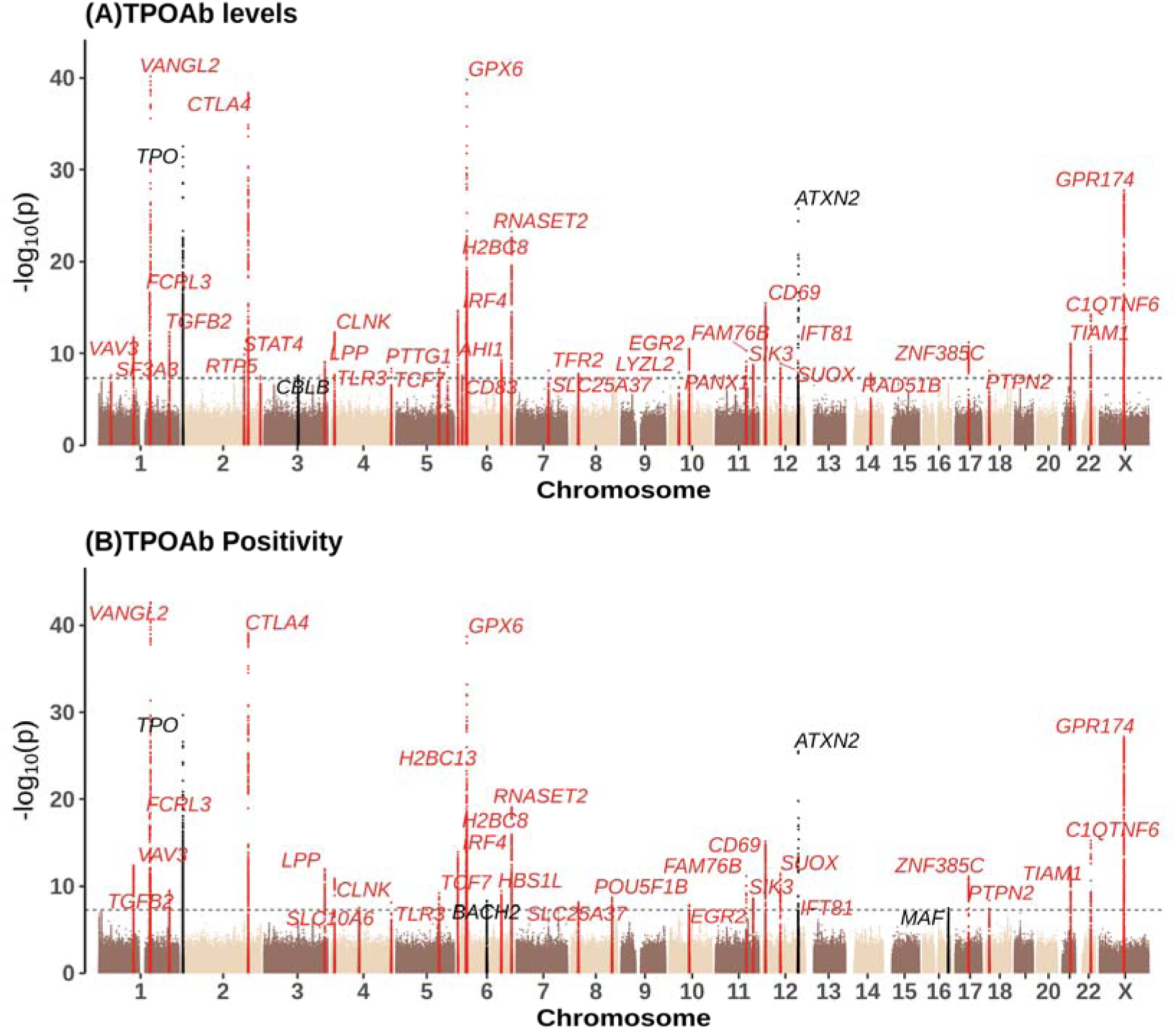
The manhattan plots for GWAS meta-analyses of thyroid autoimmunity: TPOAb levels **(A)**, TPOAb positivity **(B).** The x-coordinate of the SNPs in the plot represents their position on each chromosome when the y-coordinate implies their P-value (-log10 scale) of the association test. The black dotted horizontal line indicates the significant threshold for the genome-wide association test(i.e.,5×10-8). Genomic loci containing phenotype-associated variants previously reported in the GWAS catalog are colored in black. Before performing the GWAS analysis, TPOAb levels were natural log-transformed.

For consistency, 67 out of 71 (94.4%) loci demonstrated consistent direction and significance at *P*=0.05 between the two hospitals (**Figure S14A, Table S11**). When replicating GWAS for TPOAb levels and positivity in the NIPT PLUS cohort, 62 out of 71 (87.3%) loci showed consistent directions of genetic effects (**Figure S14B, Table S11**). Comparing the results with external datasets, we initially compared our results with the largest previous GWAS study on TPOAb levels and positivity, conducted by Marco Medici et al. in the European population in 2014 (N_TPOAb-levels_=16,528 and N_TPOAb-positivity_=18,297)[34]. Since the study involved 2.14 million variants, only 58 of our lead variants (for their proxy SNPs) were available for analysis in this study. Among these SNPs, 50 loci (86.2%) exhibited a consistent direction of effect between Chinese pregnant women and the European population **(Figure S15, Table S12)**.

Given the unavailability of previous GWAS summary data for TPOAb in the East Asian population and the high correlation between TPOAb positivity and autoimmune thyroid diseases such as Graves’ disease and Hashimoto’s thyroiditis[35], we also examined our associations for TPOAb positivity using the GWAS results for Graves’ disease and Hashimoto’s thyroiditis from the Biobank Japan (BBJ)[30]. Of these 33 loci, *H2BC8* (rs1051365311) and its proxy SNP were not available in the BBJ dataset. According to the result, 15 loci (*FCRL3, VANGL2, CTLA4, LPP, CLNK, TCF7, IRF4, GPX6, HBS1L, RNASET2, FAM76B, IFT81, MAF, C1QTNF6,* and *GPR174*) in Grave’s disease **(Table S13, Figure S16A)** and 2 loci (*GPX6, VANGL2*) in Hashimoto’s thyroiditis showed significant associations **(Table S13, Figure S16B).** The high degree of concordance (32 out of 33) in the direction of locus effects suggests a shared genetic basis between TPOAb positivity during pregnancy and autoimmune thyroid diseases.

### Pairwise genetic correlation among thyroid traits

To explore the pairwise genetic correlation among the thyroid traits investigated, we employed LD score regression[36]. The analysis revealed a significant genetic correlation between TSH and all other thyroid traits, including TPOAb levels and positivity. Conversely, FT4 exhibited correlation with thyroid dysfunctions but not with TPOAb levels and positivity **(Figure S17, Table S14)**. Additionally, we observed a correlation between TPOAb level/positivity and subclinical hypothyroidism. Nonetheless, this genetic correlation lost significance when the analysis was restricted to euthyroid individuals, suggesting that the observed correlation stemmed from shared genetics between TPOAb and TSH.

Consistent with the established inverse correlation in TSH and FT4 regulation through the Hypothalamus-Pituitary-Thyroid Axis (HPT) [37], we observed a significant inverse genetic correlation between TSH and FT4 levels (the genetic correlation calculated by LDSC: rg_TSH-FT4_=-0.16, se=0.04, *P*=8.6E-5)**(Figure S17, Table S14)**, along with shared associations at 11 loci (TSH: *CAPZB, TNS1, GLIS3, XPA, MBIP, DET1, MAF,* and FT4: *FIG4, GLIS3, FOXE1, DET1*) (Bonferroni-corrected threshold p□<□0.05/60) (**Figure S18, Table S15**). At these 11 loci, alleles associated with higher TSH levels were consistently associated with lower FT4 except for three loci-*FIG4, XPA*, and *FOXE1* – where genetic effects were observed in the same direction for both thyroid-related hormones.

### The influence of thyroid-related traits on gestational complications and fetal obstetric outcomes

Previous large meta-analyses of observational studies have demonstrated an association between subclinical thyroid dysfunction and maternal complications and adverse birth outcomes, including gestational diabetes mellitus (GDM)[38], hypertensive disorders[39], miscarriage [40], preterm birth, and low birth weight[4,5]. However, RCTs investigating treatments for mild thyroid function abnormalities have yielded inconclusive results [27], resulting in uncertainty and controversy regarding the relationship between subclinical dysfunction and pregnancy outcomes, as well as the therapeutic value of addressing mild abnormalities[41,42]. High-quality Mendelian randomization studies in clinical thyroidology are deemed crucial for gaining valuable insights into the relationship between thyroid functions and birth outcomes[43]. However, to date, only one MR study has explored the causal effect of TSH and FT4 levels within the normal range on birth weight and did not identify statistically significant causal associations [44].

Here, we applied joint phenotypic association and Mendelian randomization to investigate the impact of thyroid function during pregnancy on gestational complications such as hypertension, and gestational diabetes, as well as birth outcomes including birth weight, birth length, and gestational duration (**Table S18**). Adhering to the three preconditions for a robust MR analysis, we utilize strong instrumental variables (F > 10), accounted for heterogeneity and horizontal pleiotropy by applying an inverse weighted approach of random effect (IVWRE) as primary results, and conducted several sensitivity analyses, including six additional two-sample MR approaches and three scenarios including overlapping (Meta-Meta) and non-overlapping samples (Longgang-Bao and Baoan-Longgang)[45]. In addition, for MR analysis of the relationship between thyroid traits and pregnancy complications, we accounted for the bi-directionality of the causal estimates (**Method)**. Notably, due to the extreme imbalance in the number of cases and controls of gestational hypertension and preeclampsia, we employed quantitative systolic and diastolic blood pressure as indicators for hypertension. Considering the substantial genetic correlation among the gestational thyroid traits (**Figure S17)**, we employed a Bonferroni testing criteria based on the total number of investigated gestational complication and fetal obstetric outcomes (N=14). The statistical results of all methods are presented in **Table S18 and S20**, while the instrumental variables used are listed in **Table S19**.

Phenotypic association tests revealed numerous strong associations between TSH and FT4 within the normal range, thyroid dysfunction, and glucose and blood pressure traits, including significant associations between ISH and GDM (P < 0.004) (**Figure 5A, Table S17**). Mendelian randomization suggested consistent potential causal associations for a few of these correlations after Bonferroni correction, including a significant negative effect of TSH on fasting plasma glucose (FPG), oral glucose tolerance at 1 hour (OGTT1H), and systolic blood pressure (SBP), as well as a significant effect of FT4, SHO, SHP on SBP (**Figure 5B-C, Table S18,** P < 0.004). Additionally, we found a nominally significant protective effect of genetically increased thyroid hormone (FT4) on GDM, consistent between MR and observational studies (observational study: OR_longgang_ [95%CI]: 0.861[0.833∼0.89], P_longgang_ =8.49E-19; MR: OR[95%CI]:0.844[0.732∼0.972], P=1.88E-02). Reverse Mendelian randomization did not identify significant potential causal associations between the pregnancy phenotypes and thyroid-related phenotypes after Bonferroni correction (**Figure 5B, Table S20**).

**Figure 5.**
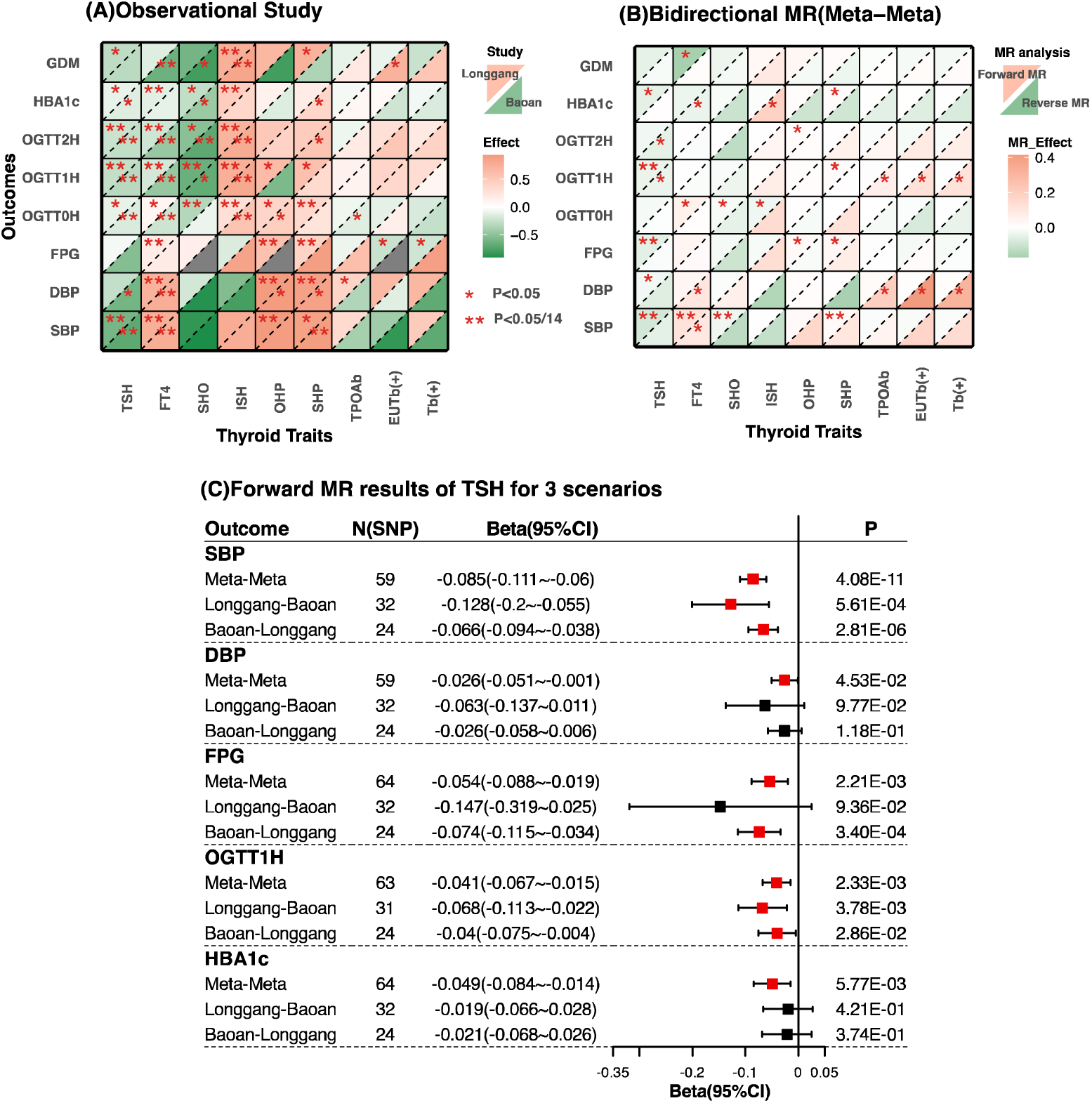
The association between thyroid-related traits and maternal blood glucose and blood pressure traits during pregnancy: (A)Observational analysis. (B) Bidirectional mendelian randomization. (C)The forest plot of MR results under 3 scenarios between TSH and 5 blood glucose and blood pressure traits. In Figure A, the upper left triangle of each grid indicates the phenotypic association results in the Longgang cohort, while the lower right triangle indicates the phenotypic association results in the Baoan cohort. To facilitate the comparison of effect estimates corresponding to different traits, we converted the absolute values of the estimates of phenotypic associations into Q-quantiles for plotting. Shaded areas indicate that the results of the associations between FPG and each of SHO, OHP, and EUTb(+) are unavailable due to an insufficient number of subjects in Baoan hospital. In Figure B, the upper left triangle of each grid indicates the results of forward MR (thyroid traits as exposure variables), while the lower right triangle indicates reverse MR (thyroid traits as outcome variables). In Figure C, the MR results for three scenarios are shown: (1) Meta-Meta: the two-sample MR method is conducted in one-sample setting; (2) Baon-Longgang: Baoan Study served as the source of exposed GWAS data, while the Longgang Study provided the outcome population. (3) Longgang-Baoan: Longgang Study served as the source of exposed GWAS data, while the Baoan Study provided the outcome population. TSH: TSH levels within reference range during pregnancy; FT4: FT4 levels within reference range during pregnancy; SHO: subclinical hypothyroidism during pregnancy; ISH: isolated hypothyroxinemia during pregnancy, OHP: overt hyperthyroidism during pregnancy; SHP: subclinical hyperthyroidism during pregnancy, TPOAb: TPOAb levels, Tb(+): TPOAb positivity during pregnancy and EUTb(+): TPOAb positivity with normal functioning thyroids(ie, euthyroid). SBP: systolic blood pressure; DBP: diastolic blood pressure, HBA1c, Glycated hemoglobin; FPG, Fasting plasma glucose, OGTT0H, Oral glucose tolerance test 0 hour; OGTT1H, Oral glucose tolerance test 1 hour; OGTT2H, Oral glucose tolerance test 2 hour, GDM, Gestational diabetes mellitus.

As for birth outcomes, observational analysis in the Longgang and Baoan study consistently showed that maternal FT4 levels within the reference range were significantly associated with lower birth weight, shorter birth length and a higher risk of low birth weight. Correspondingly, we also observed a consistently significant positive correlation between isolated hypothyroxinemia during pregnancy and greater birth weight, as well as a significant association between TPOAb levels and decreasing birth weight (**Figure 6A, Table S17**). However, in MR analysis, we only discovered a significant potential causal effect of elevated TSH levels during pregnancy on greater gestational duration (days) after Bonferroni correction (β_MR-IVWRE_ 95%CI =0.07[0.04-0.11], P_MR-IVWRE_=7.80E-5) **( Figure 6B, Table S18)**. Similar results were observed across other methods of Mendelian randomization, except for simple mode (SM), which showed attenuation toward the null. We did not identify MR associations between thyroid dysfunction and birth outcomes (**Figure 6B, Table S18**).

**Figure 6.**
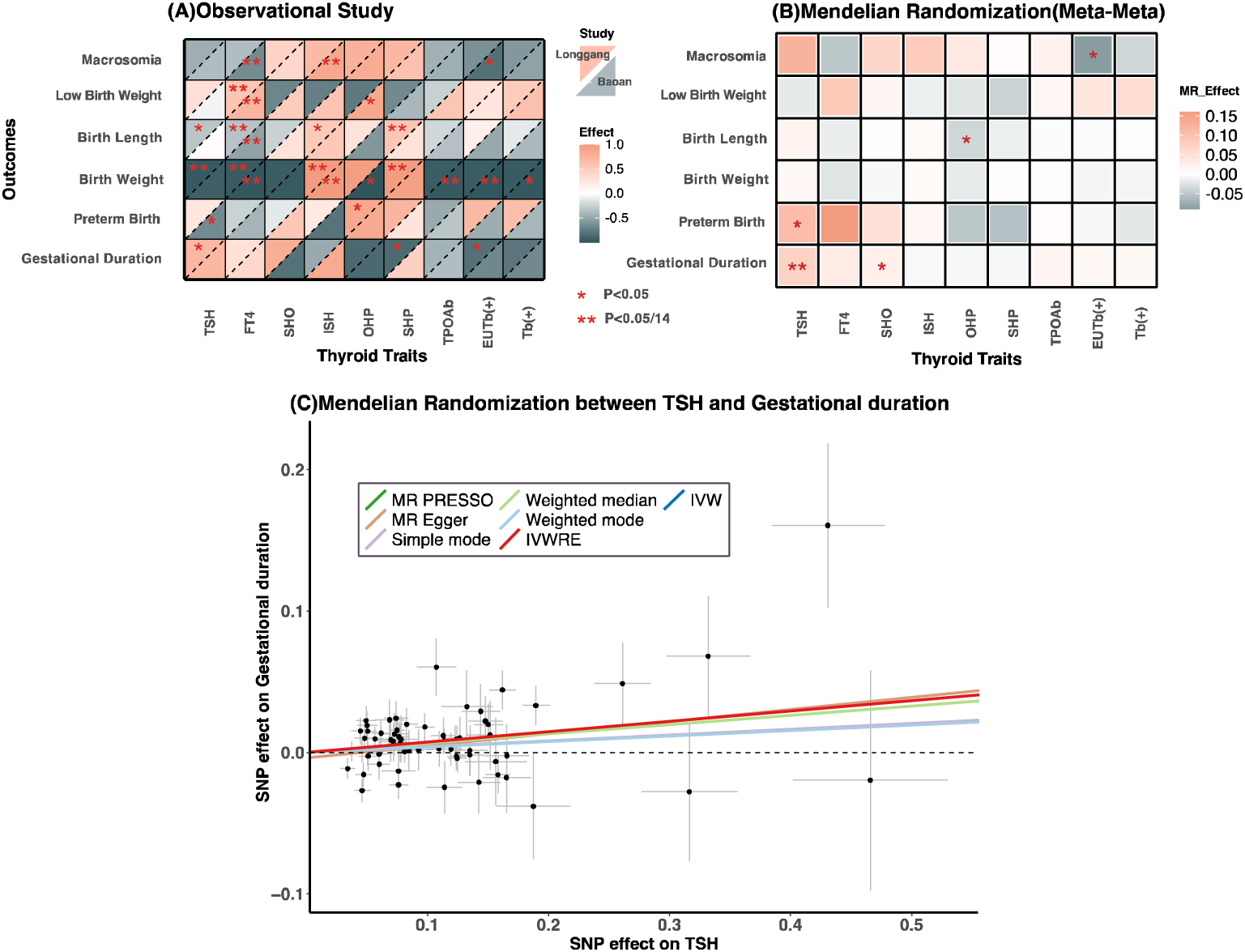
The association between thyroid-related traits during pregnancy and birth outcomes: (A)Prospective observational analysis. (B) Mendelian randomization. (C)Mendelian randomization analysis for casual associations between TSH and thyroid cancer and between TSH and gestational duration. In Figure A, the upper left triangle of each grid indicates the phenotypic association results in the Longgang cohort, while the lower right triangle indicates the phenotypic association results in the Baoan cohort. In Figure C, TSH and gestational duration are based on summary statistics from the meta-analysis of the Baoan and Longgang Study. (N_TSH_=□67,471, and N_gestational duration_ =□51,592). The results of the IVW, IVWRE, MR Egger, Simple mode, and Weighted mode methods are based on the analysis of the IVs excluded by MR PRESSO. The crosshairs on the plots represent the 95% confidence intervals for each SNP-TSH or SNP-outcome association. IVW, Inverse Variance Weighted; IVWRE, Inverse Variance Weighted with Multiplicative Random Effects. TSH: TSH levels within reference range during pregnancy; FT4: FT4 levels within reference range during pregnancy; SHO: subclinical hypothyroidism during pregnancy; ISH: isolated hypothyroxinemia during pregnancy, OHP: overt hyperthyroidism during pregnancy; SHP: subclinical hyperthyroidism during pregnancy, TPOAb: TPOAb levels; Tb(+): TPOAb positivity during pregnancy and EUTb(+):TPOAb positivity with normal functioning thyroids(ie, euthyroid).

### Bidirectional Mendelian randomization study between gestational thyroid traits and 220 phenotypes in BioBank Japan

We further investigated the potential relationship between thyroid-related traits during pregnancy and 220 health phenotypes and outcomes in Biobank Japan (BBJ), with an average age of 62, by conducting a bidirectional Mendelian randomization analysis (**Methods**). Consistent significant associations in both directions suggest genetic sharing rather than causal relation. The Bonferroni testing criteria are based on the total number of investigated BBJ phenotypes (N=220), considering the substantial genetic correlation among the gestational thyroid traits (**Figure S17)**. Results surpassing the Bonferroni and MR Egger pleiotropy testing criteria, which mainly pertain to BBJ thyroid and cardiac disorders, are discussed herein. The instrumental variables used in the forward MR analysis are presented in **Table S22**.

Regarding thyroid autoimmunity, forward MR analysis suggested that genetically determined elevated susceptibility to TPOAb positivity is associated with an increased risk for all 7 thyroid disorders, with 6 of these associations remaining significant after Bonferroni correction: Graves’ disease (OR [95%CI]:1.55[1.41∼1.704], P=9.68E-20), thyroid preparations usage (1.364[1.259∼1.478], P=2.66E-14), hypothyroidism (1.441[1.297∼1.6], P=9.10E-12), hyperthyroidism(1.478[1.318∼1.658], P=2.37E-11), Hashimoto thyroiditis(1.454[1.27∼1.666], P=6.61E-08), and goiter (1.163[1.077∼1.256], P=1.12E-04) (**Figure 7**, **Figure S19A∼F, Table S21)**. Similar observations apply to TPOAb levels and for euthyroid TPOAb positivity with normal TSH and FT4 levels **(Figure S19G∼N)**. However, in the reverse MR, statistics are unavailable for thyroid cancer, hypothyroidism, Hashimoto thyroiditis and goiter due to unavailability of instrumental variables. Significant bidirectionality was observed for Graves’ disease and thyroid preparations, and same effect direction was observed for hyperthyroidism (**Figure 7**). Therefore, we infer that TPOAb positivity and levels during pregnancy are genetically correlated with the BBJ thyroid disorders. In addition to thyroid-related diseases, forward MR also revealed that genetic TPOAb positivity is associated with a reduced risk of depression (OR [95%CI]: OR[95%CI]:0.848[0.793∼0.907], P=1.60E-06) and an increased risk of Cardiac valvular disease (OR [95%CI]: OR[95%CI]:1.109[1.052∼1.169], P=1.24E-04) **(Figure S19O∼P)**. Reverse MR is unavailable for both disorders due to a lack of IVs (**Figure 7, Table S23)**.

**Figure 7.**
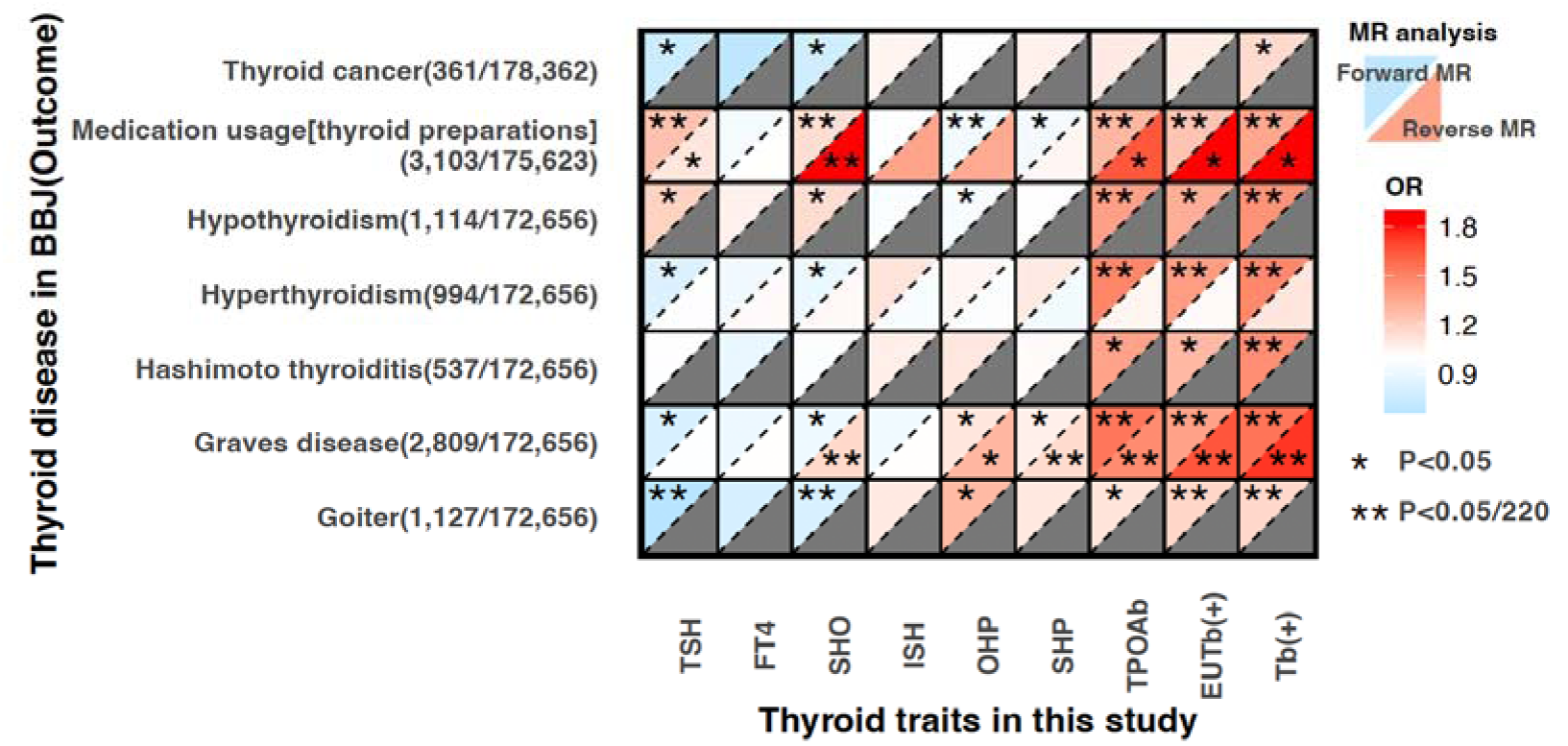
The results of bidirectional mendelian randomization analysis between thyroid-related traits during pregnancy and thyroid-related diseases in middle-aged and elderly people. The upper left triangle of each grid indicates the results of forward MR (thyroid traits as exposure variables), while the lower right triangle indicates the results of reverse MR (thyroid traits as outcome variables). Shaded areas indicate that reverse MR results with goiter, Hashimoto’s thyroiditis, hypothyroidism and thyroid cancer as the exposure variable are not available due to insufficient number of IVs. TSH: TSH levels within reference range during pregnancy; FT4: FT4 levels within reference range during pregnancy; SHO: subclinical hypothyroidism during pregnancy; ISH: isolated hypothyroxinemia during pregnancy, OHP: overt hyperthyroidism during pregnancy; SHP: subclinical hyperthyroidism during pregnancy, TPOAb: TPOAb levels, Tb(+): TPOAb positivity during pregnancy and EUTb(+): TPOAb positivity with normal functioning thyroids (ie, euthyroid).

In the forward MR analysis of both normal TSH, FT4 and thyroid dysfunction, we observed that genetically elevated TSH levels, and a high risk of subclinical hypothyroidism were linked to an increased usage of thyroid preparations, while overt hyperthyroidism was linked to a decreased usage of thyroid preparations (*P* < 1.72E-04) (**Figure 7, Figure S20A∼C and Table S21**). Except for overt hyperthyroidism, TSH and subclinical hypothyroidism suggest bi-directionality of the MR effect. In addition, genetically elevated TSH (OR [95%CI]:0.66[0.55-0.79], P=1.12E-05) and subclinical hypothyroidism (OR[95%CI]:0.81[0.74-0.89], P=8.54E-06) were associated with a lower risk of goiter (**Figure S20D-E**). However, reverse MR is unavailable for both disorders due to a lack of IVs (**Figure 7, Table S23**). Moreover, we found genetically elevated TSH levels but not thyroid dysfunction are associated with a reduced risk of atrial fibrillation (OR [95%CI]: 0.868[0.81∼0.931], P=7.91E-05) **(Figure S20F)**, which did not demonstrate evidence of causality in the reverse MR, suggesting potential causal relation.

## Discussion

Utilizing hospital electronic medical records and NIPT data from 85,421 Chinese pregnancies, we conducted the first and most comprehensive GWAS studies to date of thyroid traits during pregnancy, encompassing thyroid-related hormones, dysfunction, and autoimmunity. Simultaneously, our GWAS facilitates the exploration of thyroid function’s impact on gestational complications as well as on birth outcomes, utilizing the methods of Mendelian randomization.

In summary, we identified 176 genome-wide loci, among which 125 genome-wide associations had not been previously reported for the 8 thyroid-related traits. This significantly enhances our understanding of the genetic basis of thyroid traits during pregnancy. The high consistency of genetic effects observed across two independent hospitals, and in comparison with external datasets demonstrates the robustness of our GWAS results. Particularly, while the *TRH* and *TRHR* genes are well-established in the thyroid axis[37], our study is the first to identify them as genome-wide association loci for thyroid function (FT4), signifying a significant power gain in our analysis. Moreover, we identified 33 independent association signals in 13 loci (TSH: *CAPZB, IGFBP5, PDE8B, VEGFA, GLIS3, XPA, MBIP, CEP128, DET1*, and *MAF*; FT4: *GLIS3*, *FOXE1*, and *DET1*) by the stepwise conditional analysis, suggesting allelic heterogeneity at the thyroid-associated loci. Additionally, comparing the genetic associations with normal TSH and FT4 within 8-12 gestations suggests that individuals carrying specific genetic variants have a higher risk of hypothyroxinemia, despite hCG stimulation.

In the first GWAS of gestational thyroid dysfunction, we identified 18, 8, 12 and 7 genome-wide loci associated with subclinical hypothyroidism, isolated hypothyroxinemia, and subclinical hyperthyroidism, and overt hyperthyroidism during pregnancy, respectively. Additionally, our study has doubled the number of genome-wide loci associated with TPOAb levels and positivity. These loci also exhibited connections with other thyroid-related traits and conditions, including hypothyroidism, TSH levels, and the use of thyroid preparations. Further functional and mechanistic studies may be warranted to explore potential new therapeutic targets.

We completed the first joint epidemiological and Mendelian randomization analysis of gestational thyroid phenotypes and complications. Statistics suggest that genetically predicted lower TSH and higher FT4 correlate with elevated blood glucose levels. However, we did not identify a significant MR association between thyroid dysfunction and GDM, implying a continuum of TSH and FT4 action on blood glucose and blood pressure regulation. Previous studies have yielded conflicting conclusions regarding the association between subclinical hypothyroidism and GDM[46–48], and our results suggest that the impact of abnormal thyroid function on GDM varies depending on the criteria used for TSH and blood glucose assessment. Moreover, our results suggest the involvement of TSH and FT4 in blood pressure regulation. While a previous MR analysis suggested that genetically predicted TSH elevation leads to reduced systolic blood pressure, this finding lacked significance after Bonferroni correction [49]. Our study provides more robust evidence to support this conclusion.

In the investigation of relationship of gestational thyroid phenotypes and birth outcomes, retrospective observational analyses revealed that isolated hypothyroxinemia was associated with higher birth weight, while FT4 levels within the reference range were inversely associated with birth weight, aligning with findings from a previous large meta-analysis[5]. However, we did not find sufficient evidence to suggest that abnormal TSH levels during pregnancy (overt and subclinical hyperthyroidism, subclinical hypothyroidism) and TPOAb positivity increase the risk of adverse outcomes such as preterm delivery, macrosomia, and low birth weights. Nonetheless, we cannot exclude the possibility that thyroid dysfunction and autoimmunity may contribute to other adverse outcomes. Consistent with a previous MR study[44], we also did not identify an MR association between maternal TSH levels within the reference range and birth weight. Notably, our MR analyses provide evidence suggesting that maternal TSH levels during pregnancy may potentially influence gestational duration, although the effect may be nonlinear. Notably, genetically predicted higher maternal TSH are nominally associated with a heightened risk of preterm birth, and previous research has demonstrated a U-shaped association between FT4 levels and preterm birth[50]. Further validation through intervention studies is warranted to ascertain the genuine effects.

TPOAb is commonly acknowledged as a pathogenic antibody associated with Hashimoto’s thyroiditis (HT), yet its role in Graves’ disease (GD) remains uncertain, despite the fact that 50–70% of individuals with GD also exhibit TPOAb positivity[51,52]. TPOAb has been proposed as a marker of impaired thyroid function rather than a direct cause of thyroid damage[2]. Our bidirectional MR analyses with BBJ phenotypes suggest a genetic correlation between TPOAb positivity and several thyroid disorders in older, general individuals, regardless of their thyroid function status. Additionally, we observed that genetically predicted maternal TPOAb positivity was associated with an increased risk of cardiac valvular disease, consistent with previous observations of an association between thyroid autoimmunity and rheumatic heart valve disease[53]. In addition, in line with the findings of previous MR studies conducted in population of European ancestry[23,54], identified protective effects of lower thyroid function during pregnancy against atrial fibrillation and goiter in East Asians. The consistently observed genetic correlation or MR associations of gestational thyroid phenotypes and later-age thyroid and cardiac diseases suggest the potential for early screening of thyroid disorders in early adulthood and may warrant further clinical studies.

Despite the positive findings, there are limitations in our study. Firstly, while we conducted the first and most comprehensive GWAS study of thyroid-related traits in pregnancy, focusing on a critical period of human reproduction, the study population consisted solely of Chinese women, potentially limiting the generalizability of our findings to other ancestries. However, we would like to note that non-invasive prenatal testing data are widely adopted in clinical settings due to their high sensitivity and specificity[55]. The methods used in this study[17,18] will facilitate further investigations into thyroid-related traits in pregnancy across different populations, enhancing our understanding of the trans-ancestral genetic basis. In addition, the shared genetic correlation between the gestational thyroid traits in this study and the thyroid phenotypes in European general population suggests that the existance of strong correlation between thyroid traits in different life stages.

Secondly, in our Mendelian randomization analysis examining the relationship between gestational thyroid traits and complications as well as birth outcomes, we utilized a strategy involving two-sample MR analytical methods within a single cohort setting, which has been empirically proved not to be prone to sample-overlapping bias if the IVs were sufficiently strong[45]. Nonetheless, to enhance the robustness of our findings, replication in a two-sample MR without sample overlap would be ideal. We attempted to address this by conducting analyses in multiple scenarios, including using exposures from one hospital and outcomes from the other hospitals, to support our MR results. These strategies consistently suggested that genetically determined elevated TSH levels during pregnancy were associated with lower glycemic levels, reduced blood pressure, and longer gestational duration. Furthermore, in our MR study of gestational thyroid traits and birth outcomes, we did not differentiate between fetal genetic and intrauterine effects of exposure on birth outcomes. This aspect could benefit from expansion of sample size in family genomes from birth cohorts in Chinese ancestry[56]. Additionally, when analyzing the impact of thyroid traits on other outcomes, we focused solely on additive linear effects, without considering nonlinear effects. Finally, in the bidirectional MR study between gestational thyroid traits and phenotypes in BBJ, we were unable to distinguish genetic correlation from potential causal effect of the exposures of thyroid related traits during pregnancy on a later-age health result. This purpose can be addressed when GWAS studies of the same thyroid traits investigated in this study among general East Asian population become available, enabling a multi-variable Mendelian randomization analysis integrating GWAS statistics of thyroid traits in pregnancy and among the general populations[57].

Despite the genetic findings, our study suggests two clinical implications. First, serum levels of TSH and FT4 exhibit substantial interindividual variation among healthy individuals, with much smaller intraindividual variation[58]. This indicates that each person has a unique setpoint for TSH and FT4[59]. Hence, when initiating RCTs or treatments for thyroid disorders in pregnant women, it may be beneficial to consider the genomic profile of the patients, as revealed in our study. This approach can help prevent overtreatment, may explain the varying responses of the pregnant women to medications such as levothyroxine, commonly prescribed for gestational thyroid disorders[27]. Secondly, as pregnancy screening represents the most comprehensive physical examination during early adulthood for most women, it is valuable to explore early diagnosis and preventive strategies for thyroid disorders later in life, starting from this critical early period.

## Methods

### Longgang and Baoan Study

The Longgang Study enrolled 70,608 pregnant women who received NIPT results during the first or second trimester at Longgang District Maternal and Child Health Hospital, located in the eastern region of Shenzhen, China, between 2017 and 2022. Of these, 58,351 participants had available measurements for TSH, FT4, and TPOAb levels.

The Baoan Study included 50,948 pregnant women who visited the Baoan District Maternal and Child Health Hospital in the western region of Shenzhen, China, and completed an NIPT test during the first or second trimester between 2017 and 2022. Of these, 33,781 participants had available TSH measurements, 33,516 had FT4 measurements, and 16,393 had TPOAb measurements.

This study was reviewed and approved by the Ethics Committee of School of Public Health (Shenzhen), Sun Yat-Sen University (2021. No.8), as well as the Institutional Board of Shenzhen Baoan Women’s and Children’s Hospital (LLSC2021-04-01-10-KS) and Longgang District Maternity and Child Healthcare Hospital of Shenzhen City (LGFYYXLLL-2022-024). The study strictly adhered to regulations governing ethical considerations and personal data protection. Data collection was approved by the Human Genetic Resources Administration of China (HGRAC). Written informed consent of all participants were obtained.

### Biochemical measurements and definition of thyroid-related traits

Because TSH and FT4 concentrations during pregnancy are related to gestational week, pregnancy-specific reference ranges are recommended[1]. In this project, the diagnosis of thyroid dysfunction was based on a method-specific and pregnancy-specific reference interval, as recommended by the Chinese Medical Association Guideline on the diagnosis and management of thyroid diseases during pregnancy and postpartum (2nd edition)[60]. We also used the cutoff value provided by the kit company to determine whether the subject was in a TPOAb positivity state.

In the Longgang study, thyroid function tests measured concentrations of thyroid stimulating hormone (TSH), free thyroxine (FT4), and anti-thyroid peroxidase antibody (TPOAb). These measurements were taken during a physical examination using the UniCel DxI 800 Immunoassay System (Beckman Coulter Inc., Brea, CA, USA) along with its respective system accessory kit. The reference ranges for TSH are 0.05-3.55 mIU/L for the first trimester, 0.21-3.31 mIU/L for the second trimester, and 0.43-3.71 mIU/L for the third trimester. The reference ranges for FT4 are 9.01-15.89 pmol/L for the first trimester, 6.62-13.51 pmol/L for the second trimester, and 6.42-10.75 pmol/L for the third trimester[60]. TPOAb positivity was defined according to the cutoff (TPOAb > 9 IU/mL).

In Baoan study, serum levels of TSH, FT4, and TPOAb were measured by Abott I2000 Immunoassay System. The reference ranges of TSH are (0.07-3.38 mIU/L), (0.34-3.51 mIU/L) and (0.34-4.32 mIU/L) for the first trimester, second trimester and third trimester, respectively. And the reference ranges of FT4 are (11.30-17.80 pmol/L), (9.30-15.20 pmol/L) and (7.90-14.10 pmol/L) for the first trimester, second trimester, and third trimester, respectively[60]. The cut-off value we used for TPOAb positivity is 5.6 IU/ml.

In each study, a pregnant woman with both TSH and FT4 levels within the pregnancy-specific reference range was considered a subject with normal thyroid function (ie, euthyroidism). The serum TSH and FT4 levels were analyzed as continuous variables after inverse normal transformation, based on the euthyroid subjects. Additionally, for the GWAS of continuous TPOAb levels, pregnant women exhibiting TPOAb levels below the minimum detection limits of the assay (0.25 IU/ml for the Longgang Study and 0.159 IU/ml for the Baoan Study) were excluded. TPOAb levels were natural log-transformed before performing the GWAS analysis.

The thyroid dysfunction was defined following the 2017 medical guidelines of the American Thyroid Association for the Diagnosis and Management of Thyroid Disease During Pregnancy and the Postpartum[1]. Specifically, overt hyperthyroidism was defined as a TSH concentration below the lower limit of the reference range, accompanied by an FT4 concentration exceeding the upper limit of the reference range. Subclinical hyperthyroidism was defined as a TSH concentration below the lower limit of the reference range while maintaining a normal FT4 concentration. Subclinical hypothyroidism was defined by as a TSH concentration upper the limit of the reference range with normal FT4 concentration. Isolated hypothyroxinemia is typically defined as having an FT4 concentration below the lower limit of the reference range while maintaining a normal maternal TSH concentration.

To comprehend the genetic underpinnings of these thyroid dysfunctions, we conducted a GWAS analysis on four categorical traits: subclinical hypothyroidism, isolated hypothyroxinemia, overt hyperthyroidism, and subclinical hyperthyroidism (case group). We used pregnant women with euthyroid function, defined as those maintaining TSH and FT4 levels within the pregnancy-specific reference range throughout pregnancy, as the control group. Specifically, we excluded individuals who tested positive for TPOAb from both the case and control populations in the analysis for isolated hypothyroxinemia **(Table S1)**. It is worth noting that we also examined overt hypothyroidism; however, the statistical power was limited due to the small number of cases (N_Longgang_=464, N_Baoan_=148) and we did not identify robust genetic associations.

### Statistical analysis

For all participants, baseline characteristics were described using mean ± standard deviation, and results of thyroid function tests (TSH, FT4, and TPOAb) for different pregnancies were presented in median [interquartile range]. In the phenotype association analysis of thyroid-related traits with gestational complications and fetal obstetric outcomes, quantitative traits were analyzed using multiple linear regression, and categorical traits were analyzed using logistic regression. All statistical analyses were conducted using R version 4.3.2.

### Genotyping, Imputation, and Variant annotation

Non-invasive prenatal testing (NIPT), which employs low-pass massively parallel sequencing of cell-free DNA fragments from peripheral blood of a pregnant woman, has gained prominence as an unparalleled screening test for fetal aneuploidy due to its high sensitivity and specificity[61]. Our prior research and recent study have illustrated that when NIPT data (0.06x – 0.3x) are integrated with a reference panel, it can achieve high accuracy in genotype imputation, matching or even exceeding the performance of an array[17,18].

In summary, we first collected data from NIPT for 121,578 pregnant women in Baoan and Longgang Hospitals in Shenzhen city, Guangdong Province. Each of these participants was whole-genome sequenced to 9.9-21.9 million cleaned reads, representing a sequencing depth of around 0.11x-0.25x. Next, we applied BWA[62] to align the cleaned reads to the Genome Reference Consortium Human Reference 38 (GRCh38) and used the rmdup option in samtools[63] to remove potential PCR duplicates. The GATK realignment and base quality recalibration method was utilized to align the reads and adjust base quality scores[64]. After that, the alignment files were stored as bam files.

Subsequently, we employed the Glimpse software (version 1.1.1)[65], which implements an algorithm for genotype imputation, in conjunction with a deep whole-genome sequencing reference panel of 10K Chinese[66], to perform individual genotype imputation for the NIPT data. Finally, variant annotation was carried out using Ensembl Variant Effect Predictor[67] (version 101), utilizing indexed GRCh38 cache files (version 109). All the data used for annotation were obtained in advance from the Ensembl FTP server (https://ftp.ensembl.org/pub/). Considering that a variant can span across multiple transcripts, we employed the "--pick" option to assign a single consequence block to each variant based on a predefined set of default criteria in VEP. In addition, the "--nearest" option was applied to identify the nearest gene with a protein-coding transcription start site (TSS) for variants located in the intergenic region.

### Genome-wide association analysis in individual Studies

The association of the SNPs was analyzed by GWAS in PLINK 2.0, using linear regression for three continuous traits (levels of TSH, FT4, and TPOAb) and logistic regression for five binary phenotypes (subclinical hypothyroidism, isolated hypothyroxinemia, subclinical hyperthyroidism, overt hyperthyroidism, and TPOAb positivity), separately in two cohorts (Baoan and Longgang). The genotype-phenotype association was conducted using an additive genetic model on SNP dosages. Moreover, analyses were adjusted for the gestational week of the thyroid test, maternal age, and the top ten principal components to account for population stratification.

### Meta-analysis

We conducted fixed-effect GWAS meta-analyses based on an inverse-variance weighting of Baoan and Longgang using METAL software[68]. To visualize the meta-analysis results, we generated a Manhattan plot and a quantile–quantile (QQ) plot with an R script.

In terms of interpreting the results of the meta-analysis, we defined the lead SNP as the SNP with the most significant P-value in the single-SNP association test within a 1-Mbp window. A locus was considered to have reached the significance threshold if the lead SNP within that locus had a P-value ≤ 5 × 10^−8^. A locus is considered novel locus for the trait if its leading SNP is in linkage equilibrium (LD R^2^<0.2) with any known variants (listed in the GWAS Catalog as of December 31, 2023) on the same chromosome. Specifically, using the GWAScatalog database, we obtained all the trait-related known variants located on the same chromosome as our novel lead variants (Triat ID in the Experimental Factor Ontology: TSH: EFO_0004748, FT4: EFO_0005130, thyroid peroxidase antibody measurement: EFO_0005666, and hyperthyroidism: EFO_0009189)[69]. And then, for all of our novel SNPs, we use East Asians (EAS) and Europeans (EUR) populations in the 1000G genome to calculate the linkage disequilibrium R-square between variants we identified and all known SNPs on the same chromosome using LDpair (https://ldlink.nih.gov/?tab=ldpair). And the results of linkage disequilibrium R-square between novel variants and known variants are shown in **Table S4A**.

### Conditional analysis

Stepwise conditional analyses were performed for the GWAS-meta results to identify independent signals that affect TSH and FT4 levels, conditioning on the most significant variants (known and novel) identified in our GWAS. After conditioning on the lead genome-wide significant variant, variants identified within a 1 MB region of the variant with a p-value < 5.0 × 10^−8^ were considered independent significant signals. These conditional analyses were repeated, adding in the conditional lead variants until no variant had a conditional p-value less than the genome-wide significance (P < 5.0 × 10^−8^). To determine whether the identified independent significant signals at known loci were independent novel signals, we analyzed East Asian (EAS) and European (EUR) populations from the 1000 Genomes Project. We used LDpair (https://ldlink.nih.gov/?tab=ldpair) to calculate the linkage disequilibrium R-square (R²) between the variants we identified and those previously associated with trait-linked traits. Subsequently, we defined an SNP as an independent significant signal at a locus if the R² was less than 0.2 in both EAS and EUR populations.

### Consistency and replication

To enhance the reproducibility of the findings, we checked internal and external consistency across different datasets. Consistency between the two hospitals involved examining the consistency of the effect direction of the lead SNPs and assessing the significance of the GWAS P-values in the Baoan and Longgang Studies. On the other hand, consistency of our meta-GWAS results and external datasets involved examining the consistency in an independent dataset from three separate cohorts (ThyroidOmics Consortium, the GWAS meta-analysis of Alexander T Williams et al., the GWAS meta-analysis of M. Medici et al)[10,11,34].

Beyond the participants previously referenced, our research included 4,688 individuals who attended Baoan District Maternal and Child Health Hospital for maternity check-ups during their 40-week gestation. Throughout 2020 and 2021, these participants underwent non-invasive prenatal testing (NIPT) in the first or second trimester. These samples notably underwent more in-depth sequencing than standard NIPT, achieving an average depth of 0.3x. The serum levels of TSH, FT4, and TPOAb in NIPT PLUS cohort in were measured by Abott I2000 Immunoassay System with its respective system accessory kit. Using a process similar to Baoan Study, we completed GWAS for thyroid-related traits in the NIPT PLUS cohort.

### eQTL Colocalization analysis

Colocalization analyses were performed for novel lead variants using *coloc* R package[70]. Gene expression data from 49 tissues in European ancestry samples included in the GTEx Project version 8 release were used (https://gtexportal.org/)[71]. GWAS data from a region spanning ±1 Mb around each novel SNPs were compiled, integrated with the corresponding GTEx eQTL data for each specific tissue, and subsequently used as the input for related analyses. Regions that show evidence of colocalization between eQTL and GWAS signals were identified using pre-defined thresholds: PP4 ≥ 0.75 and PP4/PP3 ≥ 3[70]. Here, PP4 denotes the posterior probability of Hypothesis 4, which assesses the likelihood that both the assumed model is correct and that the GWAS and expression data are simultaneously associated with a single causal variant. PP3 denotes the posterior probability that two distinct causal variants exist for GWAS and expression data, respectively. Given the genetic structural differences between our East Asian cohort and European populations, our findings should be considered an exploratory endeavor.

### Genetic correlation estimate

The meta GWAS summary statistics were used to estimate SNP-based heritability and genetic correlation (*r_g_*) for all phenotypes using LD Score Regression[36,72]. GCTA was used to calculate LD score files using a deep whole-genome sequencing reference panel of 10k Chinese[36,66,73].

### Mendelian randomization

Mendelian randomization (MR) is a method for inferring causal associations between exposures and outcomes through the use of genetic variants as instrumental variables [74]. To avoid bias caused by linkage disequilibrium, we selected the IV (instrumental variable) SNPs that reached genome-wide significance in GWAS meta-analysis (P < 5 × 10^−8^) and achieved independence in linkage disequilibrium (LD) r^2^ = 0.2 by GCTA-COJO analysis[75]. The SNPs with a minor allele frequency (MAF) of <0.01 were excluded[76].

Since GWAS summary data for maternal and fetal pregnancy outcomes in East Asian ancestry is currently unavailable, we conducted Mendelian randomization analyses between thyroid-related traits and maternal and fetal pregnancy outcomes using the two-sample method within a single cohort[45,77], utilizing the TwoSampleMR package after completing our GWAS-meta analysis of the Baoan Study and Longgang Study[78]. We implemented MR analyses under 3 scenarios:

1. Meta-Meta: the two-sample MR method is conducted in one-sample setting;
2. Baon-Longgang: Baoan Study served as the source of exposed GWAS data, while the Longgang Study provided the outcome population.
3. Longgang-Baoan: Longgang Study served as the source of exposed GWAS data, while the Baoan Study provided the outcome population.

In order to achieve sufficient power as much as possible, we regard meta-meta as the main MR result, and the MR results of the remaining two situations serve as support for the meta-meta result.

The variance of continuous traits explained by variants (R²) was estimated using the formula *R² = (effect size)² × 2 × MAF × (1-MAF) / [(effect size)² × 2 × MAF × (1-MAF) + (SE)² × 2 × N × MAF × (1-MAF)]*, where the effect size is measured in units of the continuous traits after inverse normal transformation. We calculated the F-statistic to evaluate the instrument strength, with values greater than 10 indicating sufficient strength. We applied inverse variance-weighted (IVW), inverse variance-weighted using multiplicative random effects (IVWRE), MR-Egger, weighted median (WM) methods, weighted mode, and outlier methods using MR PRESSO[79]. If MR PRESSO successfully excludes outlier IVs, we re-execute the MR analysis using the new set of IV SNPs. To avoid the effects of the presence of heterogeneity in IVs, the IVWRE method is regarded as the most accurate estimation.

### Maternal and fetal pregnancy outcomes

We made full use of relevant data on gestational diabetes and gestational hypertension in our cohort. Because the prevalence of gestational hypertension and preeclampsia was less than 2% in our cohort (gestational hypertension: 0.95% for Longgang and 0.47% for Baoan; preeclampsia: 0.42% for Longgang and 1.39% for Baoan), we did not include these two pregnancy complications in our analysis.

Finally, we included five blood glucose traits (FPG: Fasting Plasma Glucose; HBA1c: Glycated Hemoglobin; OGTT0H: Oral Glucose Tolerance Test at 0 hours; OGTT1H: Oral Glucose Tolerance Test at 1 hour; OGTT2H: Oral Glucose Tolerance Test at 2 hours) and two blood pressure traits (SBP: Systolic Blood Pressure; DBP: Diastolic Blood Pressure) in our analysis. We completed the meta-GWAS analyses for these seven quantitative traits and Gestational Diabetes Mellitus (GDM).

Furthermore, we conducted GWAS and meta-GWAS analyses for six birth outcomes, including gestational duration (birth gestational days), preterm birth (deliveries occurring at less than 37 weeks gestational age), birth weight, birth length, low birth weight (defined as birth weight less than 2500g), and macrosomia (defined as birth weight more than 4000g)[5,80]. For the birth outcomes of birth gestational days and preterm birth, we limited the study subjects to mothers who delivered live-born offspring through vaginal delivery.

Given that some studies have shown that TPOAb positivity with euthyroid is also a risk factor for various pregnancy complications[1], we included euthyroid TPOAb positivity as an exposure to our analysis. Therefore, after deciding on the list of IV SNPs, we conducted MR analyses of associations between 9 thyroid-related traits (TSH levels within reference range during pregnancy, FT4 levels within reference range during pregnancy, subclinical hypothyroidism during pregnancy, isolated hypothyroxinemia during pregnancy, overt hyperthyroidism during pregnancy, subclinical hyperthyroidism during pregnancy, TPOAb levels during pregnancy, TPOAb positivity with euthyroid during pregnancy, and TPOAb positivity during pregnancy) and 14 maternal and neonatal outcomes (GDM, FPG, HBA1c, OGTT0H, OGTT1H, OGTT2H, SBP, DBP, birth gestational days, preterm birth, birth weight, birth length, low birth weight, macrosomia).

We also used phenotypic associations to explore associations between 9 thyroid traits and 14 maternal and neonatal outcomes. We collected birth outcome data for 92,132 pregnant women with thyroid function test results from two hospitals (**Table S16**). Before performing phenotypic association analyses, we excluded pregnant women with multiple pregnancies, vitro fertilization, and aborted stillbirth outcomes. In the regression analysis of birth length, birth weight, low birth weight, and Macrosomia, we adjusted for fetal sex, BMI, age, and birth gestational days. For the analysis of gestational duration and preterm birth, we adjusted only for fetal sex, maternal BMI, and maternal age.

Because the 9 thyroid trait exposures are highly correlated (Figure S17, Table S14), we computed the Bonferroni correction mainly based on the number of outcomes. A Bonferroni correction for the 14 traits (P-value<□0.05/14□=0.0036) was applied on the IVWRE P-valueCto define significant results of phenotypic associations and MR.

### Biobank Japan Study (BBJ)

To investigate the potential causal association between thyroid traits during pregnancy and the physical conditions/diseases in older age, we conducted a bidirectional two-sample Mendelian randomization study using our GWAS results and datasets from the BioBank Japan Study. The BioBank Japan Study conducted genome-wide association studies for 220 deep phenotypes (including diseases, biomarkers, and medication usage) in a cohort of 179,000 Japanese individuals[30]. Approximately 47% of these subjects were women, and the average age of all subjects was over 62 years[81].

A Bonferroni correction for the 220 traits (P-value□<□0.05/220□=0.00023) was applied on the IVWRE P-value to define significant MR results.

## Data Availability

GWAS summary statistics for the eight thyroid traits investigated can be accessed through the GWAS catalog and GWAS Atlas upon publication, with approval from the Human Genetic Resources Administration of China (HGRAC) (ID: 2024BAT00834). Researchers can register and gain access to use data. Raw sequencing data have been deposited to the Genome Sequence Archive (GSA) for Human (https://ngdc.cncb.ac.cn/gsa-human/) at the BIG Data Center, Beijing Institute of Genomics, Chinese Academy of Sciences, under the BioProject accession number (GSA-Human: HRA006833), with approval from HGRAC (ID: XXX). In compliance with the regulations of the Ministry of Science and Technology of China, the raw sequencing data contain information unique to an individual and thus require controlled access. Researchers who are interested in collaboration are welcome to contact Siyang Liu.

https://www.ebi.ac.uk/gwas/

https://ngdc.cncb.ac.cn/gwas/

## Acknowledgments

The study was supported by the Shenzhen Basic Research Foundation (20220818100717002), Guangdong Basic and Applied Basic Research Foundation (2022B1515120080), the National Natural Science Foundation of China (31900487, 82203291) and the Shenzhen Health Elite Talent Training Project. Computation of this study was supported by BrightWing High-performance Computing Platform, School of Public Health (Shenzhen) and the National Supercomputing Center in GuangZhou.

## Author contribution

Design, management, or subject recruitment of the individual studies: S. L, F. W, L. X, J. Z, and data curation: J. Z, Y. W, Z. Y, L. H, J. Z, S. H, and genotyping of the individual studies: Y. W, Y. G, Y. L, Z. Y, and statistical methods, analysis, bioinformatics, or interpretation of the results in the individual studies: Y. W, Y. G, S. C, H. Z, Z. Y, X. G; and drafting of manuscript: Y. W, S. L, Y. G, and critical revision of manuscript: all authors.

## Reference

[1] Alexander E K, Pearce E N, Brent G A, et al. 2017 Guidelines of the American Thyroid Association for the Diagnosis and Management of Thyroid Disease During Pregnancy and the Postpartum[J]. Thyroid, 2017, 27(3): 315-389.

[2] De Leo S, Pearce E N. Autoimmune thyroid disease during pregnancy[J]. Lancet Diabetes Endocrinol, 2018, 6(7): 575−586.

[3] Dong A C, Stagnaro-Green A. Differences in Diagnostic Criteria Mask the True Prevalence of Thyroid Disease in Pregnancy: A Systematic Review and Meta-Analysis[J]. Thyroid, 2019, 29(2): 278−289.

[4] Korevaar T I M, Derakhshan A, Taylor P N, et al. Association of Thyroid Function Test Abnormalities and Thyroid Autoimmunity With Preterm Birth: A Systematic Review and Meta-analysis[J]. Jama, 2019, 322(7): 632−641.

[5] Derakhshan A, Peeters R P, Taylor P N, et al. Association of maternal thyroid function with birthweight: a systematic review and individual-participant data meta-analysis[J]. Lancet Diabetes Endocrinol, 2020, 8(6): 501−510.

[6] Lee S Y, Cabral H J, Aschengrau A, et al. Associations Between Maternal Thyroid Function in Pregnancy and Obstetric and Perinatal Outcomes[J]. J Clin Endocrinol Metab, 2020, 105(5): e2015−23.

[7] Panicker V, Wilson S G, Spector T D, et al. Heritability of serum TSH, free T4 and free T3 concentrations: a study of a large UK twin cohort[J]. Clin Endocrinol (Oxf), 2008, 68(4): 652−9.

[8] Hansen P S, Brix T H, Sørensen T I, et al. Major genetic influence on the regulation of the pituitary-thyroid axis: a study of healthy Danish twins[J]. J Clin Endocrinol Metab, 2004, 89(3): 1181−7.

[9] Nolan J, Campbell P J, Brown S J, et al. Genome-wide analysis of thyroid function in Australian adolescents highlights SERPINA7 and NCOA3[J]. Eur J Endocrinol, 2021, 185(5): 743−753.

[10] Teumer A, Chaker L, Groeneweg S, et al. Genome-wide analyses identify a role for SLC17A4 and AADAT in thyroid hormone regulation[J]. Nat Commun, 2018, 9(1): 4455.

[11] Williams A T, Chen J, Coley K, et al. Genome-wide association study of thyroid-stimulating hormone highlights new genes, pathways and associations with thyroid disease[J]. Nat Commun, 2023, 14(1): 6713.

[12] Krassas G E, Poppe K, Glinoer D. Thyroid Function and Human Reproductive Health[J]. Endocrine Reviews, 2010, 31(5): 702−755.

[13] Korevaar T I M, Medici M, Visser T J, et al. Thyroid disease in pregnancy: new insights in diagnosis and clinical management[J]. Nat Rev Endocrinol, 2017, 13(10): 610−622.

[14] Porcu E, Medici M, Pistis G, et al. A meta-analysis of thyroid-related traits reveals novel loci and gender-specific differences in the regulation of thyroid function[J]. PLoS Genet, 2013, 9(2): e1003266.

[15] Fitzgerald S P, Bean N G, Fitzgerald S P, et al. The application of new concepts of the assessment of the thyroid state to pregnant women[J]. Front Endocrinol (Lausanne), 2022, 13: 987397.

[16] Cappola A R, Casey B M. Thyroid Function Test Abnormalities During Pregnancy[J]. Jama, 2019, 322(7): 617−619.

[17] Liu S, Huang S, Chen F, et al. Genomic Analyses from Non-invasive Prenatal Testing Reveal Genetic Associations, Patterns of Viral Infections, and Chinese Population History[J]. Cell, 2018, 175(2): 347−359.e14.

[18] Liu S, Huang S, Liu Y, et al. Utilizing Non-Invasive Prenatal Test Sequencing Data Resource for Human Genetic Investigation[J]. bioRxiv, 2023: 2023.12.11.570976.

[19] Hershman J M. The role of human chorionic gonadotropin as a thyroid stimulator in normal pregnancy[J]. J Clin Endocrinol Metab, 2008, 93(9): 3305−6.

[20] Kwak S H, Park Y J, Go M J, et al. A genome-wide association study on thyroid function and anti-thyroid peroxidase antibodies in Koreans[J]. Hum Mol Genet, 2014, 23(16): 4433−42.

[21] Zhan M, Chen G, Pan C M, et al. Genome-wide association study identifies a novel susceptibility gene for serum TSH levels in Chinese populations[J]. Hum Mol Genet, 2014, 23(20): 5505−17.

[22] Huang L, Bai F, Zhang Y, et al. Preliminary study of genome-wide association identified novel susceptibility genes for thyroid-related hormones in Chinese population[J]. Genes Genomics, 2022, 44(8): 1031−1038.

[23] Zhou W, Brumpton B, Kabil O, et al. GWAS of thyroid stimulating hormone highlights pleiotropic effects and inverse association with thyroid cancer[J]. Nat Commun, 2020, 11(1): 3981.

[24] Thareja G, Belkadi A, Arnold M, et al. Differences and commonalities in the genetic architecture of protein quantitative trait loci in European and Arab populations[J]. Hum Mol Genet, 2023, 32(6): 907−916.

[25] Chaker L, Razvi S, Bensenor I M, et al. Hypothyroidism[J]. Nature Reviews Disease Primers, 2022, 8(1): 30.

[26] Stagnaro-Green A. Thyroid and pregnancy — time for universal screening? [J]. Nature Reviews Endocrinology, 2017, 13(4): 192−194.

[27] Casey B M, Thom E A, Peaceman A M, et al. Treatment of Subclinical Hypothyroidism or Hypothyroxinemia in Pregnancy[J]. N Engl J Med, 2017, 376(9): 815−825.

[28] Lee S Y, Pearce E N. Assessment and treatment of thyroid disorders in pregnancy and the postpartum period[J]. Nat Rev Endocrinol, 2022, 18(3): 158−171.

[29] Poppe K, Velkeniers B, Glinoer D. The role of thyroid autoimmunity in fertility and pregnancy[J]. Nat Clin Pract Endocrinol Metab, 2008, 4(7): 394−405.

[30] Sakaue S, Kanai M, Tanigawa Y, et al. A cross-population atlas of genetic associations for 220 human phenotypes[J]. Nat Genet, 2021, 53(10): 1415−1424.

[31] Chu X, Pan C M, Zhao S X, et al. A genome-wide association study identifies two new risk loci for Graves’ disease[J]. Nat Genet, 2011, 43(9): 897−901.

[32] Kochi Y, Yamada R, Suzuki A, et al. A functional variant in FCRL3, encoding Fc receptor-like 3, is associated with rheumatoid arthritis and several autoimmunities[J]. Nat Genet, 2005, 37(5): 478−85.

[33] Mamidi M K, Huang J, Honjo K, et al. FCRL1 immunoregulation in B cell development and malignancy[J]. Front Immunol, 2023, 14: 1251127.

[34] Medici M, Porcu E, Pistis G, et al. Identification of novel genetic Loci associated with thyroid peroxidase antibodies and clinical thyroid disease[J]. PLoS Genet, 2014, 10(2): e1004123.

[35] Antonelli A, Ferrari S M, Corrado A, et al. Autoimmune thyroid disorders[J]. Autoimmun Rev, 2015, 14(2): 174−80.

[36] Bulik-Sullivan B K, Loh P R, Finucane H K, et al. LD Score regression distinguishes confounding from polygenicity in genome-wide association studies[J]. Nat Genet, 2015, 47(3): 291−5.

[37] Ortiga-Carvalho T M, Chiamolera M I, Pazos-Moura C C, et al. Hypothalamus-Pituitary-Thyroid Axis[J]. Compr Physiol, 2016, 6(3): 1387−428.

[38] Wang D, Wan S, Liu P, et al. Relationship between excess iodine, thyroid function, blood pressure, and blood glucose level in adults, pregnant women, and lactating women: A cross-sectional study[J]. Ecotoxicol Environ Saf, 2021, 208: 111706.

[39] Han Y, Wang J, Wang X, et al. Relationship Between Subclinical Hypothyroidism in Pregnancy and Hypertensive Disorder of Pregnancy: A Systematic Review and Meta-Analysis[J]. Front Endocrinol (Lausanne), 2022, 13: 823710.

[40] Thangaratinam S, Tan A, Knox E, et al. Association between thyroid autoantibodies and miscarriage and preterm birth: meta-analysis of evidence[J]. Bmj, 2011, 342: d2616.

[41] Brent G A. The debate over thyroid-function screening in pregnancy[J]. N Engl J Med, 2012, 366(6): 562−3.

[42] Stagnaro-Green A. Clinical guidelines: Thyroid and pregnancy-time for universal screening?[J]. Nat Rev Endocrinol, 2017, 13(4): 192−194.

[43] Medici M, Peeters R P, Teumer A, et al. The importance of high-quality mendelian randomisation studies for clinical thyroidology[J]. Lancet Diabetes Endocrinol, 2019, 7(9): 665−667.

[44] Zhang X, Wu P, Chen Y, et al. Does Maternal Normal Range Thyroid Function Play a Role in Offspring Birth Weight? Evidence From a Mendelian Randomization Analysis[J]. Front Endocrinol (Lausanne), 2020, 11: 601956.

[45] Minelli C, Del Greco M F, Van Der Plaat D A, et al. The use of two-sample methods for Mendelian randomization analyses on single large datasets[J]. Int J Epidemiol, 2021, 50(5): 1651−1659.

46. Fernández Alba J J, Castillo Lara M, Jiménez Heras J M, et al. High First Trimester Levels of TSH as an Independent Risk Factor for Gestational Diabetes Mellitus: A Retrospective Cohort Study[J]. J Clin Med, 2022, 11(13).

[47] Knight B A, Shields B M, Hattersley A T, et al. Maternal hypothyroxinaemia in pregnancy is associated with obesity and adverse maternal metabolic parameters[J]. Eur J Endocrinol, 2016, 174(1): 51−7.

[48] Haddow J E, Craig W Y, Neveux L M, et al. Free Thyroxine During Early Pregnancy and Risk for Gestational Diabetes[J]. PLoS One, 2016, 11(2): e0149065.

[49] Giontella A, Lotta L A, Overton J D, et al. Association of Thyroid Function with Blood Pressure and Cardiovascular Disease: A Mendelian Randomization[J]. J Pers Med, 2021, 11(12).

[50] Zhou Y, Liu Y, Zhang Y, et al. Identifying Non-Linear Association Between Maternal Free Thyroxine and Risk of Preterm Delivery by a Machine Learning Model[J]. Front Endocrinol (Lausanne), 2022, 13: 817595.

[51] Soh S B, Aw T C. Laboratory Testing in Thyroid Conditions-Pitfalls and Clinical Utility[J]. Ann Lab Med, 2019, 39(1): 3−14.

[52] Daramjav N, Takagi J, Iwayama H, et al. Autoimmune Thyroiditis Shifting from Hashimoto’s Thyroiditis to Graves’ Disease[J]. Medicina (Kaunas), 2023, 59(4).

[53] Evangelopoulou M E, Alevizaki M, Toumanidis S, et al. Mitral valve prolapse in autoimmune thyroid disease: an index of systemic autoimmunity?[J]. Thyroid, 1999, 9(10): 973−7.

[54] Ellervik C, Roselli C, Christophersen I E, et al. Assessment of the Relationship Between Genetic Determinants of Thyroid Function and Atrial Fibrillation: A Mendelian Randomization Study[J]. JAMA Cardiol, 2019, 4(2): 144−152.

[55] Norton M E, Jacobsson B, Swamy G K, et al. Cell-free DNA analysis for noninvasive examination of trisomy[J]. N Engl J Med, 2015, 372(17): 1589−97.

[56] Huang S, Liu S, Huang M, et al. The Born in Guangzhou Cohort Study enables generational genetic discoveries[J]. Nature, 2024, 626(7999): 565-573.

[57] Richardson T G, Sanderson E, Elsworth B, et al. Use of genetic variation to separate the effects of early and later life adiposity on disease risk: mendelian randomisation study[J]. Bmj, 2020, 369: m1203.

[58] Andersen S, Pedersen K M, Bruun N H, et al. Narrow individual variations in serum T(4) and T(3) in normal subjects: a clue to the understanding of subclinical thyroid disease[J]. J Clin Endocrinol Metab, 2002, 87(3): 1068−72.

[59] Kuś A, Chaker L, Teumer A, et al. The Genetic Basis of Thyroid Function: Novel Findings and New Approaches[J]. J Clin Endocrinol Metab, 2020, 105(6).

60. Writing Committee for Guidelines on Diagnosis and Management Ofthyroid Diseases During Pregnancy and Postpartum;Chinese Society Ofendocrinology C M A. Guideline on diagnosis and management of thyroid diseases during pregnancy and postpartum(2 edition)[J]. Chin J Endocrinol Metab, 2019, 35(8).

[61] Hartwig T S, Ambye L, Sørensen S, et al. Discordant non-invasive prenatal testing (NIPT)-a systematic review[J]. Prenat Diagn, 2017, 37(6): 527−539.

[62] Li H, Durbin R. Fast and accurate short read alignment with Burrows-Wheeler transform[J]. Bioinformatics, 2009, 25(14): 1754−60.

[63] Li H, Handsaker B, Wysoker A, et al. The Sequence Alignment/Map format and SAMtools[J]. Bioinformatics, 2009, 25(16): 2078−9.

[64] Depristo M A, Banks E, Poplin R, et al. A framework for variation discovery and genotyping using next-generation DNA sequencing data[J]. Nat Genet, 2011, 43(5): 491−8.

[65] Rubinacci S, Ribeiro D M, Hofmeister R J, et al. Efficient phasing and imputation of low-coverage sequencing data using large reference panels[J]. Nat Genet, 2021, 53(1): 120−126.

[66] Cheng S, Xu Z, Bian S, et al. The STROMICS genome study: deep whole-genome sequencing and analysis of 10K Chinese patients with ischemic stroke reveal complex genetic and phenotypic interplay[J]. Cell Discov, 2023, 9(1): 75.

[67] Cunningham F, Allen J E, Allen J, et al. Ensembl 2022 [J]. Nucleic Acids Res, 2022, 50(D1): D988-d995.

[68] Willer C J, Li Y, Abecasis G R. METAL: fast and efficient meta-analysis of genomewide association scans[J]. Bioinformatics, 2010, 26(17): 2190−1.

[69] Sollis E, Mosaku A, Abid A, et al. The NHGRI-EBI GWAS Catalog: knowledgebase and deposition resource[J]. Nucleic Acids Res, 2023, 51(D1): D977−d985.

[70] Giambartolomei C, Vukcevic D, Schadt E E, et al. Bayesian test for colocalisation between pairs of genetic association studies using summary statistics[J]. PLoS Genet, 2014, 10(5): e1004383.

71. The Genotype-Tissue Expression (GTEx) project[J]. Nat Genet, 2013, 45(6): 580-5.

[72] Bulik-Sullivan B, Finucane H K, Anttila V, et al. An atlas of genetic correlations across human diseases and traits[J]. Nat Genet, 2015, 47(11): 1236−41.

[73] Yang J, Lee S H, Goddard M E, et al. GCTA: a tool for genome-wide complex trait analysis[J]. Am J Hum Genet, 2011, 88(1): 76−82.

[74] Geng T, Smith C E, Li C, et al. Childhood BMI and Adult Type 2 Diabetes, Coronary Artery Diseases, Chronic Kidney Disease, and Cardiometabolic Traits: A Mendelian Randomization Analysis[J]. Diabetes Care, 2018, 41(5): 1089−1096.

[75] Yang J, Ferreira T, Morris A P, et al. Conditional and joint multiple-SNP analysis of GWAS summary statistics identifies additional variants influencing complex traits[J]. Nat Genet, 2012, 44(4): 369−75, s1-3.

[76] Cui Z, Hou G, Meng X, et al. Bidirectional Causal Associations Between Inflammatory Bowel Disease and Ankylosing Spondylitis: A Two-Sample Mendelian Randomization Analysis[J]. Front Genet, 2020, 11: 587876.

[77] Hemani G, Zheng J, Elsworth B, et al. The MR-Base platform supports systematic causal inference across the human phenome[J]. Elife, 2018, 7.

[78] Hemani G, Tilling K, Davey Smith G. Orienting the causal relationship between imprecisely measured traits using GWAS summary data[J]. PLoS Genet, 2017, 13(11): e1007081.

[79] Verbanck M, Chen C Y, Neale B, et al. Detection of widespread horizontal pleiotropy in causal relationships inferred from Mendelian randomization between complex traits and diseases[J]. Nat Genet, 2018, 50(5): 693−698.

[80] Goldenberg R L, Culhane J F, Iams J D, et al. Epidemiology and causes of preterm birth[J]. Lancet, 2008, 371(9606): 75-84.

[81] Nagai A, Hirata M, Kamatani Y, et al. Overview of the BioBank Japan Project: Study design and profile[J]. J Epidemiol, 2017, 27(3s): S2-s8.

