## Supplemental Figure for "Genome-wide association studies of thyroid-related hormones, dysfunction, and autoimmunity among 85,421 Chinese pregnancies"

\# Those authors contribute equally

*: Correspondence should be addressed to

Siyang Liu

Likuan Xiong

Fengxiang Wei

### Supplementary Figures

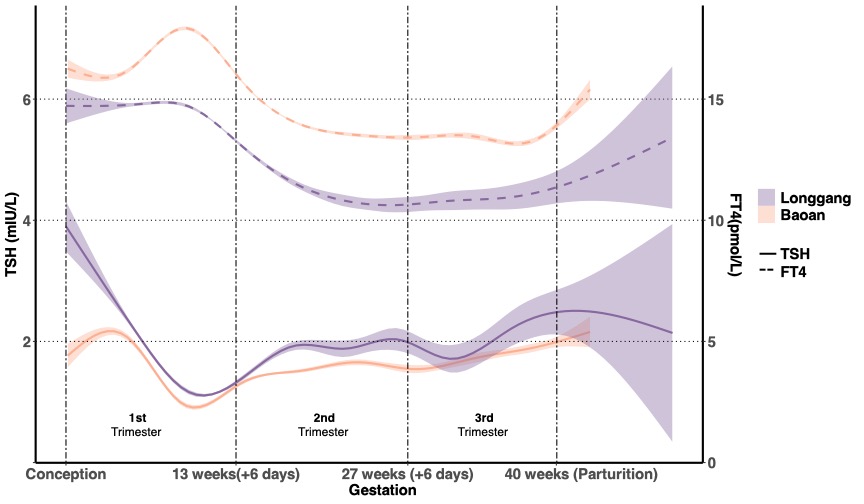

#### Figure S1. Changes in thyroid function during pregnancy.

The left and right y-axes indicate the ranges of variation in TSH and FT4 concentrations, respectively. The sample sizes for Longgang and Baoan were 58,351 and 33,781, respectively.

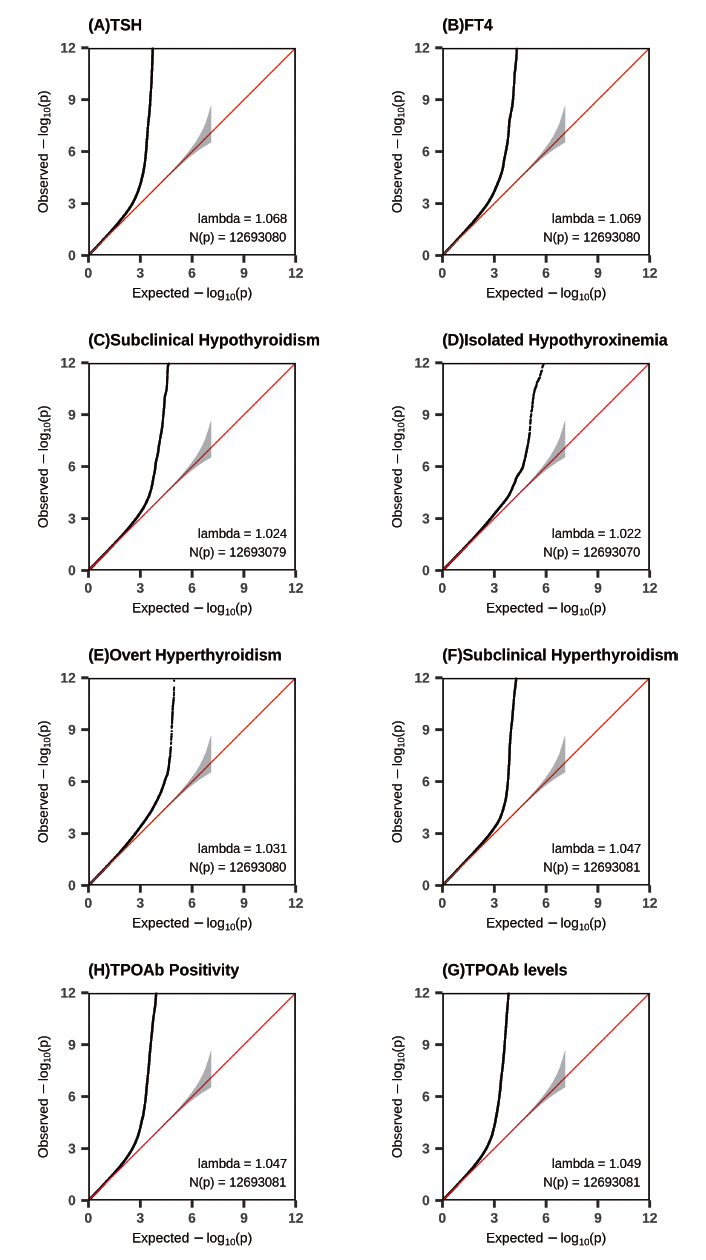

#### Figure S2. QQ plots for GWAS meta-analyses of 8 thyroid-related traits during pregnancy.

The plots show the observed -log10(P-value) in our GWAS meta-analyses against expected -log10(P-value). The red line indicates the distribution of P-values under the null hypothesis, and the gray shaded area indicates standard errors.

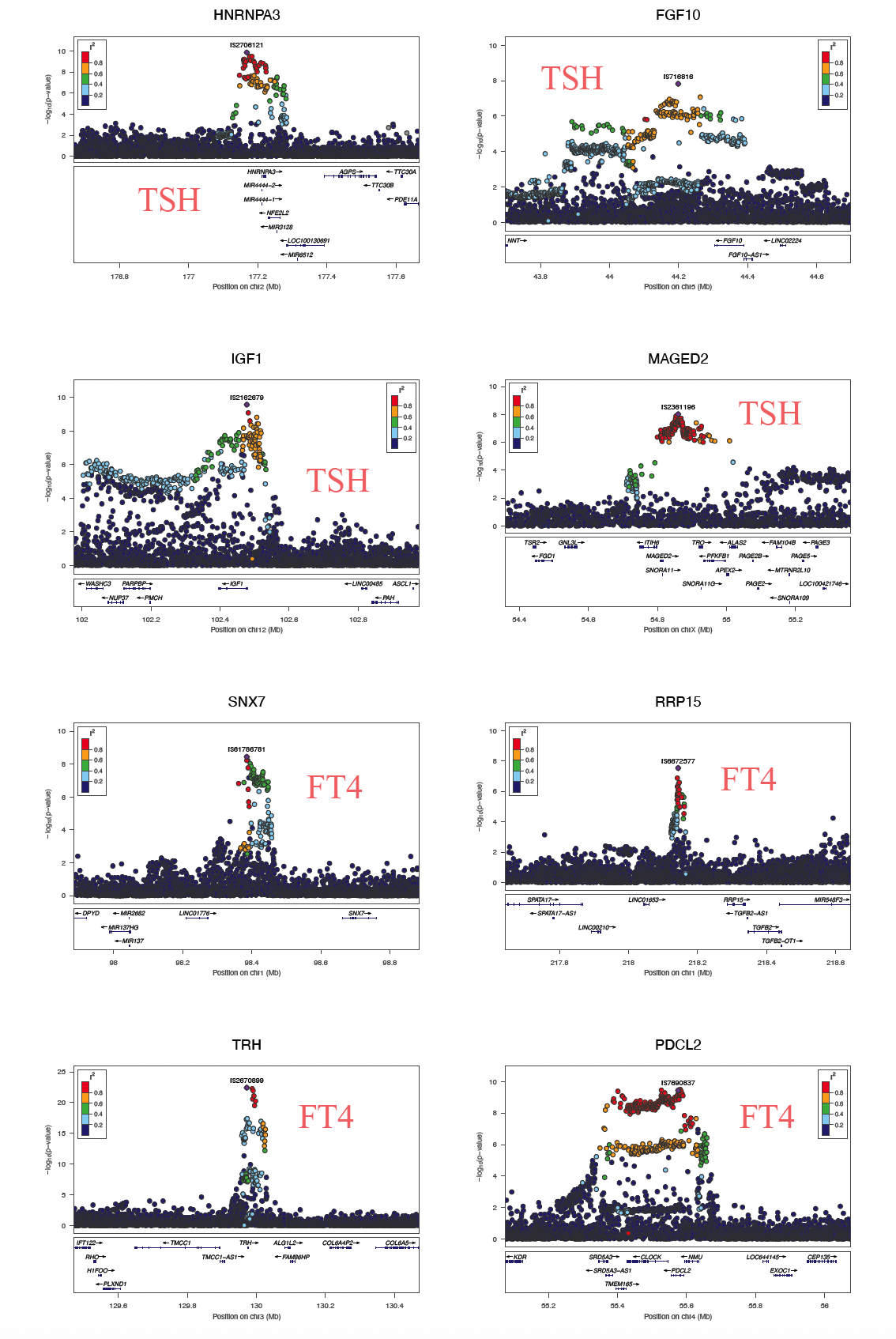

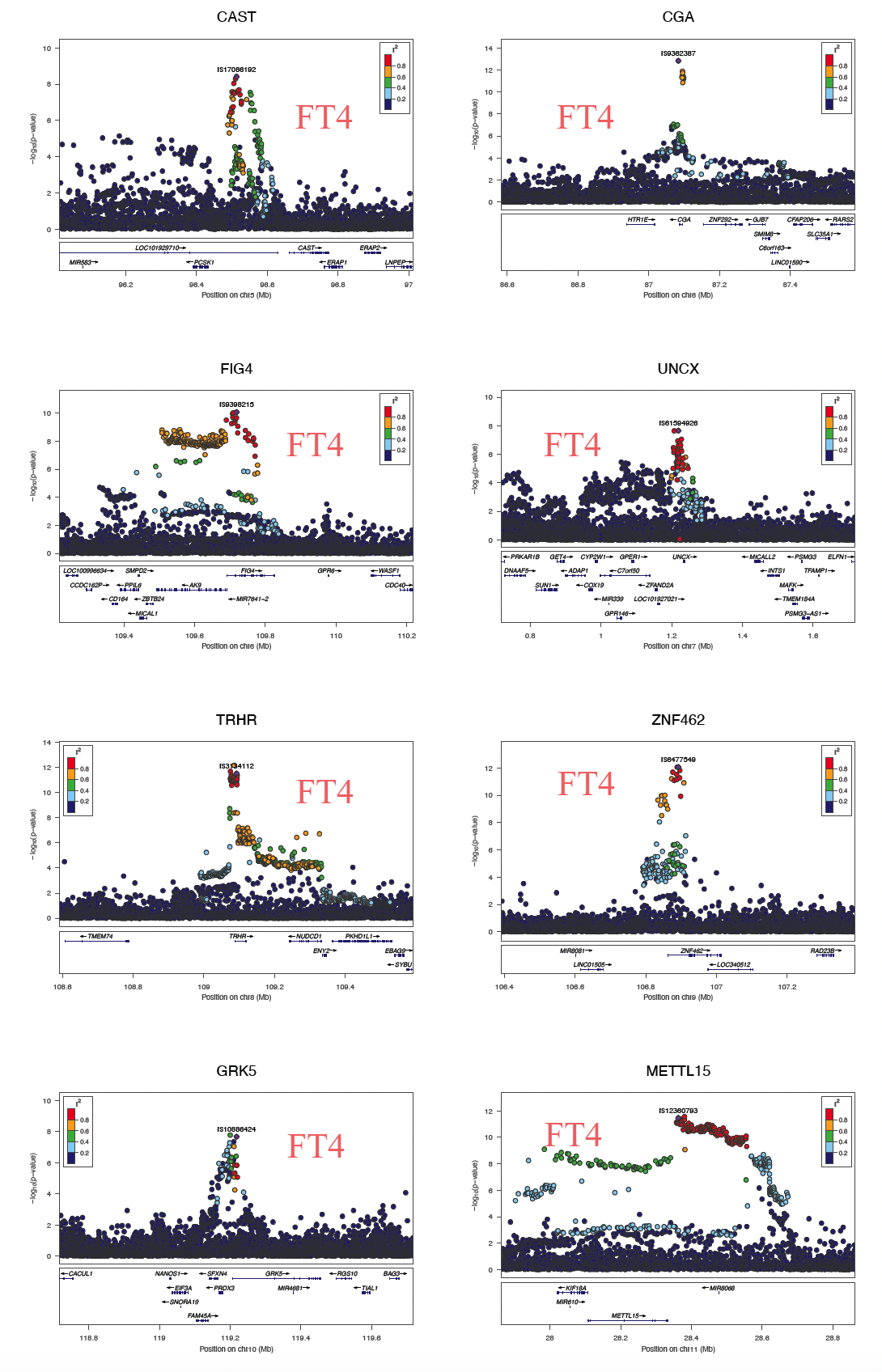

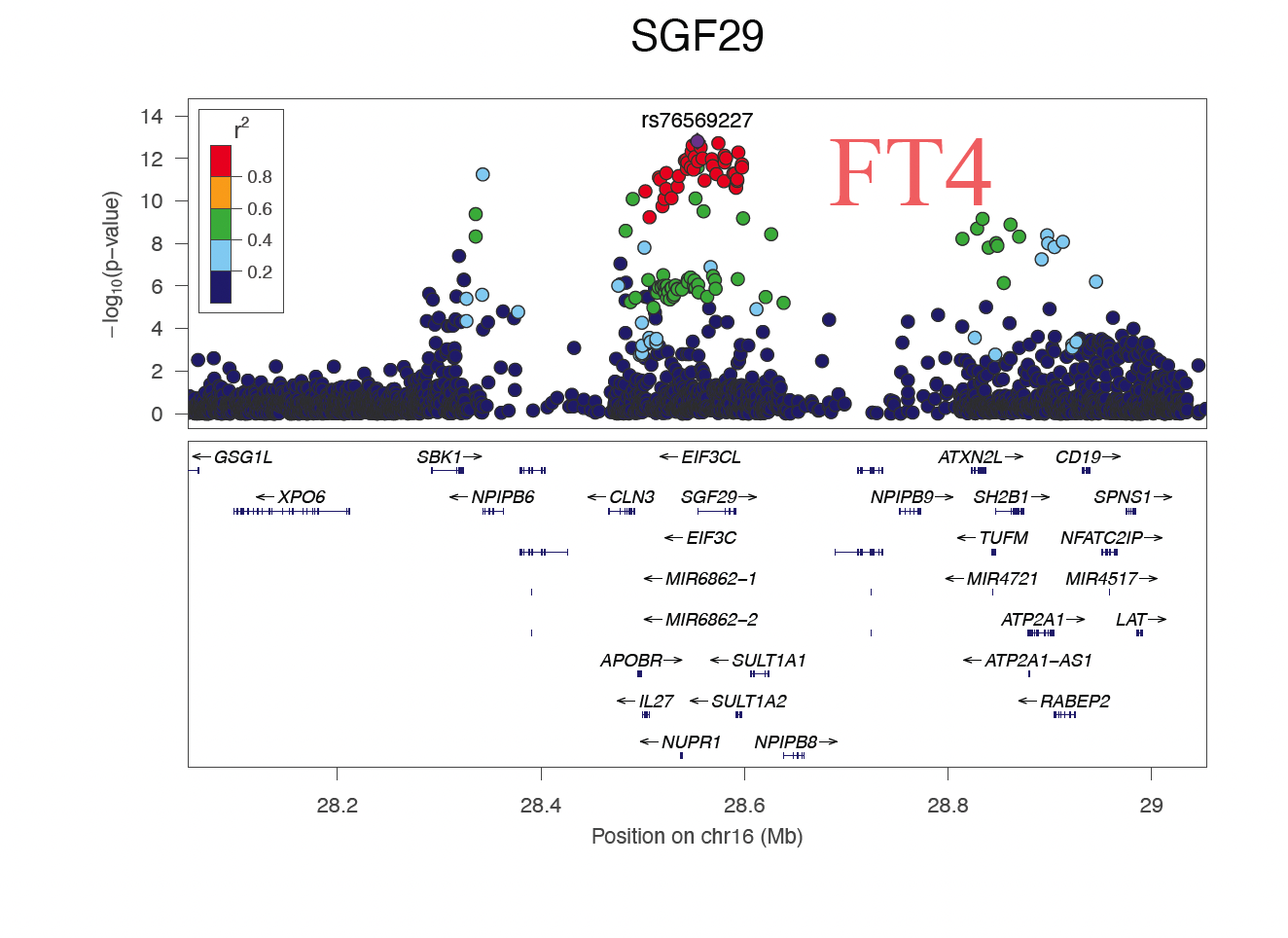

#### Figure S3. Locuszoom plots of genome-wide significant and novel loci associated with TSH/FT4 levels in the study.

For all the 17 novel lead SNPs listed in **Table S2**, the regional plots display the P-value and the LD R^2^ of SNPs located in the 500 kbp flanking regions upstream and downstream. These plots are generated using the LocusZoom software.

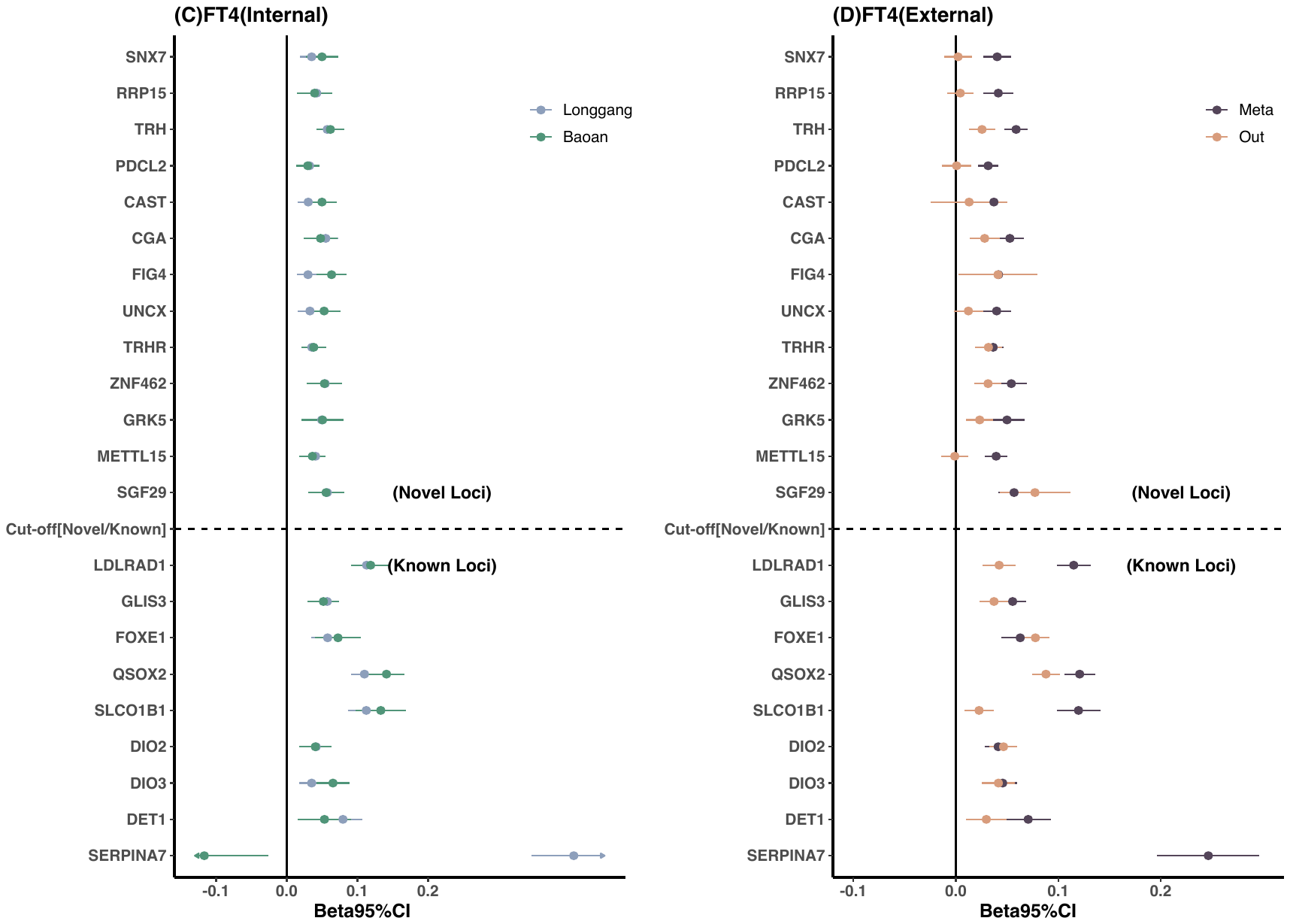

#### Figure S4. Graphical representation for comparison of effect size estimates of TSH /FT4 associated loci between two hospitals and with European GWAS results.

Shown are effect sizes and 95% confidence intervals for (A) 38 TSH-related loci in the GWAS of Longgang and Baoan Studies, (B) 38 TSH-related loci in Meta-GWAS of this study and an external study, (C) 22 FT4-related loci in the GWAS of Longgang and Baoan Studies, and (D) 22 FT4-related loci in Meta-GWAS of this study and an external study. To facilitate the presentation, when the effect size is smaller than 0 in the Longgang or meta study, we present the results after flipping the alleles of the lead SNP. The external datasets were obtained from the largest previously published GWAS study of these two phenotypes.

Beta: effect size of the GWAS; 95% CI: 95% confidence interval.

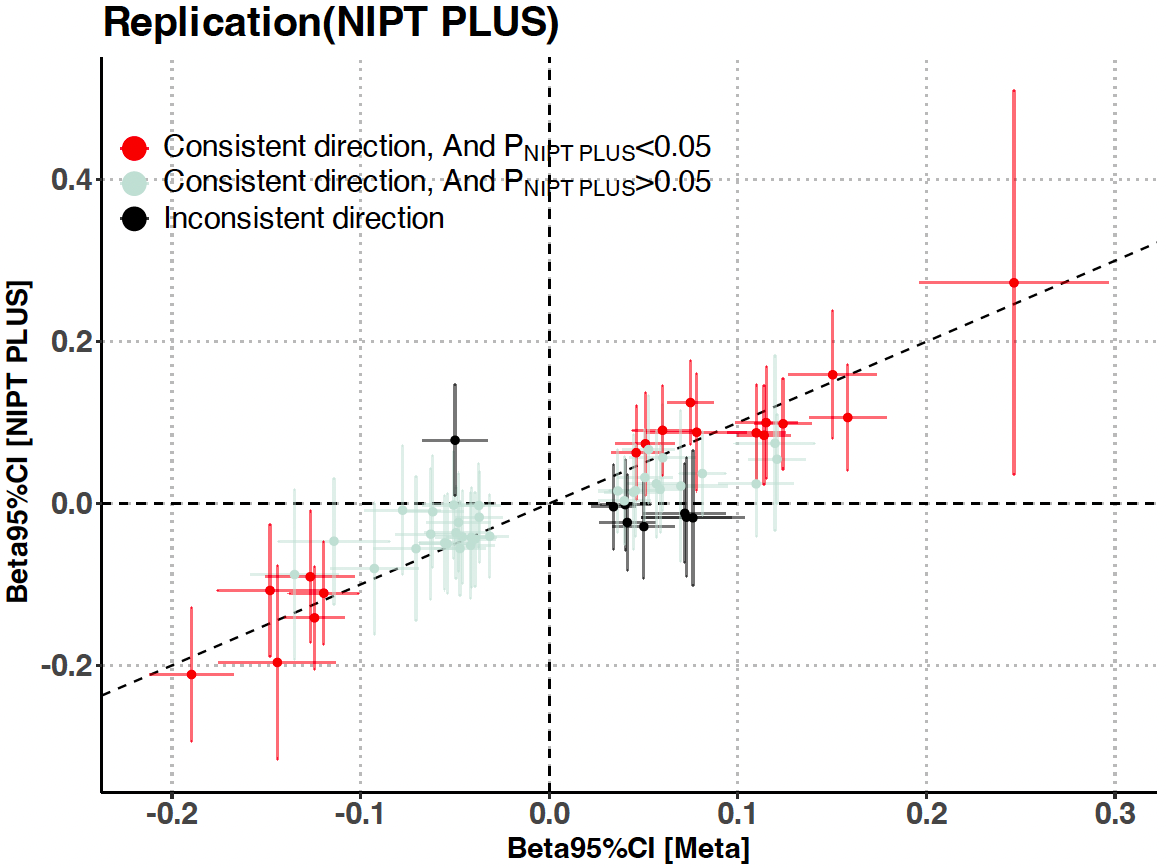

#### Figure S5. Graphical representation for replication of effect size estimates of TSH /FT4 associated loci between GWAS meta and NIPT PLUS.

Beta: effect size of the GWAS; 95% CI: 95% confidence interval.

#### Figure S6. Comparison of the frequency of the effect allele between Chinese pregnant women and European populations for 17 novel loci of TSH/FT4 levels.

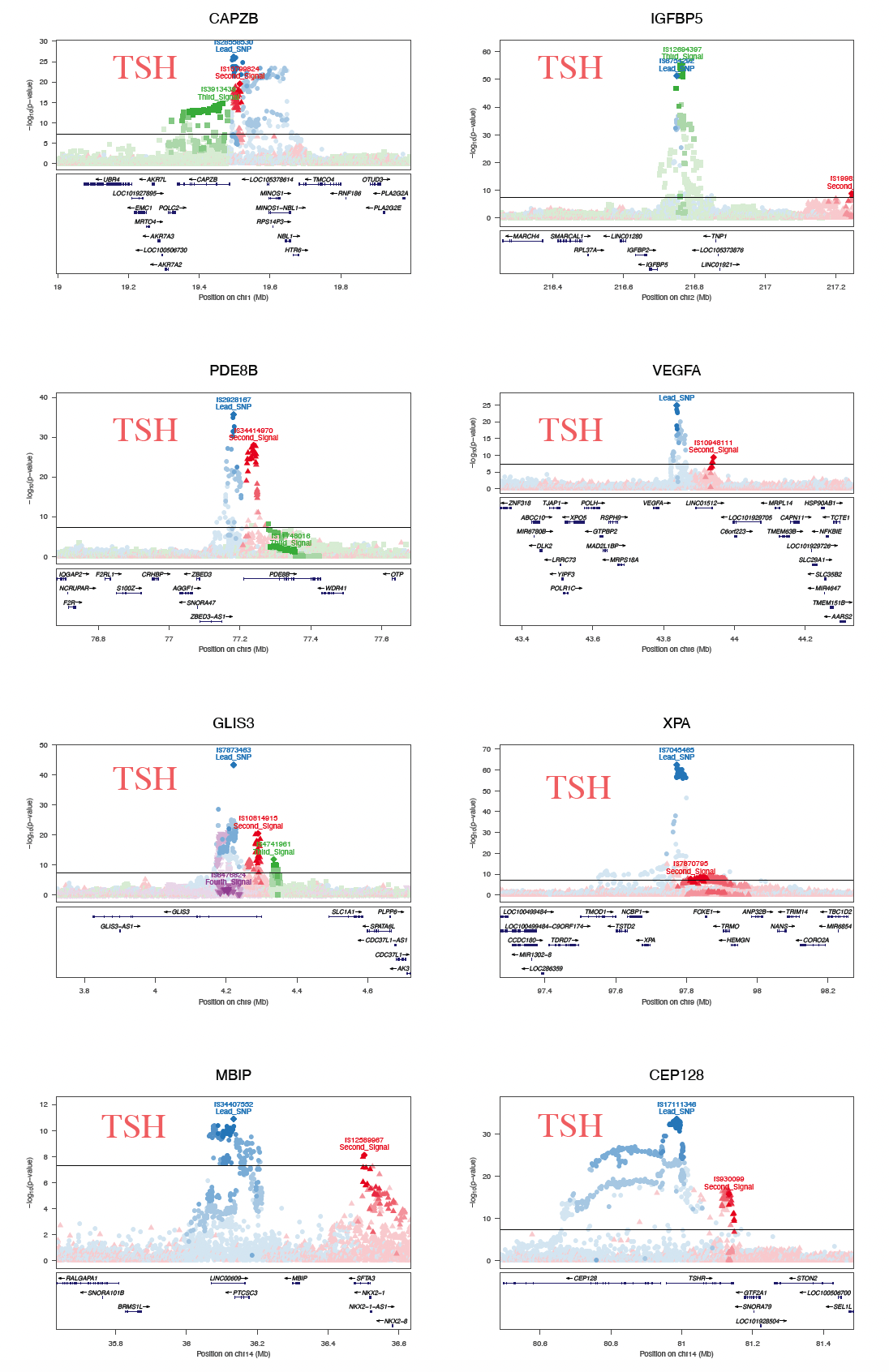

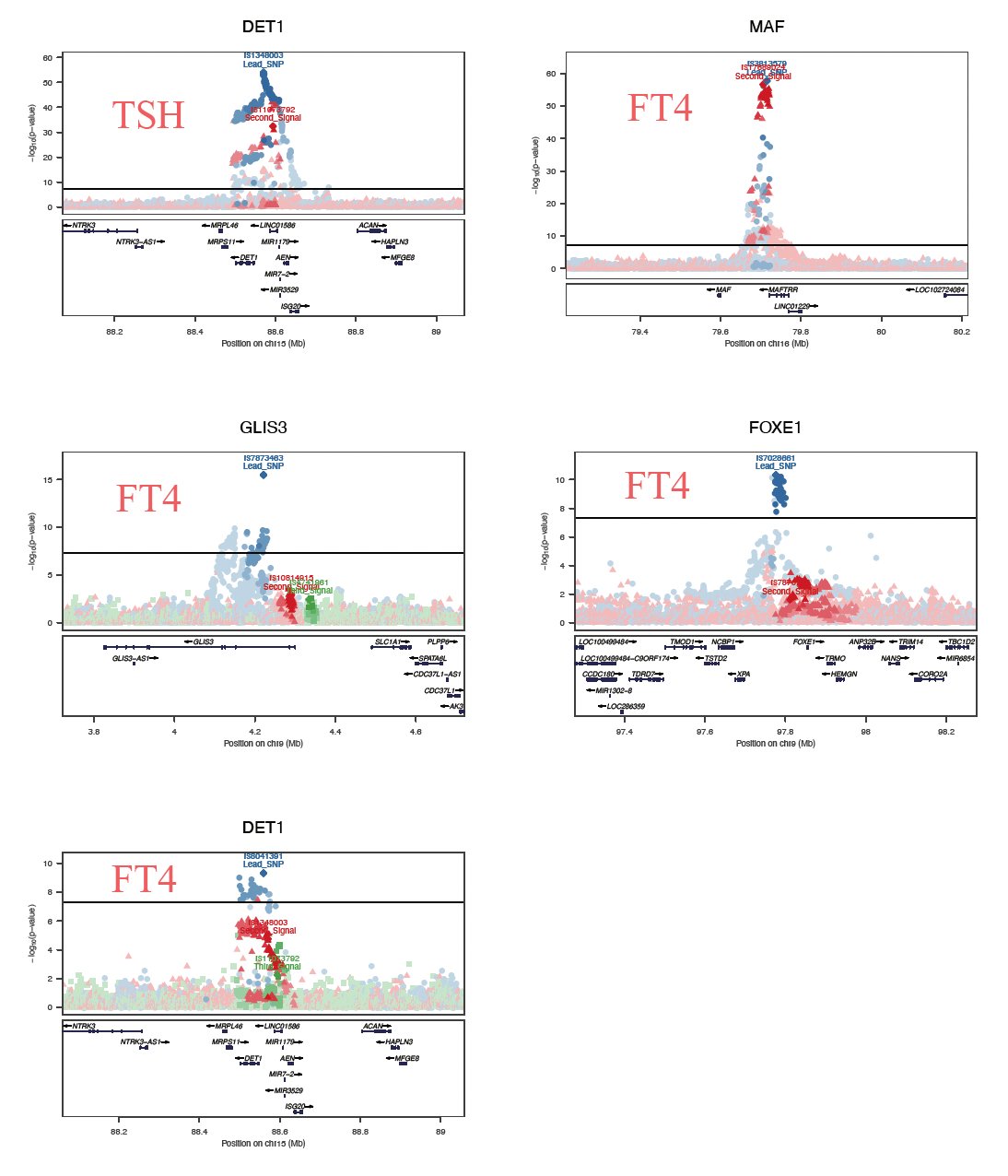

#### Figure S7. Locuszoom plots of the loci including more than a significant signal in stepwise conditional analyses.

For all 13 loci listed in **Table S7**, regional plots display the P-value and the LD R^2^ of multiple signals located in the 500 kbp flanking regions upstream and downstream. These plots are generated using LocusZoom software. Different colors indicate the number of conditional times of signals, and shades of colors show the LD R^2^ of conditional signals and SNPs located in the upstream and downstream 500 kbp flanking regions.

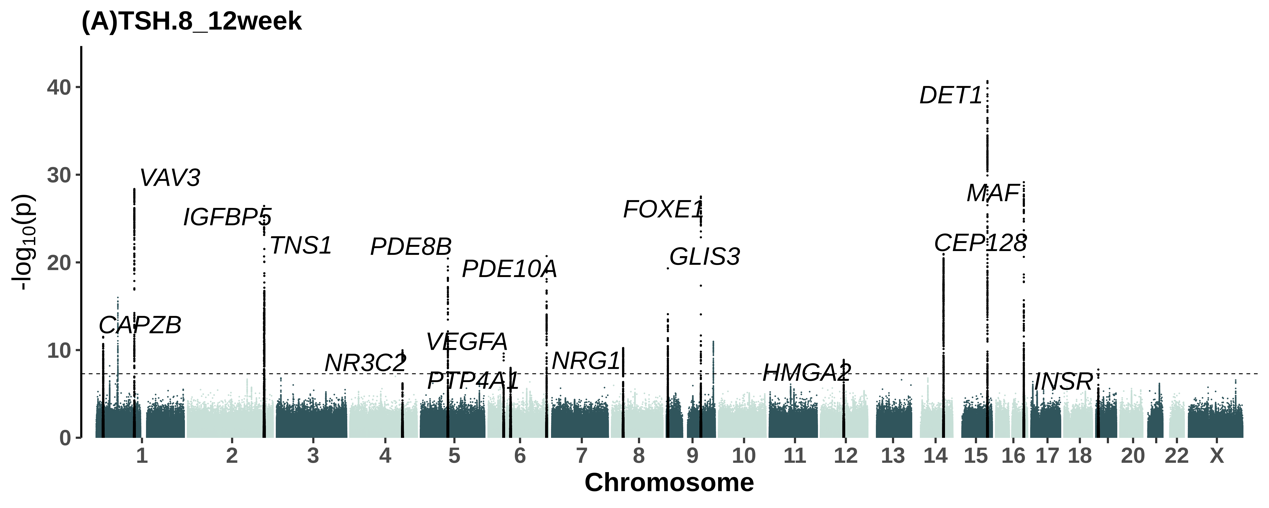

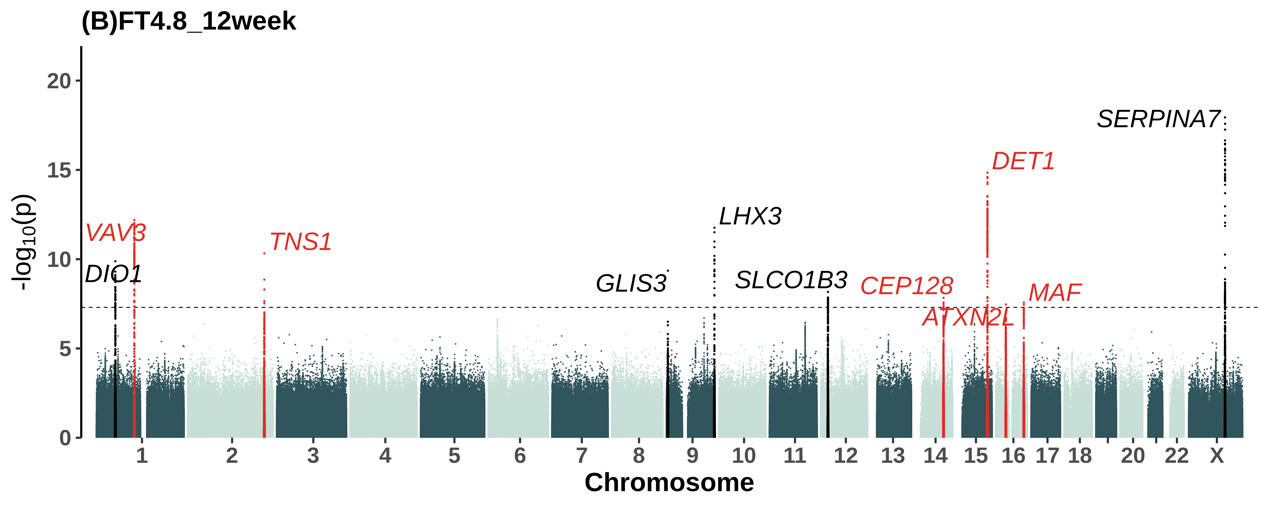

#### Figure S8. The Manhattan plots for GWAS meta-analyses of TSH (A) levels, and FT4 (B) levels within 8 to 12 weeks.

The x-coordinates of the SNPs in the plot represent their positions on each chromosome, while the y-coordinates indicate their P-values (-log10 scale) in the association test. The black dotted horizontal line represents the significance threshold for the genome-wide association test (i.e., 5E-8). Genomic loci containing phenotype-associated variants previously reported in the GWAS catalog are colored in black, and novel loci are colored in red.

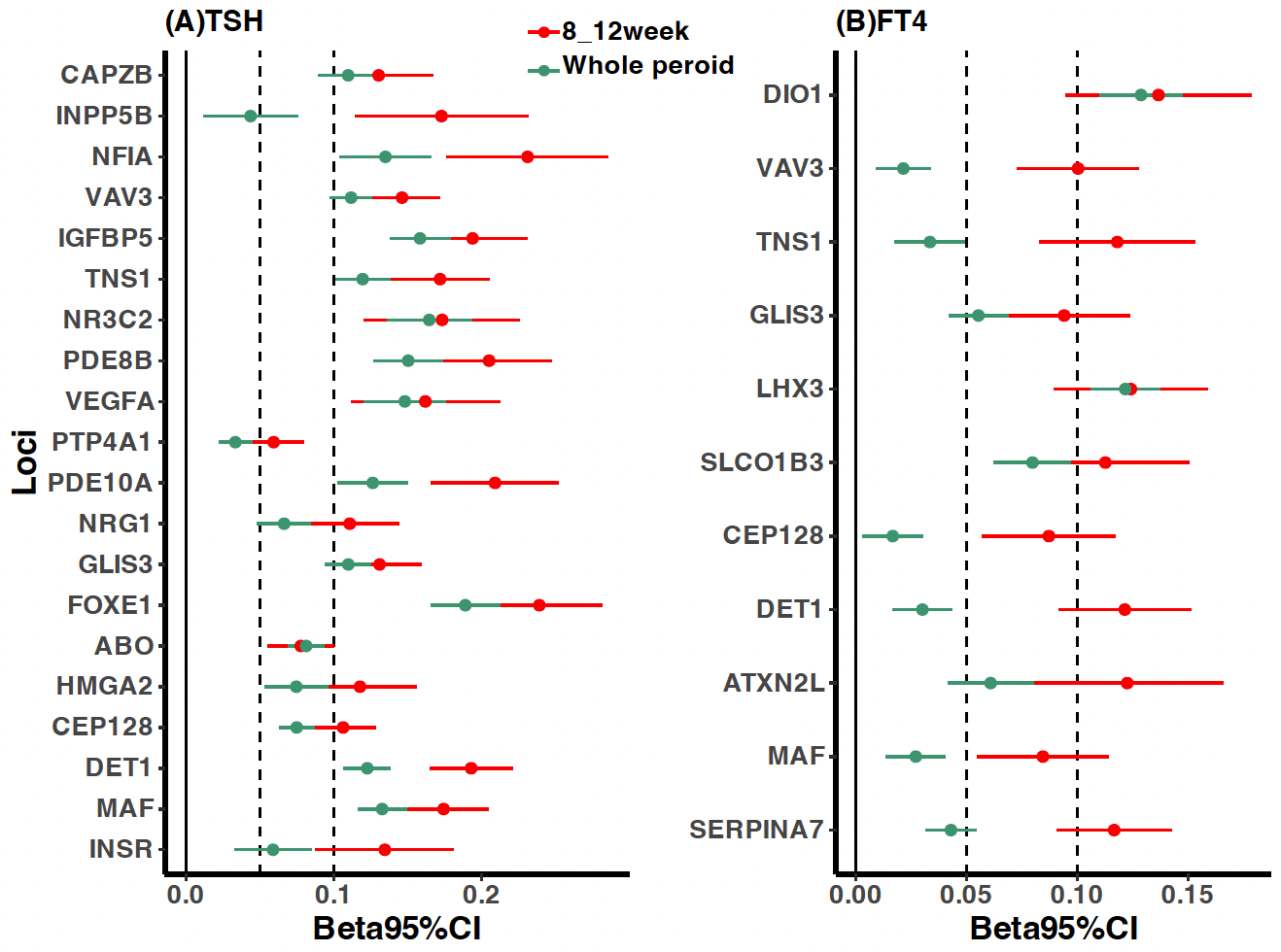

#### Figure S9. Graphical representation for comparison of effect size estimates of TSH /FT4 associated loci within 8 to 12weeks and the whole period.

Shown are effect sizes and 95% confidence intervals for (A)17 TSH-related loci in the GWAS for TSH levels over 8 to 12 weeks and over the whole period, and (B) 11 FT4-related loci in the GWAS for TSH levels over 8 to 12 weeks and over the whole period. To facilitate the presentation of the results, when the effect size is smaller than 0 in the 8 to 12 weeks group, we present the results after flipping the alleles of the lead SNP.

Beta: effect size of the GWAS; 95% CI: 95% confidence interval.

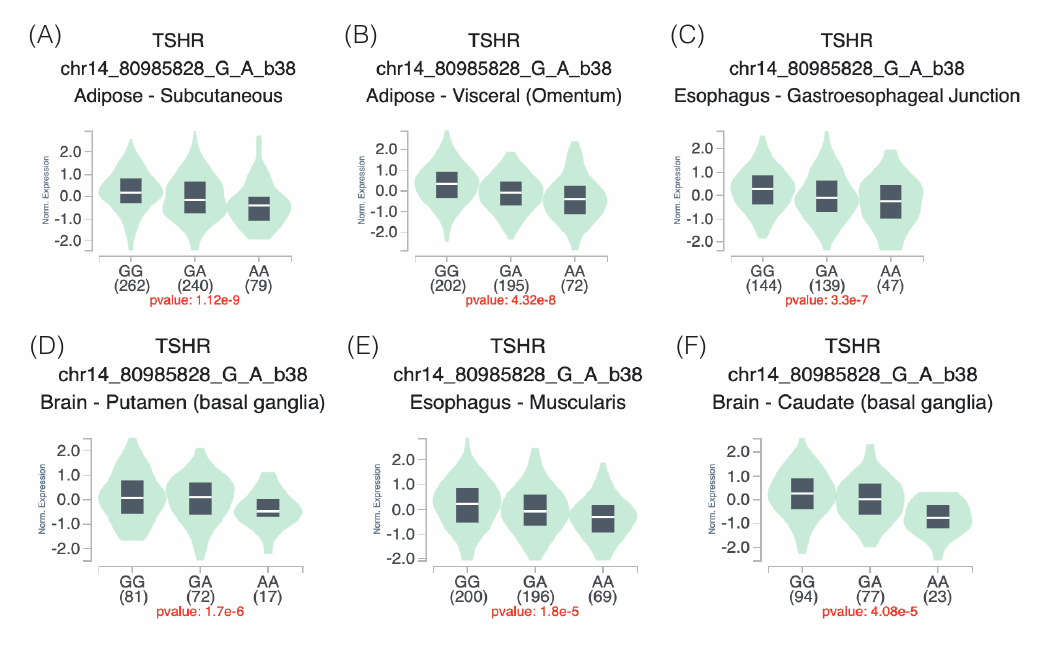

#### Figure S10. eQTL violin plot for the chr14: 80985828, rs17111346 TSHR locus obtained from GTEx Portal ( <https://gtexportal.org/home/snp/rs17111346>).

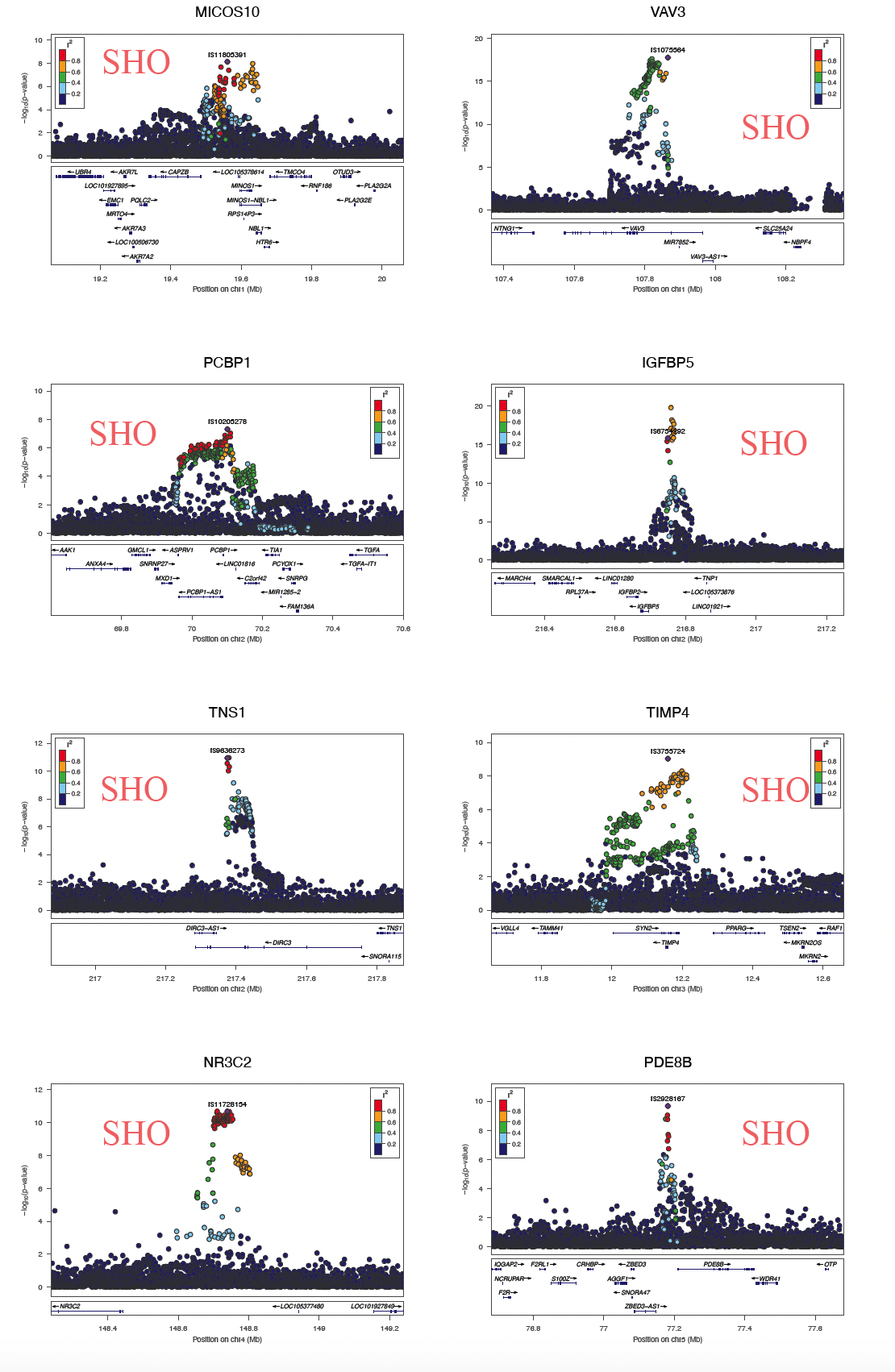

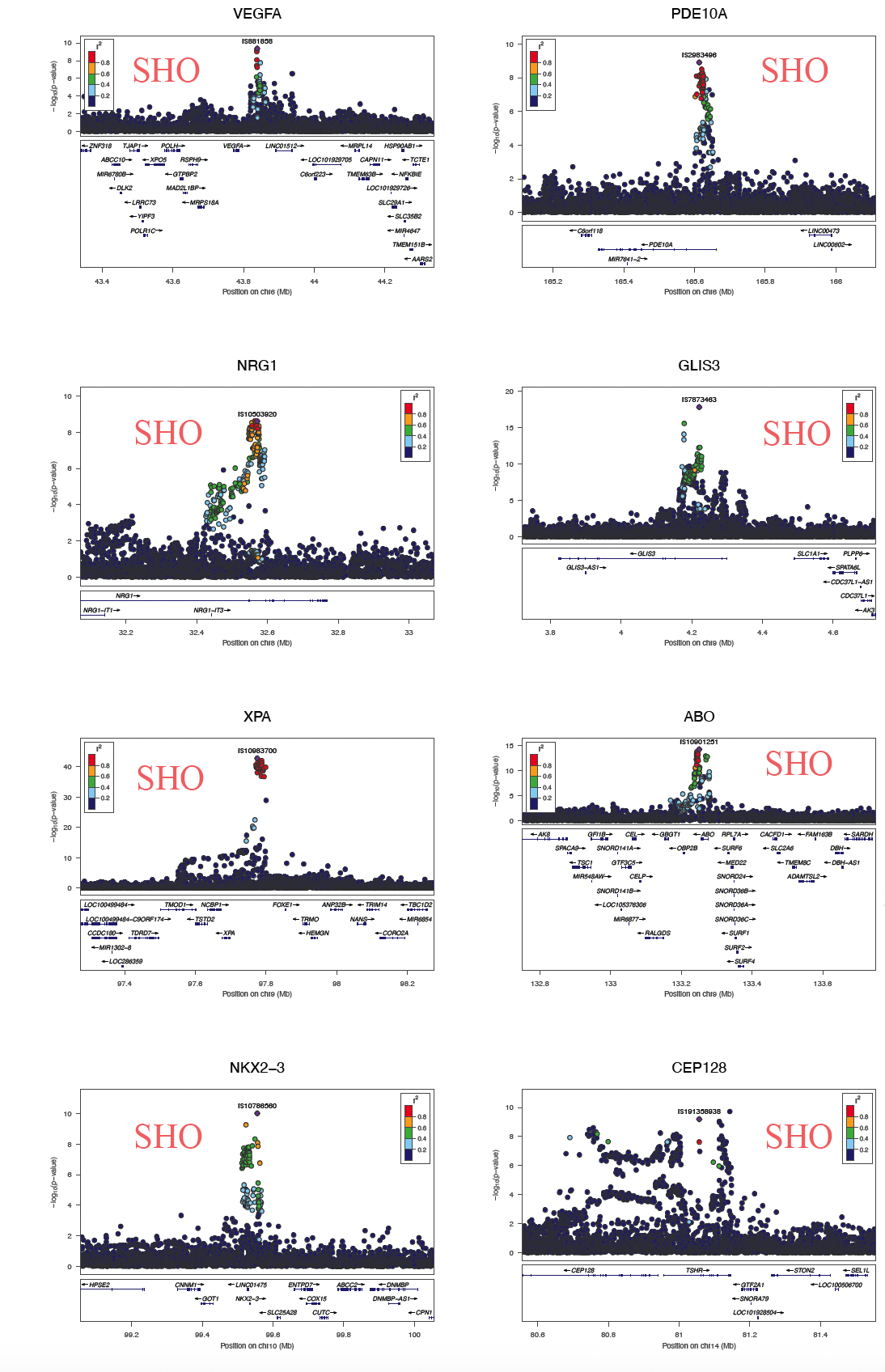

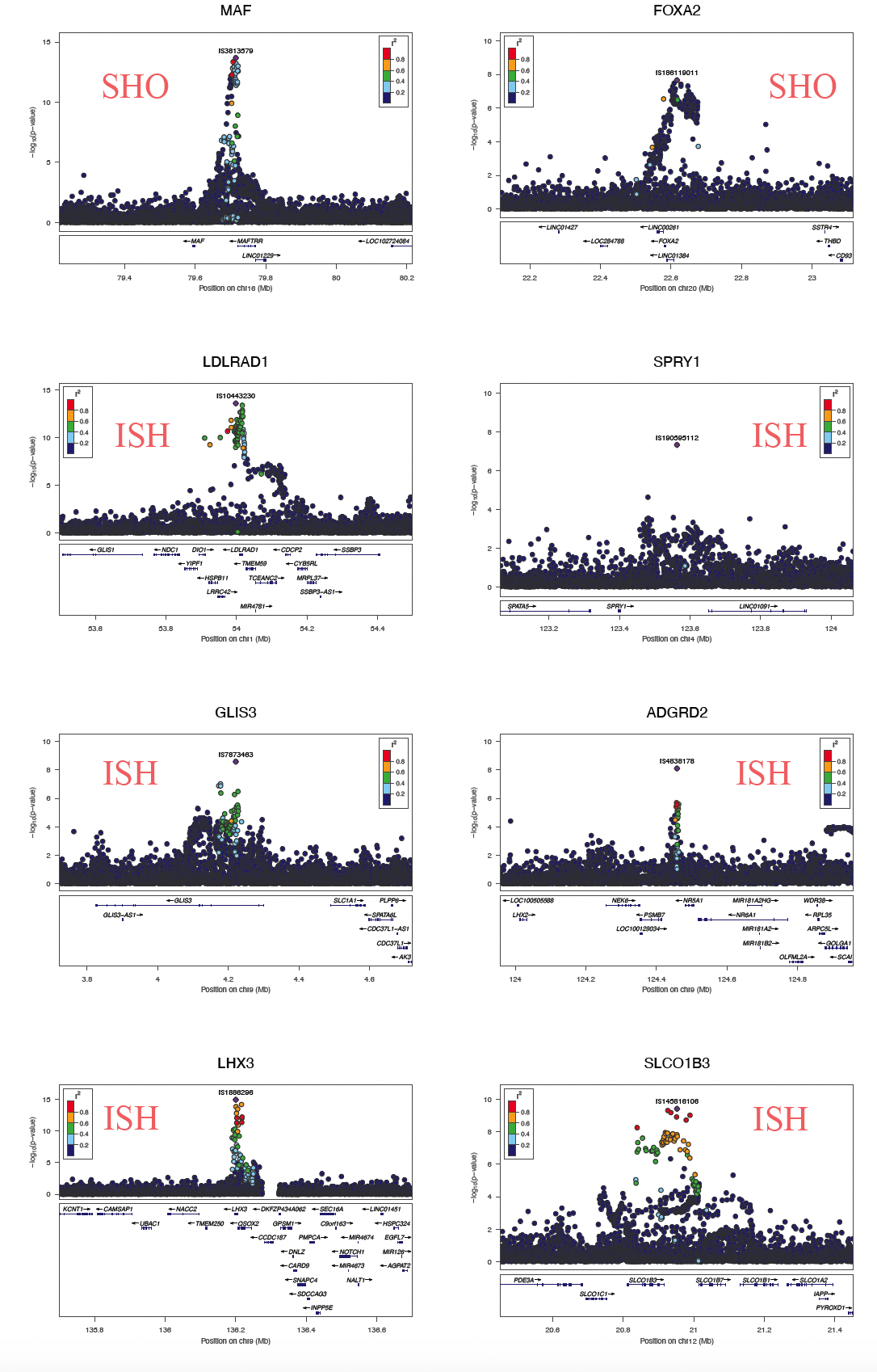

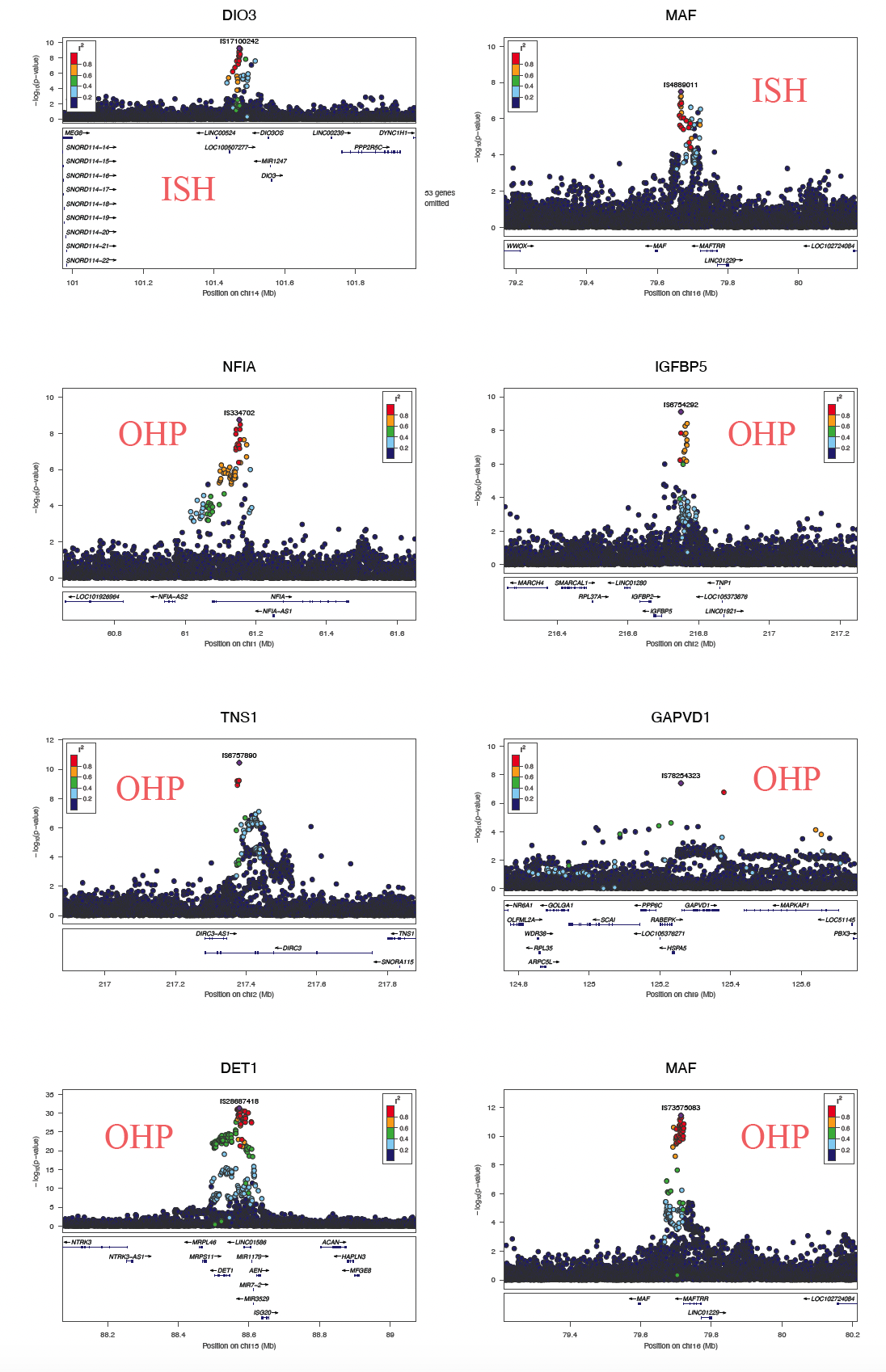

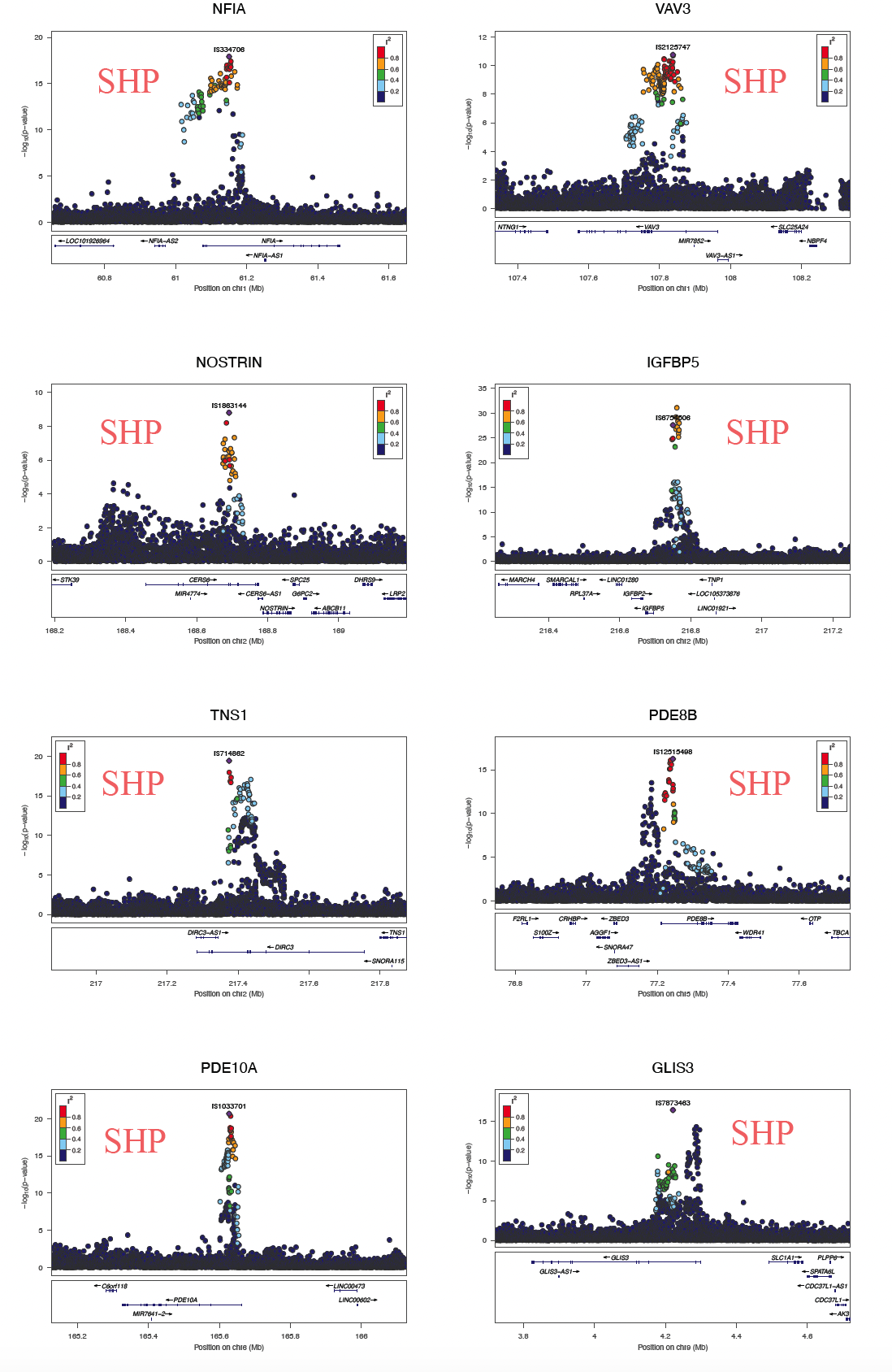

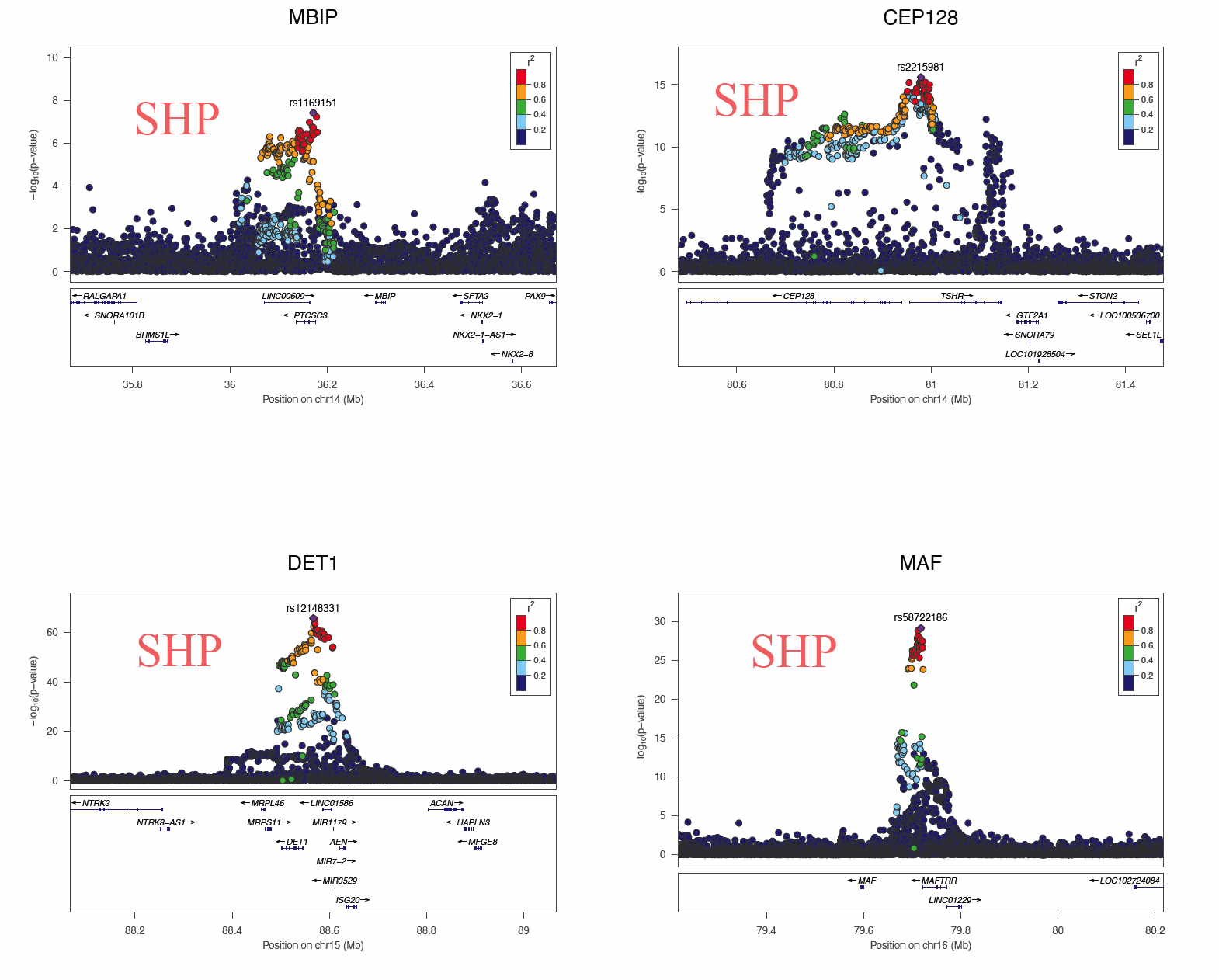

#### Figure S11. Locuszoom plots of genome-wide significant and novel loci associated with thyroid dysfunctions in the study.

For all 44 novel lead SNPs listed in **Table S9**, the regional plots display the P-value and the LD R^2^ for SNPs located in the 500 kbp flanking regions upstream and downstream, generated using LocusZoom software.

Abbreviations: SHO, subclinical hypothyroidism during pregnancy; ISH, isolated hypothyroxinemia during pregnancy; OHP, overt hyperthyroidism during pregnancy; SHP, subclinical hyperthyroidism during pregnancy.

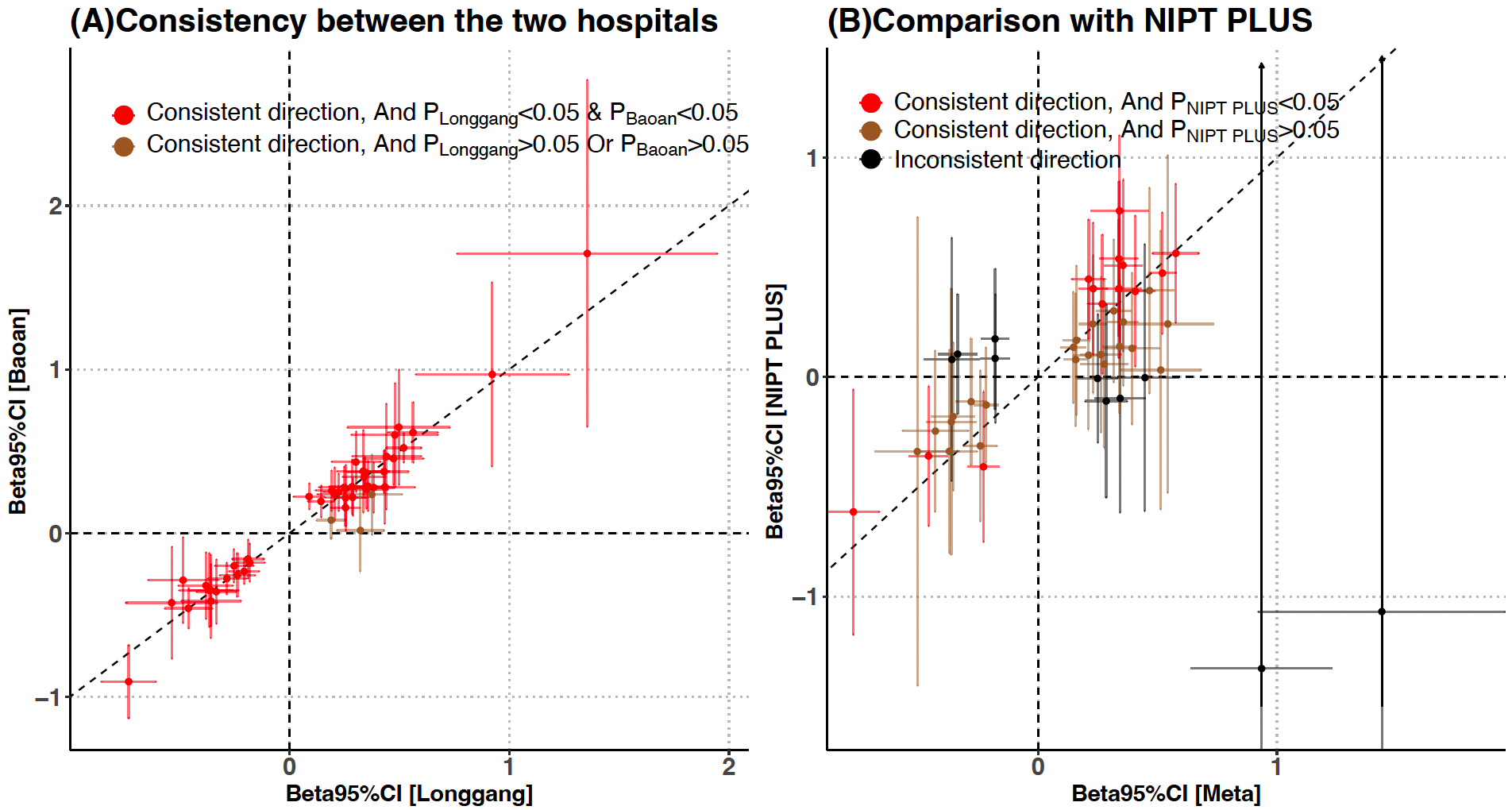

#### Figure S12. Graphical representation for comparison of effect size estimates of 45 thyroid dysfunction-associated loci between (A) Longgang Study and Baoan Study (B)GWAS meta and NIPT PLUS.

Beta, effect size of the GWAS; 95% CI, 95% confidence interval.

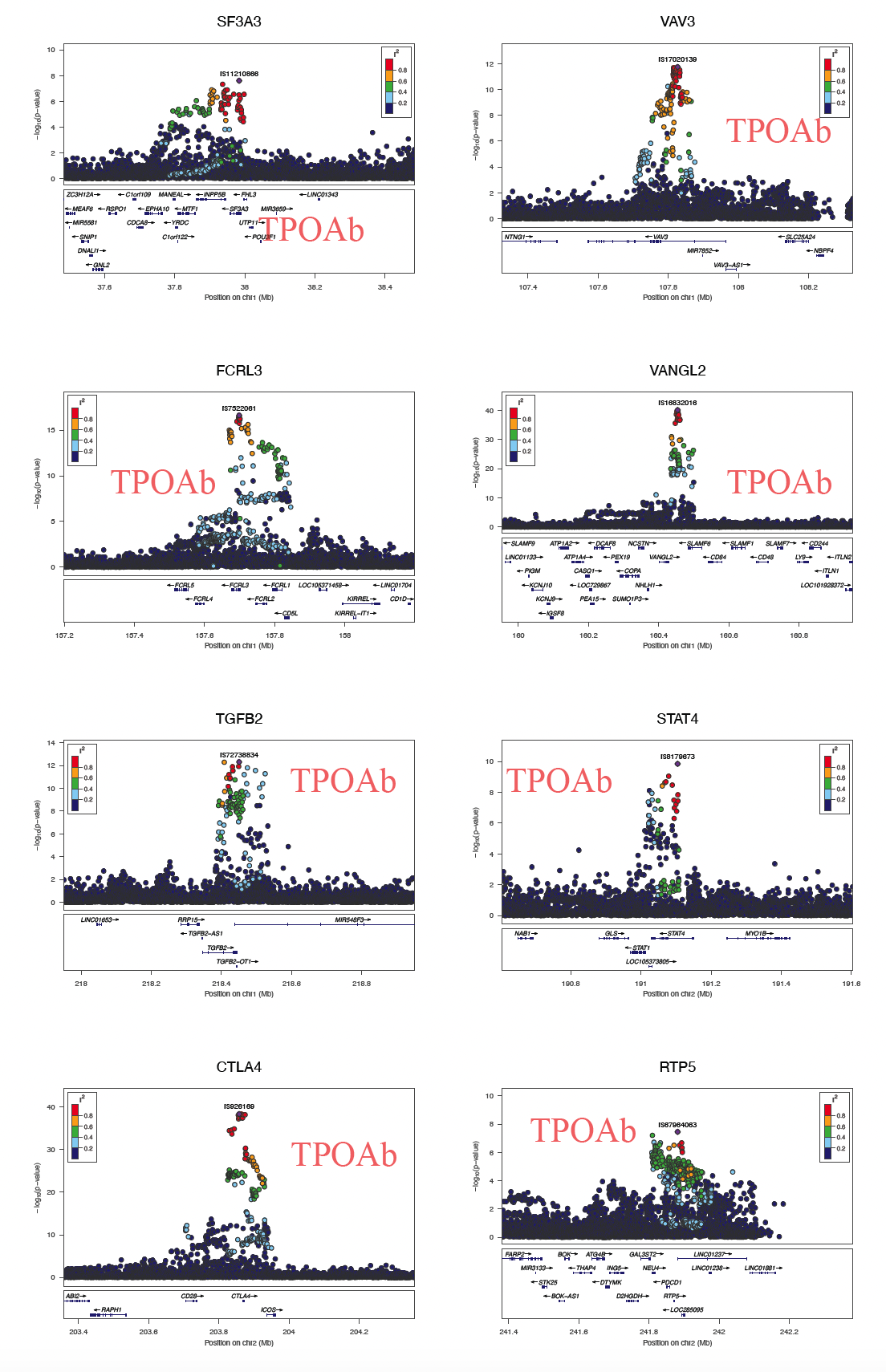

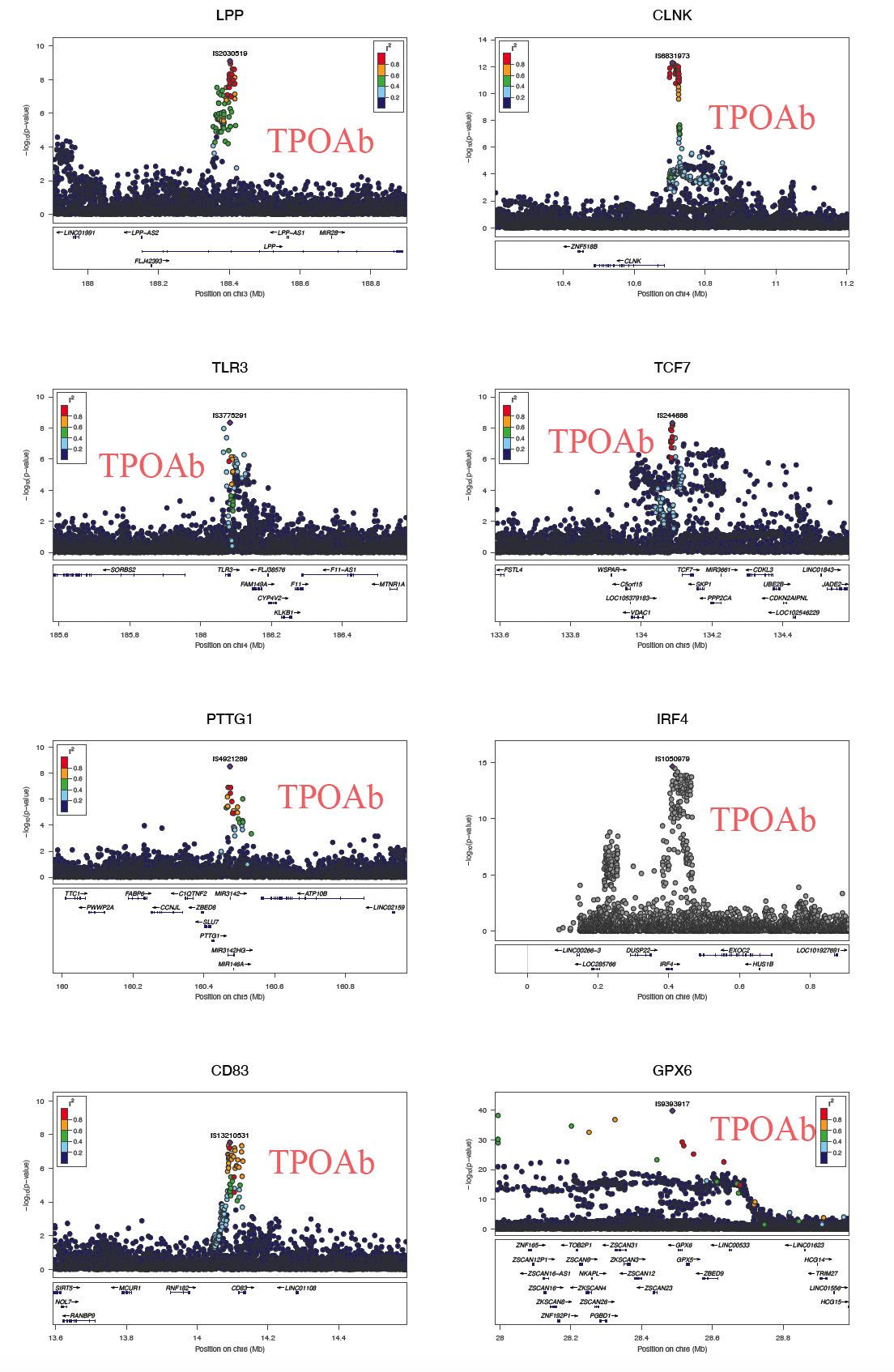

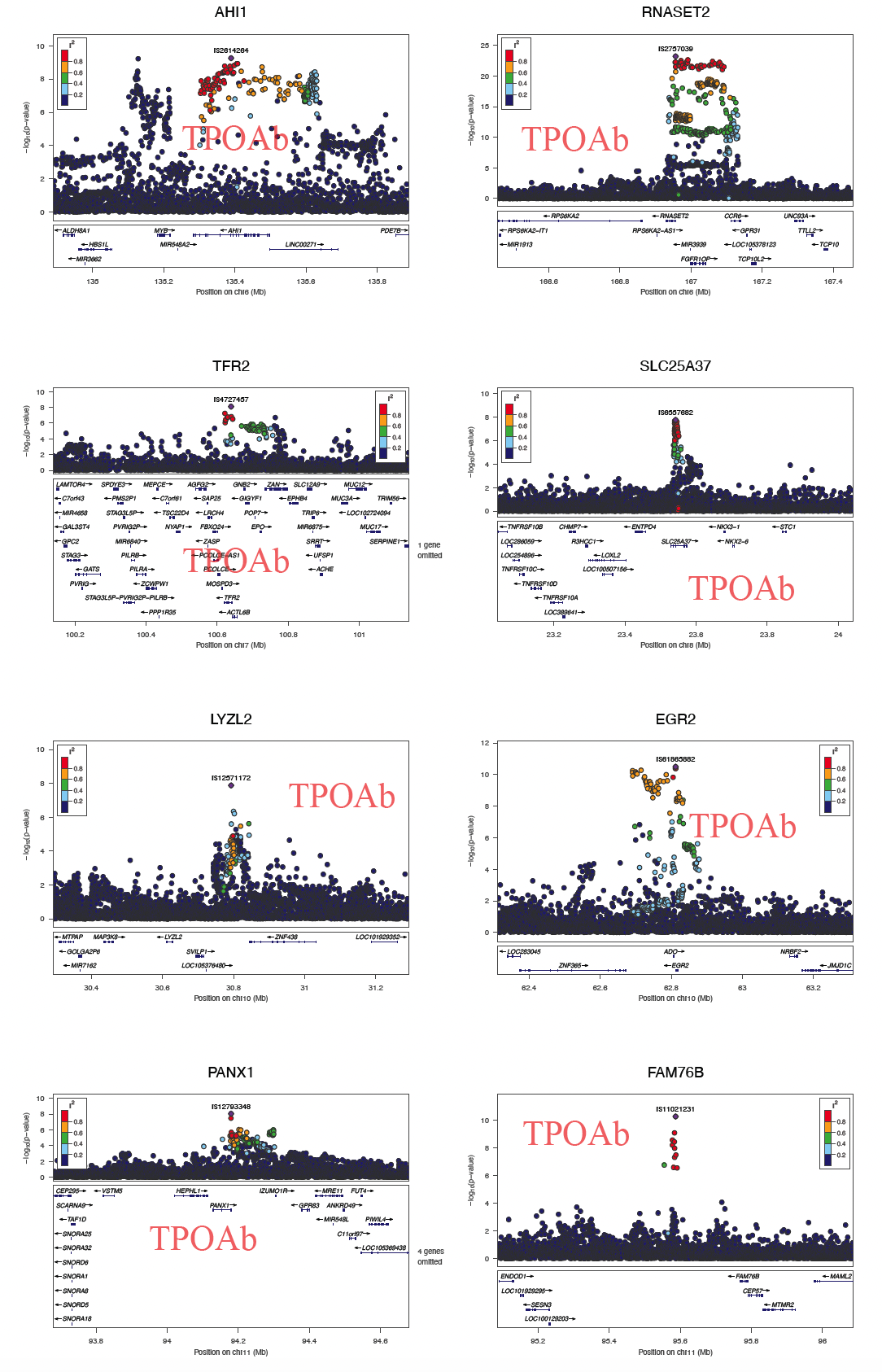

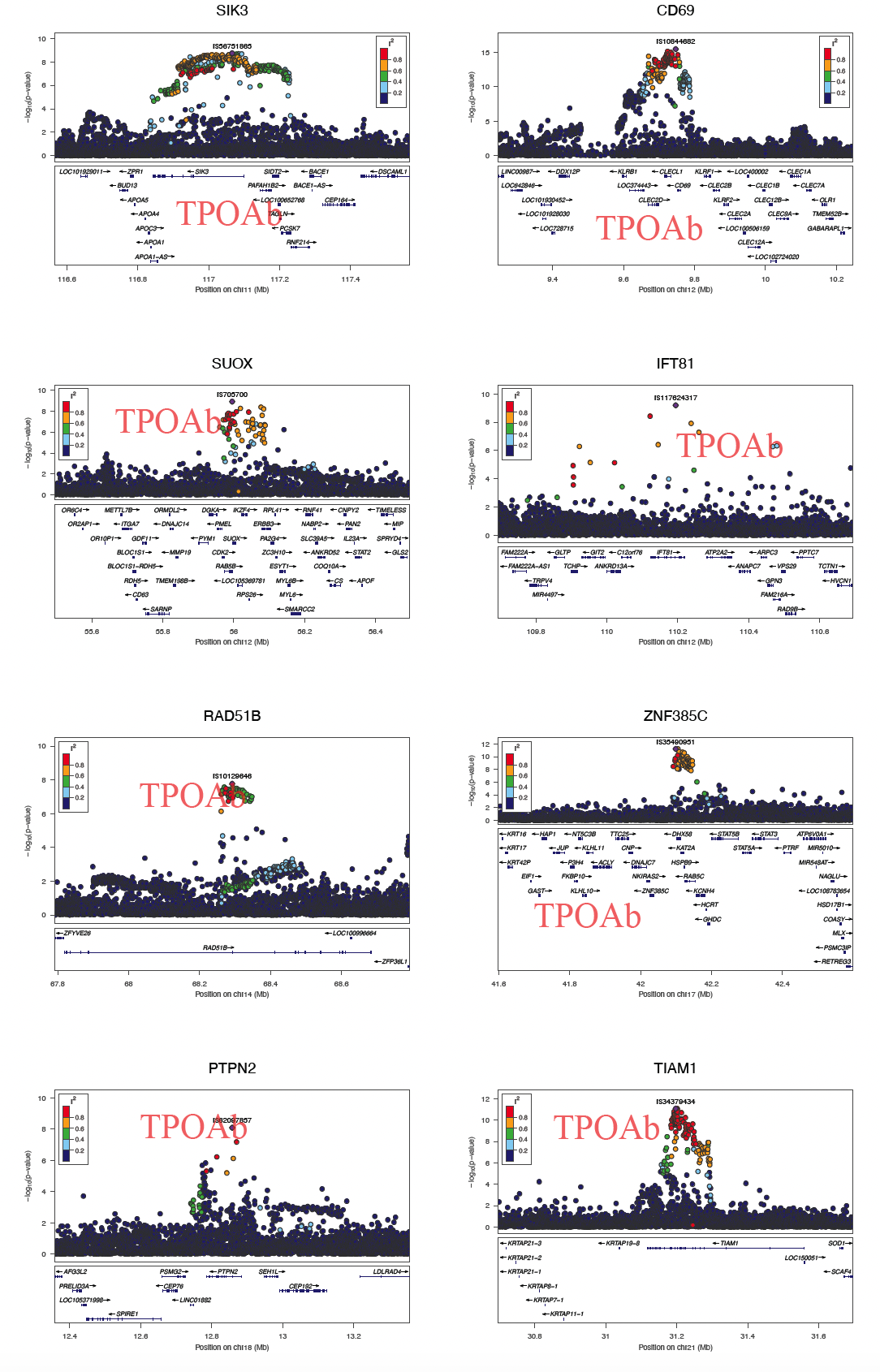

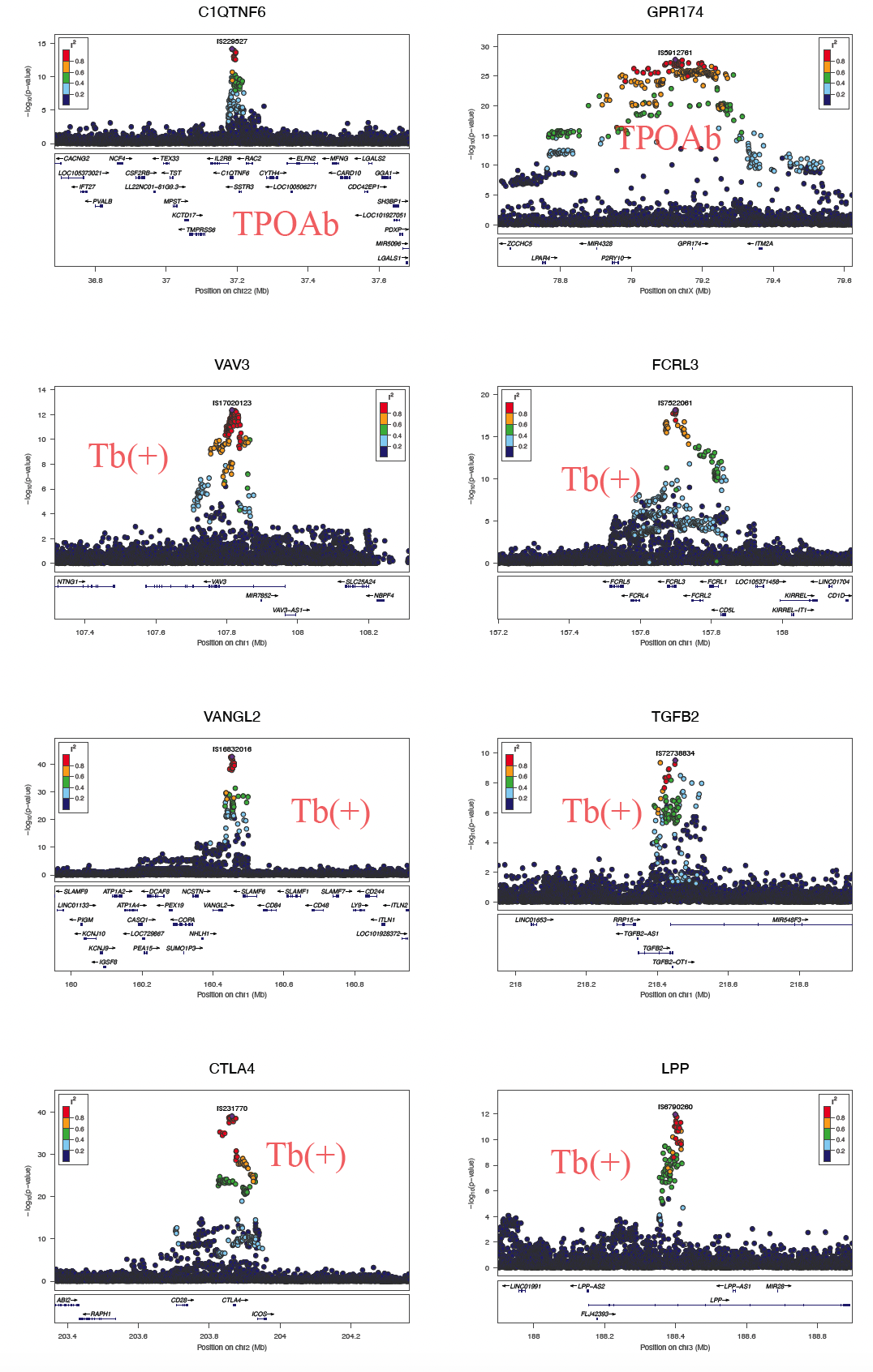

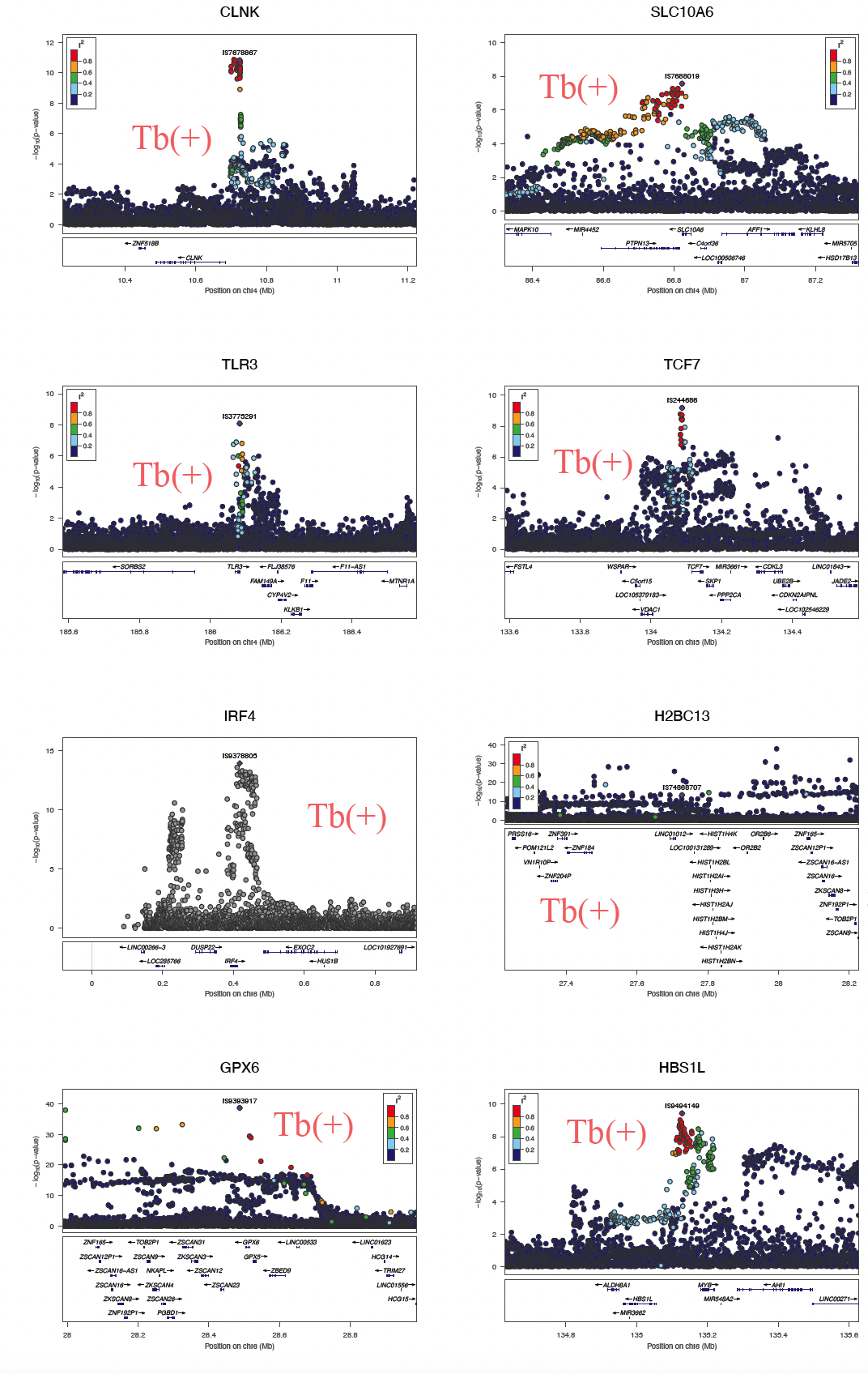

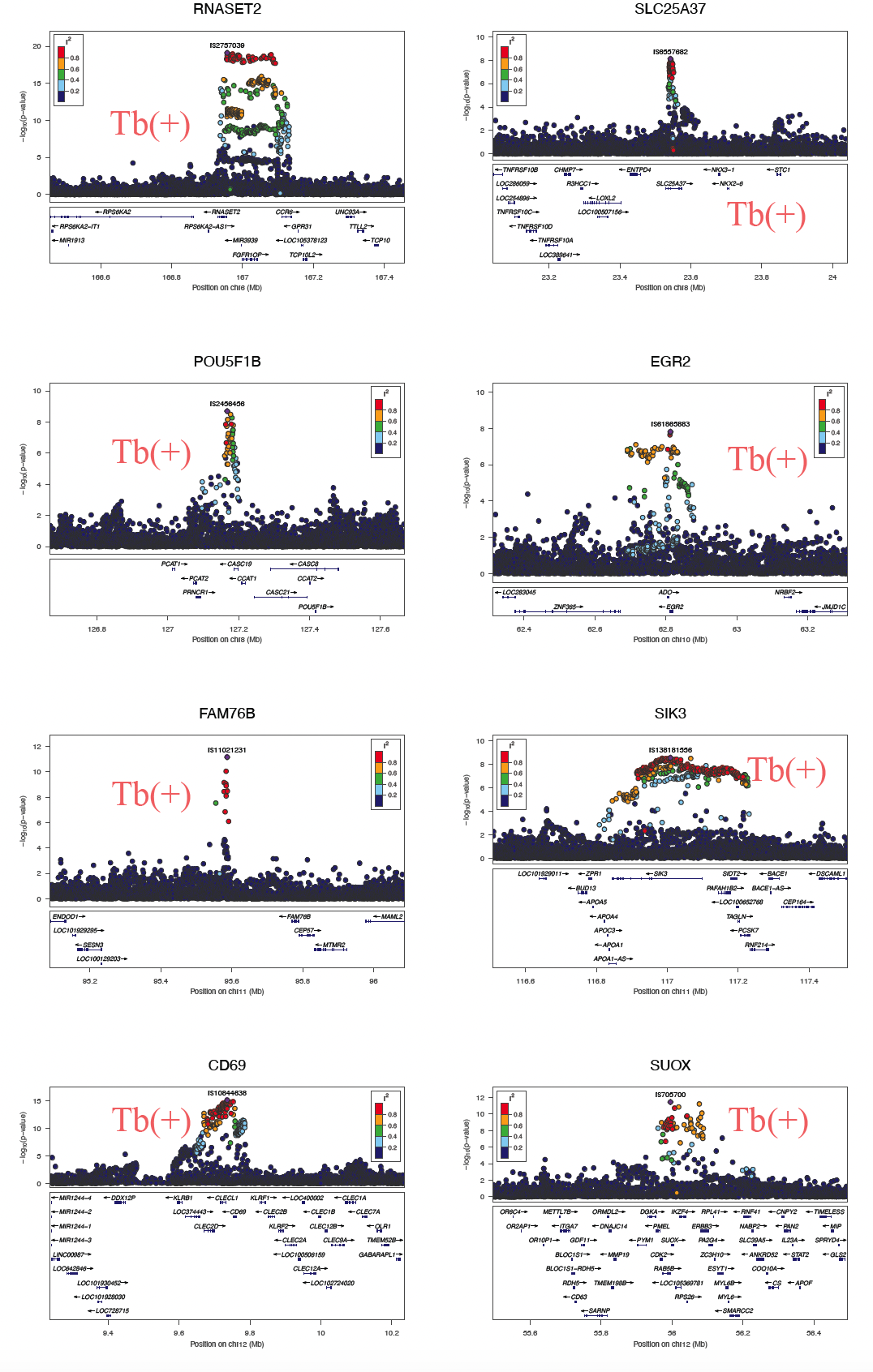

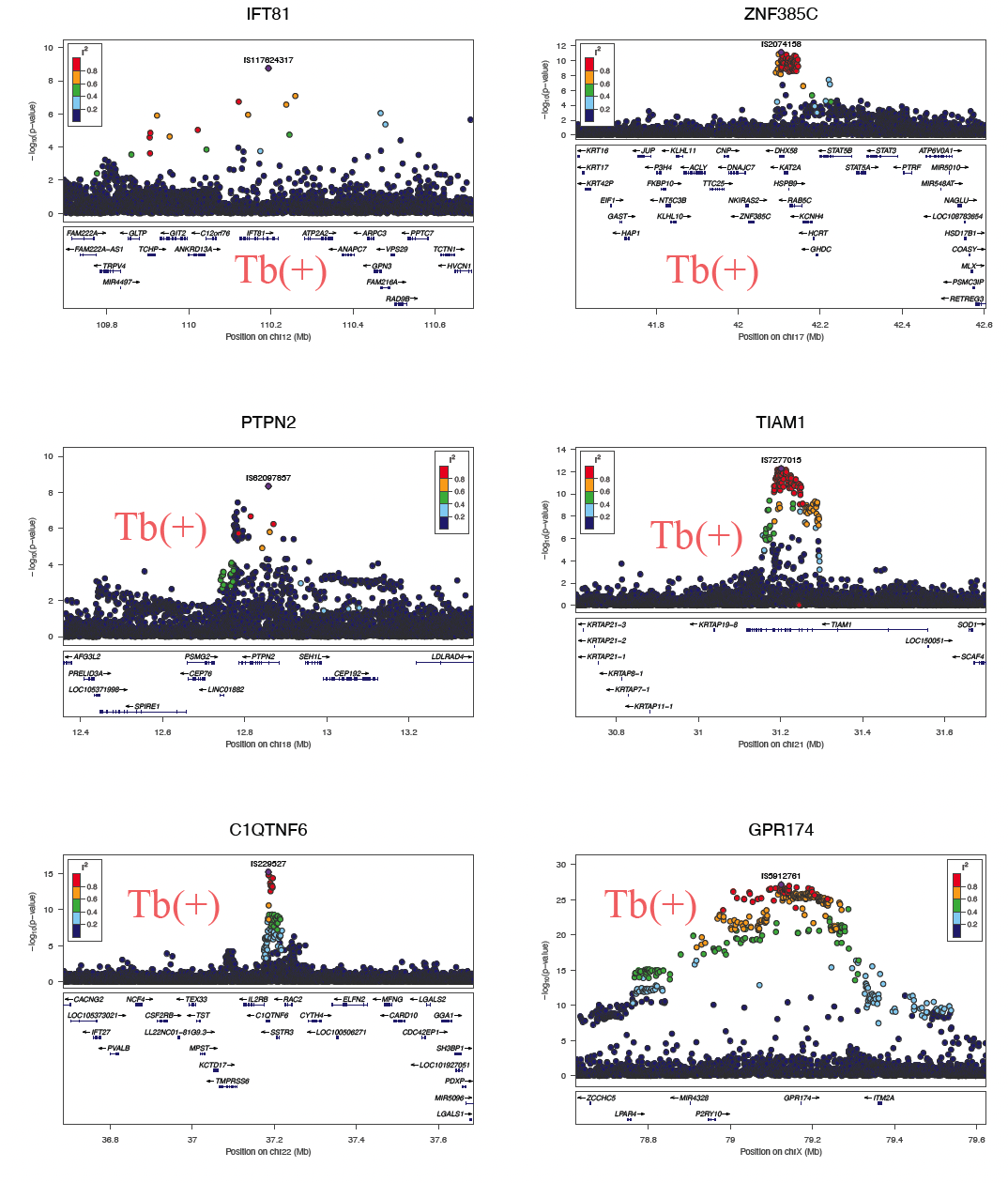

#### Figure S13. Locuszoom plots of genome-wide significant and novel loci associated with TPOAb levels/ positivity in the study.

For the 35 novel lead SNPs in **Table S10**, regional plots showing the P-value and the LD R^2^ of SNPs located in the upstream and downstream 500kbp flanking region are demonstrated using the Locuszoom software. We were unable to generate the LocusZoom plot for *H2BC8* locus (lead SNP: rs1051365311) due to a lack of LD information.

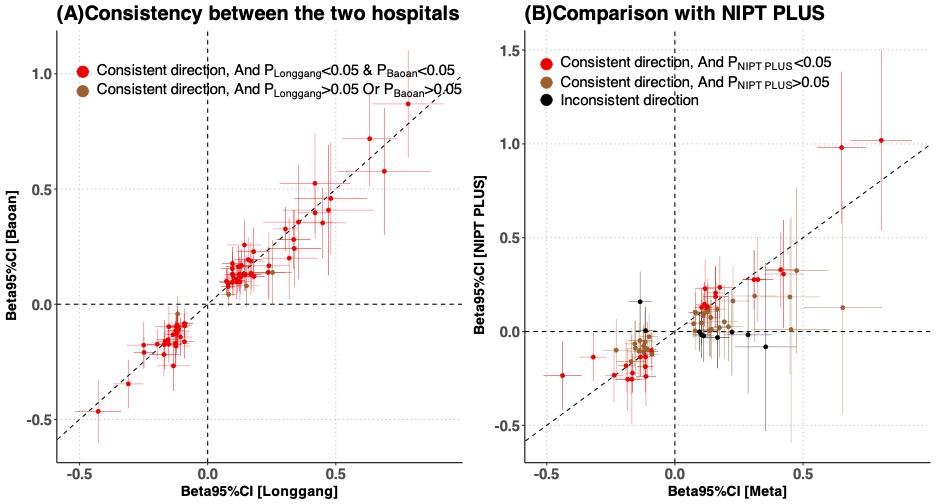

#### Figure S14. Graphical representation for comparison of effect size estimates of 71 TPOAb levels/positivity associated loci between (A) Longgang Study and Baoan Study, (B)GWAS meta and NIPT PLUS.

Beta: effect size of the GWAS; 95% CI: 95% confidence interval.

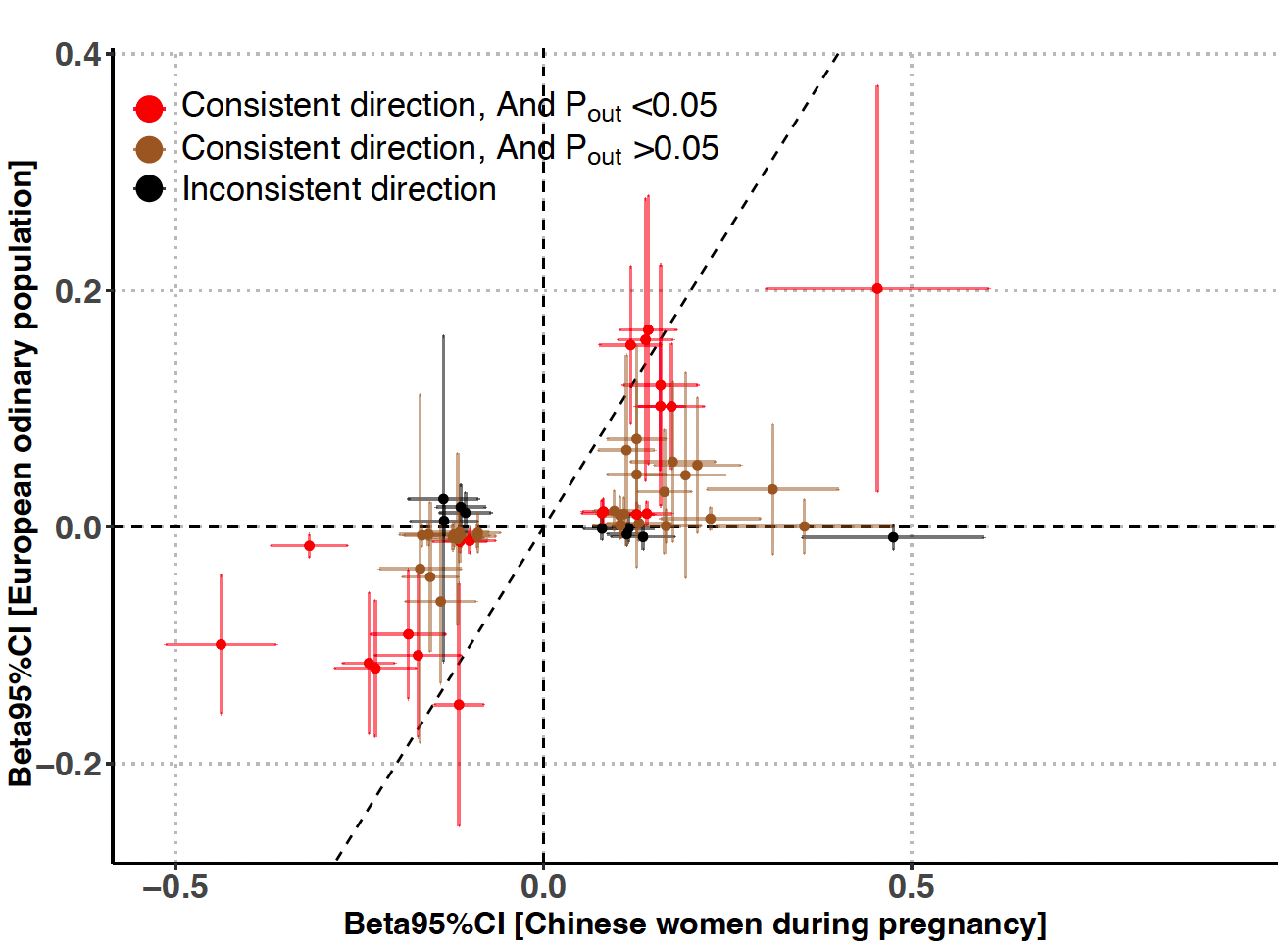

#### Figure S15. Graphical representation for comparison of effect size estimates of TPOAb levels/positivity associated loci between GWAS meta and European population.

Among the 71 loci, 58 loci (or their proxy SNPs) are available in European data.

Beta: effect size of the GWAS; 95% CI: 95% confidence interval.

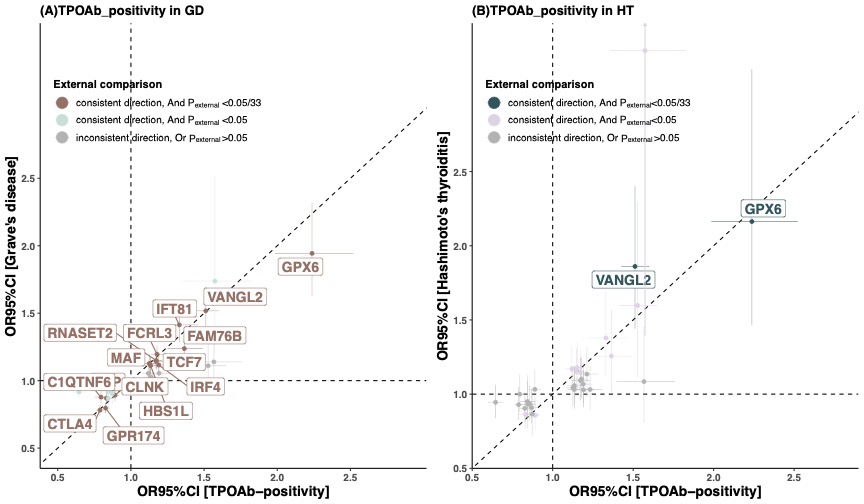

#### Figure S16. Graphical representation for external comparison of TPOAb positivity in BBJ study.

Shown are effect sizes and 95% confidence intervals for (A) the 33 loci in our Meta-GWAS and external GWAS summary data of Graves’ disease (GD) from the BBJ Study, and (B) the 33 loci in our Meta-GWAS and external GWAS summary data of Hashimoto’s thyroiditis (HT) from the BBJ Study. The colors of the dots and gene labels vary depending on whether the external comparison was successful.

OR: odd ratio of the GWAS; 95% CI: 95% confidence interval.

#### Figure S17. Estimated genetic correlations between maternal thyroid-related traits

Due to the large variance in the heritability estimates for overt hyperthyroidism (OHP), LDSC software was unable to calculate the genetic correlation between OHP and other thyroid traits.

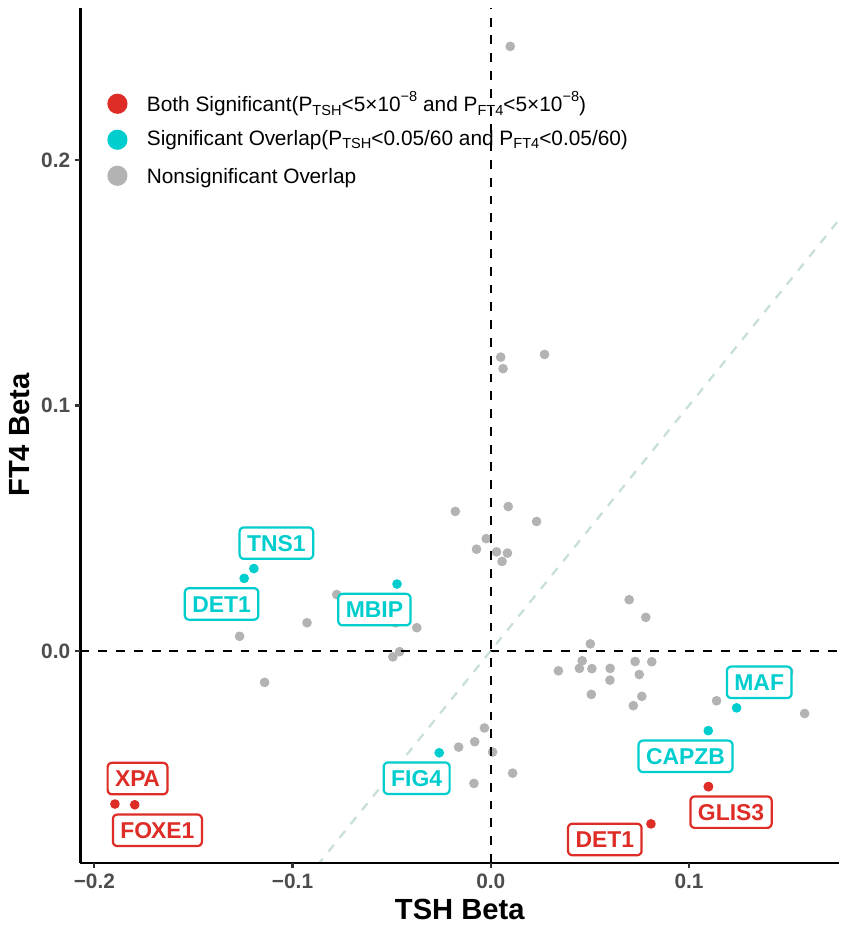

#### Figure S18. The overlap associations between genome-wide variants of TSH and FT4 levels.

The overlapping associations (Bonferroni-corrected threshold p < 0.05/60) could be observed at 11 loci (TSH: *CAPZB, TNS1, GLIS3, XPA, MBIP, DET1, MAF,* and FT4: *FIG4, GLIS3, FOXE1, DET1*). Beta: effect size of the GWAS.

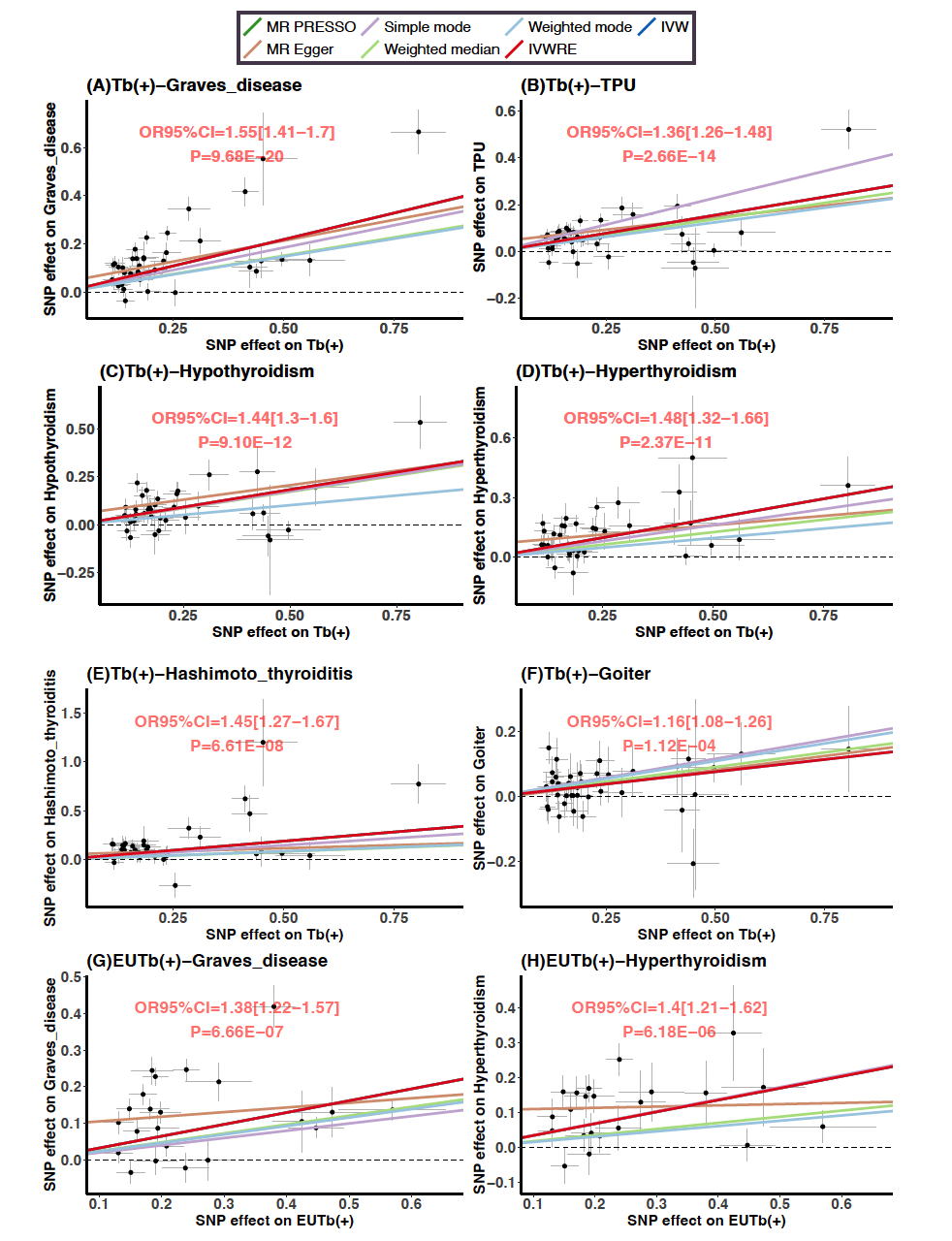

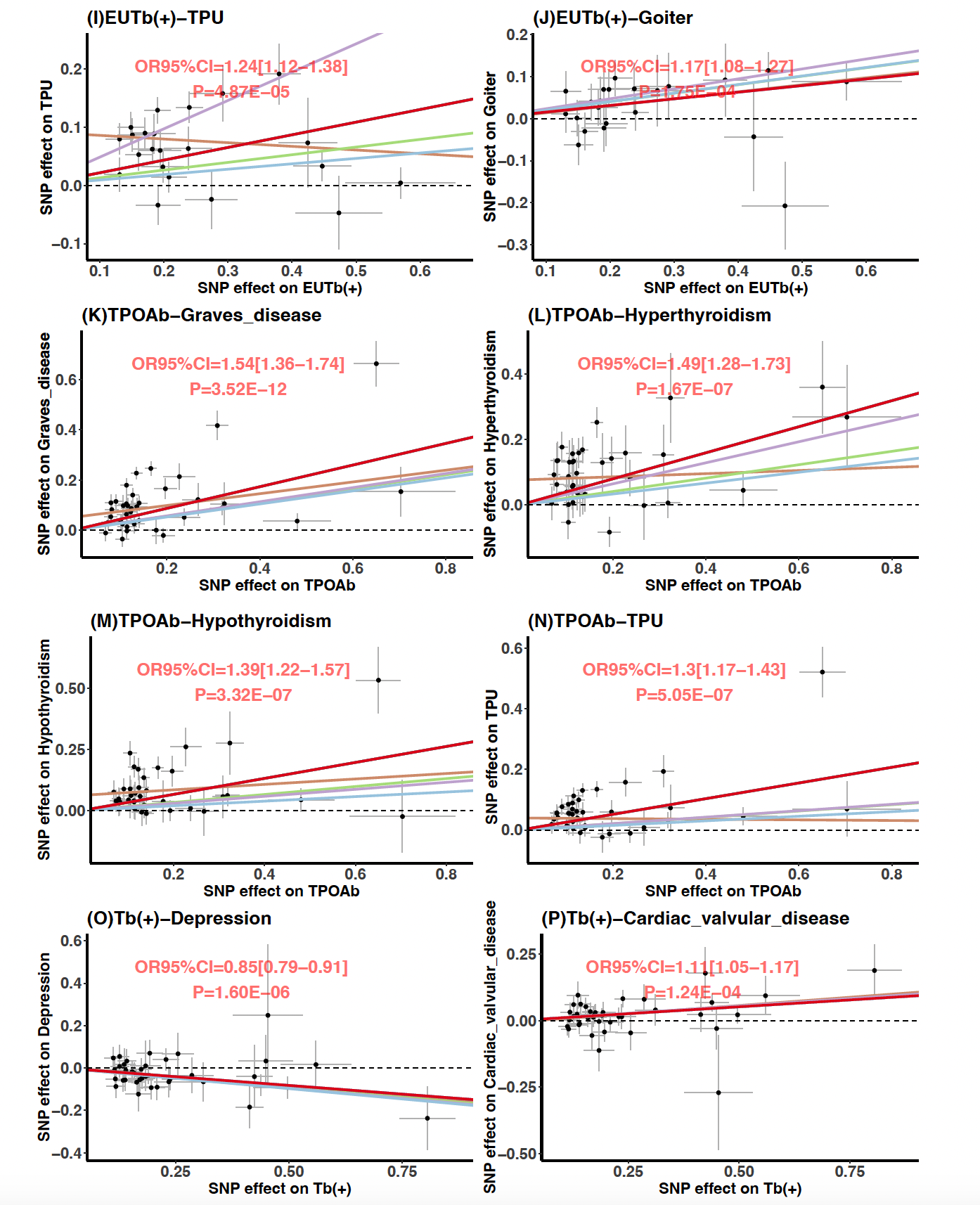

#### Figure S19. Scatter plots of mendelian randomization analysis results of TPOAb traits in this study and thyroid-related diseases(A~N), Depression(G), and cardiac valvular disease in BBJ study.

**1. Figure A~F:** Mendelian randomization analyses results between TPOAb positivity and Graves’ disease (A), Thyroid preparations Usage(B), Hypothyroidism (C), Hyperthyroidism(D), Hashimoto thyroiditis(E), and Goiter(F).

**2. Figure G~J:** Mendelian randomization analyses results between TPOAb with positivity with normal functioning thyroids(ie, euthyroid) and Grave disease (G), Hyperthyroidism(H), Thyroid preparations Usage(I), Goiter(J).

**3. Figure K~N:** Mendelian randomization analyses results between TPOAb levels and Grave disease(K), Hyperthyroidism(L), Hypothyroidism(M), Thyroid preparations Usage(N).

**4. Figure O~P:** Mendelian randomization analyses results between TPOAb positivity and depression(O), cardiac valvular disease(P).

Abbreviation: TPU: Medication usage of thyroid preparations.

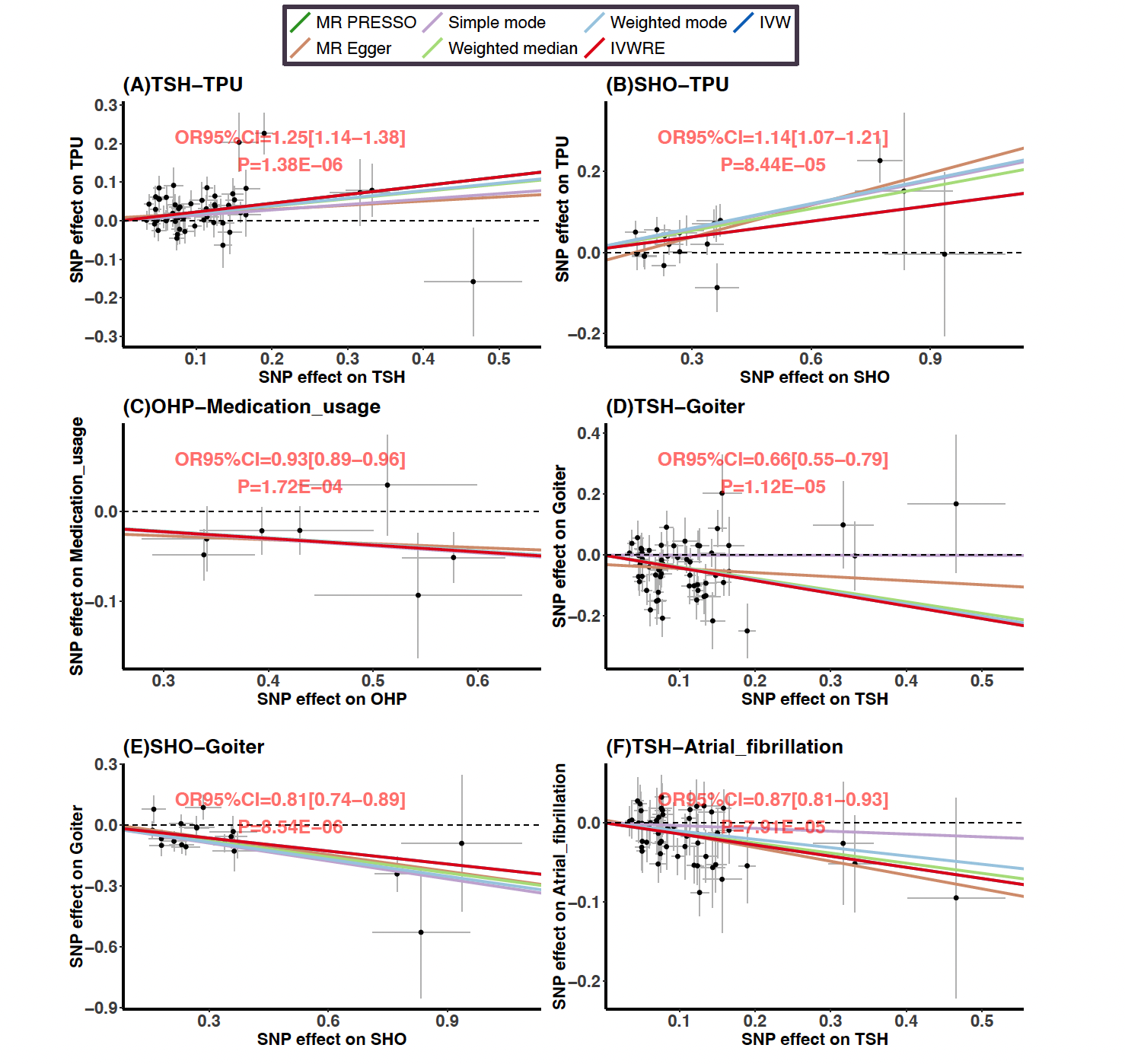

#### Figure S20. Scatter plots of Mendelian randomization analysis results of thyroid-related hormones and dysfunctions during pregnancy in this study and thyroid-related diseases in BBJ study.

(A) TSH and medication usage of thyroid preparations; (B) subclinical hypothyroidism and medication usage of thyroid preparations; (C) overt hyperthyroidism and medication usage of thyroid preparations; (D) TSH and goiter;

(E) subclinical hypothyroidism and goiter; (F) TSH and atrial fibrillation.

Abbreviation: TPU: Medication usage of thyroid preparations.
